# How dangerous is omicron and how effective are vaccinations?

**DOI:** 10.1101/2022.01.27.22269909

**Authors:** Igor Nesteruk, Oleksii Rodionov

**Affiliations:** Institute of Hydromechanics, National Academy of Sciences of Ukraine, Kyiv, Ukraine; Igor Sikorsky Kyiv Polytechnic Institute, Kyiv, Ukraine; Private consulting office, Kyiv, Ukraine

**Keywords:** COVID-19 pandemic, epidemic dynamics in Ukraine, EU, USA, the UK, India, Brazil, South Africa, Argentina, and Australia, efficiency of vaccinations, mathematical modeling of infection diseases, statistical methods

## Abstract

The sharp increase in the number of new COVID-19 cases in late 2021 and early 2022, which is associated with the spread of a new strain of coronavirus - omicron - is of great concern and makes it necessary to make at least approximate forecasts for the pandemic dynamics of the epidemic. As this rapid growth occurs even in countries with high levels of vaccinations, the question arises as to their effectiveness. The smoothed daily number of new cases and deaths per capita and the ratio of these characteristics were used to reveal the appearance of new coronavirus strains and to estimate the effectiveness of quarantine, testing and vaccination. The third year of the pandemic allowed us to compare the pandemic dynamics in the period from September 2020 to January 2021 with the same period one year later for Ukraine, EU, the UK, USA, India, Brazil, South Africa, Argentina, Australia, and in the whole world. Record numbers of new cases registered in late 2021 and early 2022 once again proved that existing vaccines cannot prevent new infections, and vaccinated people can spread the infection as intensively as non-vaccinated ones. Fortunately, the daily number of new cases already diminishes in EU, the UK, USA, South Africa, and Australia. In late January - early February 2022,the maximum averaged numbers of new cases are expected in Brazil, India, EU, and worldwide. “Omicron” waves can increase the numbers of deaths per capita, but in highly vaccinated countries, the deaths per case ratio significantly decreases.

## Introduction

The averaged daily numbers of new COVID-19 cases per capita (DCC) and deaths per capita (DDC) may indicate the effectiveness of quarantine, testing, vaccination, treatment, and also the virulence of coronavirus strains circulating in particular regions at some fixed periods of time. The DCC and DDC values can be calculated with the use of the accumulated number of cases per capita (CC) and deaths per capita (DC) and the simple smoothing procedure proposed in [1-3]. The CC and DC numbers are regularly reported by the World Health Organization, [4] and COVID-19 Data Repository by the Center for Systems Science and Engineering (CSSE) at Johns Hopkins University (JHU), [5].

Since we are already in the third year of the pandemic, it is reasonable to compare the DCC and DDC numbers for the same periods in 2020, 2021, and 2022 in order to find some seasonal and global trends. Since the influence of vaccinations in 2020 and early 2021 can be neglected, the comparison of the COVID-19 pandemic dynamics for the same period one year later enables us to reveal some influences of vaccination levels.

In [6] the DCC values for Argentina, Brazil, India, South Africa, Ukraine, EU, the UK, USA and the whole world for the period from March to August 2020 were compared with the corresponding values in 2021. In this paper we will compare the DCC, DDC, DDC/DCC values and vaccination levels in the same countries and Australia for the period from September to January in 2020-2021 and 2021-2022, and will try to reveal some trends and make some predictions.

### Data and smoothing procedure

The CC figures (per 1.000.000 persons of population) registered by JHU, [5] are shown in Table 1 (August 25, 2020 – January 25, 2021) and Table 2 (August 25, 2021 – January 20, 2022). The accumulated numbers of deaths per capita DC (per million) are shown in Table 3 (August 25, 2020 – January 25, 2021) and Table 4 (August 25, 2021 – January 20, 2022). We denote these values as *V*_*j*_ corresponding time moments *t*_*j*_ measured in days. Similar to the approach proposed in [1-3], we will use the averaged CC and DC values which can be calculated with the use of smoothing

**Table 1.** Accumulated number of laboratory-confirmed COVID-19 cases per million in different regions for the period August 25. 2020 - January 25. 2021. [5].

|  | Argentina | Brazil | India | South Africa | United States | Ukraine | European Union | United Kingdom | Australia | World |
| --- | --- | --- | --- | --- | --- | --- | --- | --- | --- | --- |
| 2020-08-25 | 7885.791 | 17188.75 | 2314.142 | 10209.8 | 17403.84 | 2552.499 | 4014.002 | 4815.011 | 977.346 | 3039.414 |
| 2020-08-26 | 8117.121 | 17413.76 | 2375.637 | 10254.51 | 17531.85 | 2591.701 | 4049.832 | 4830.42 | 981.921 | 3075.218 |
| 2020-08-27 | 8338.672 | 17611.25 | 2431.088 | 10297.56 | 17682.18 | 2637.943 | 4116.519 | 4852.734 | 986.807 | 3111.653 |
| 2020-08-28 | 8595.591 | 17817.27 | 2485.969 | 10328.3 | 17823.12 | 2695.665 | 4171.747 | 4871.471 | 990.646 | 3147.77 |
| 2020-08-29 | 8797.977 | 17991.53 | 2542.493 | 10368.59 | 17952.86 | 2754.998 | 4201.316 | 4887.716 | 995.416 | 3180.812 |
| 2020-08-30 | 8955.567 | 18059.45 | 2598.838 | 10410.31 | 18053.82 | 2805.128 | 4227.249 | 4912.86 | 998.363 | 3208.572 |
| 2020-08-31 | 9159.686 | 18294.15 | 2649.018 | 10443.37 | 18155.54 | 2855.787 | 4301.327 | 4933.474 | 1001.194 | 3242.299 |
| 2020-09-01 | 9390.007 | 18501.31 | 2705.252 | 10463.66 | 18280.52 | 2905.181 | 4346.102 | 4952.533 | 1005.227 | 3276.125 |
| 2020-09-02 | 9629.735 | 18718.38 | 2765.452 | 10502.57 | 18401.24 | 2963.939 | 4400.627 | 4974.657 | 1010.112 | 3311.959 |
| 2020-09-03 | 9893.43 | 18937.12 | 2825.263 | 10542.87 | 18537.09 | 3020.695 | 4457.715 | 5000.094 | 1013.486 | 3348.215 |
| 2020-09-04 | 10127.7 | 19121.89 | 2887.292 | 10577.23 | 18682.07 | 3084.398 | 4523.528 | 5028.537 | 1016.239 | 3386.472 |
| 2020-09-05 | 10345.3 | 19280.11 | 2952.336 | 10607.31 | 18810.95 | 3151.047 | 4562.73 | 5055.118 | 1018.993 | 3421.075 |
| 2020-09-06 | 10498.48 | 19348.67 | 3017.501 | 10634.51 | 18901.63 | 3201.775 | 4595.021 | 5098.852 | 1020.66 | 3450.237 |
| 2020-09-07 | 10700.54 | 19393.14 | 3071.906 | 10648.58 | 18976.31 | 3253.608 | 4682.769 | 5142.103 | 1022.676 | 3477.969 |
| 2020-09-08 | 10964.26 | 19480.92 | 3136.285 | 10666.55 | 19055.63 | 3310.893 | 4739.078 | 5178.184 | 1026.244 | 3509.175 |
| 2020-09-09 | 11233.06 | 19654.23 | 3204.991 | 10699.69 | 19155.55 | 3370.64 | 4802.098 | 5217.198 | 1028.532 | 3545.405 |
| 2020-09-10 | 11494.1 | 19844.71 | 3274.282 | 10733.12 | 19273.23 | 3431.261 | 4874.947 | 5259.994 | 1030.083 | 3583.921 |
| 2020-09-11 | 11746.42 | 20038.8 | 3344.304 | 10765.77 | 19408.51 | 3505.501 | 4951.943 | 5311.909 | 1031.75 | 3624.119 |
| 2020-09-12 | 11982.7 | 20182.73 | 3412.032 | 10796.01 | 19544.62 | 3578.776 | 5004.536 | 5363.18 | 1033.456 | 3660.71 |
| 2020-09-13 | 12181.27 | 20248.24 | 3478.108 | 10822.31 | 19664.79 | 3637.763 | 5040.662 | 5412.001 | 1035.046 | 3692.16 |
| 2020-09-14 | 12398.55 | 20344.56 | 3538.255 | 10838.23 | 19765.25 | 3696.59 | 5137.857 | 5450.429 | 1036.869 | 3726.224 |
| 2020-09-15 | 12659.3 | 20506.02 | 3602.933 | 10851.09 | 19874.03 | 3765.585 | 5205.187 | 5496.01 | 1038.381 | 3761.776 |
| 2020-09-16 | 12915.28 | 20680.31 | 3673.188 | 10883.12 | 19991.09 | 3834.971 | 5284.936 | 5554.523 | 1039.738 | 3800.512 |
| 2020-09-17 | 13193.78 | 20850.43 | 3742.388 | 10918.56 | 20124.57 | 3919.61 | 5373.701 | 5604.371 | 1041.6 | 3840.403 |
| 2020-09-18 | 13455.69 | 21036.1 | 3809.372 | 10952.78 | 20278.28 | 3996.22 | 5474.028 | 5667.737 | 1042.53 | 3882.044 |
| 2020-09-19 | 13659.09 | 21174.22 | 3875.832 | 10986.58 | 20412.31 | 4073.176 | 5538.159 | 5732.569 | 1043.034 | 3919.015 |
| 2020-09-20 | 13843.96 | 21246.39 | 3938.241 | 11012.48 | 20540.66 | 4143.827 | 5591.09 | 5789.733 | 1043.577 | 3951.463 |
| 2020-09-21 | 14036.52 | 21328.79 | 3992.125 | 11024.55 | 20637.68 | 4207.807 | 5703.237 | 5853.891 | 1044.741 | 3984.547 |
| 2020-09-22 | 14300.24 | 21492.2 | 4051.94 | 11046.97 | 20749.33 | 4276.595 | 5786.515 | 5926.112 | 1045.904 | 4020.548 |
| 2020-09-23 | 14577.06 | 21645.13 | 4114.024 | 11078.71 | 20875.07 | 4359.371 | 5880.189 | 6016.718 | 1046.214 | 4060.134 |
| 2020-09-24 | 14872.36 | 21794.73 | 4175.78 | 11109.71 | 21020.51 | 4439.386 | 5983.603 | 6113.996 | 1046.99 | 4100.801 |
| 2020-09-25 | 15156.73 | 21945.72 | 4237.042 | 11134.36 | 21163.19 | 4523.703 | 6094.567 | 6214.777 | 1047.61 | 4142.783 |
| 2020-09-26 | 15403.38 | 22061.23 | 4300.627 | 11150.5 | 21292.44 | 4614.232 | 6166.619 | 6303.345 | 1048.541 | 4178.765 |
| 2020-09-27 | 15597.24 | 22125.66 | 4359.597 | 11171.61 | 21396.41 | 4688.611 | 6226.31 | 6386.797 | 1048.696 | 4210.245 |
| 2020-09-28 | 15856.13 | 22204.49 | 4410.256 | 11186.65 | 21509.5 | 4752.567 | 6341.679 | 6446.204 | 1049.123 | 4243.652 |
| 2020-09-29 | 16151.64 | 22357.99 | 4468.008 | 11201.69 | 21634.76 | 4838.38 | 6401.848 | 6550.93 | 1050.014 | 4279.443 |
| 2020-09-30 | 16467.22 | 22511.74 | 4530.317 | 11231.12 | 21751.36 | 4933.556 | 6528.595 | 6655.156 | 1050.712 | 4320.748 |
| 2020-10-01 | 16774.22 | 22678.96 | 4588.795 | 11260.19 | 21890.43 | 5029.698 | 6634.293 | 6756.524 | 1051.217 | 4361.2 |
| 2020-10-02 | 17096.26 | 22832.32 | 4645.832 | 11289.32 | 22050.38 | 5139 | 6750.177 | 6858.859 | 1051.682 | 4402.991 |
| 2020-10-03 | 17340.29 | 22939.83 | 4700.252 | 11320.68 | 22198.03 | 5249.084 | 6843.104 | 7047.579 | 1052.225 | 4440.578 |
| 2020-10-04 | 17508.42 | 22980.56 | 4753.676 | 11346.88 | 22297.32 | 5347.159 | 6919.248 | 7384.215 | 1052.729 | 4473.464 |
| 2020-10-05 | 17754.93 | 23102.31 | 4797.645 | 11362.3 | 22409.83 | 5436.997 | 7036.903 | 7568.961 | 1053.698 | 4513.181 |
| 2020-10-06 | 18078.13 | 23249.63 | 4849.352 | 11379.4 | 22542.72 | 5540.111 | 7150.324 | 7782.297 | 1054.009 | 4553.148 |
| 2020-10-07 | 18438.76 | 23394.2 | 4905.706 | 11411.26 | 22690.98 | 5652.541 | 7293.825 | 7989.973 | 1054.978 | 4597.438 |
| 2020-10-08 | 18777.62 | 23519.58 | 4956.298 | 11440.18 | 22864.53 | 5780.11 | 7455.497 | 8247.146 | 1055.753 | 4643.1 |
| 2020-10-09 | 19108.7 | 23649.18 | 5008.883 | 11464.51 | 23030.59 | 5917.249 | 7636.252 | 8450.409 | 1056.451 | 4688.824 |
| 2020-10-10 | 19380.9 | 23760.14 | 5062.265 | 11506.88 | 23196.08 | 6053.008 | 7800.328 | 8672.746 | 1057.188 | 4733.788 |
| 2020-10-11 | 19607.28 | 23817.1 | 5110.156 | 11533.11 | 23334.87 | 6167.118 | 7919.843 | 8861.466 | 1058.041 | 4770.262 |
| 2020-10-12 | 19816.11 | 23857.46 | 5149.873 | 11547.9 | 23466.45 | 6273.083 | 8089.458 | 9066.869 | 1059.011 | 4807.58 |
| 2020-10-13 | 20107.85 | 23919.89 | 5195.451 | 11567.52 | 23615.37 | 6395.269 | 8250.958 | 9319.57 | 1060.058 | 4848.199 |
| 2020-10-14 | 20435.26 | 24050 | 5244.043 | 11598.78 | 23789.17 | 6528.244 | 8466.412 | 9608.807 | 1060.833 | 4896.572 |
| 2020-10-15 | 20810.13 | 24192.19 | 5289.522 | 11628.26 | 23982.13 | 6649.255 | 8718.625 | 9887.092 | 1061.376 | 4948.434 |
| 2020-10-16 | 21172.93 | 24325.82 | 5334.17 | 11661.89 | 24190.08 | 6792.008 | 8971.812 | 10116.32 | 1061.648 | 5000.447 |
| 2020-10-17 | 21469.17 | 24422.9 | 5378.572 | 11694 | 24354.86 | 6944.515 | 9201.75 | 10353.41 | 1062.113 | 5047.258 |
| 2020-10-18 | 21700.74 | 24479.22 | 5418.562 | 11721.68 | 24511.72 | 7069.783 | 9407.694 | 10602.37 | 1062.462 | 5089.167 |
| 2020-10-19 | 21985.39 | 24556.69 | 5452.141 | 11746.01 | 24717.89 | 7184.491 | 9661.658 | 10878.32 | 1062.695 | 5137.209 |
| 2020-10-20 | 22343.62 | 24667.75 | 5490.927 | 11763.5 | 24903.55 | 7315.166 | 9909.978 | 11191.06 | 1064.168 | 5186.684 |
| 2020-10-21 | 22745.45 | 24788.58 | 5531 | 11797.73 | 25085.98 | 7475.633 | 10224.92 | 11582.43 | 1064.75 | 5242.964 |
| 2020-10-22 | 23103.41 | 24904.3 | 5570.017 | 11833.63 | 25321.76 | 7644.037 | 10585.84 | 11893.94 | 1065.448 | 5303.234 |
| 2020-10-23 | 23448.06 | 25044.5 | 5608.319 | 11865.23 | 25569.72 | 7823.024 | 10982.65 | 12195.11 | 1066.185 | 5366.436 |
| 2020-10-24 | 23710.48 | 25161.25 | 5644.295 | 11895.77 | 25802.39 | 7990.393 | 11326.51 | 12532.49 | 1066.883 | 5423.517 |
| 2020-10-25 | 23913.37 | 25218.51 | 5676.696 | 11922.79 | 25987.77 | 8137.77 | 11564.79 | 12822.64 | 1067.348 | 5469.114 |
| 2020-10-26 | 24170.18 | 25305.48 | 5702.869 | 11937.63 | 26182.94 | 8267.179 | 12054.02 | 13128.92 | 1067.891 | 5530.955 |
| 2020-10-27 | 24483.91 | 25445.59 | 5734.369 | 11955.82 | 26413.52 | 8425.576 | 12440.41 | 13464.83 | 1068.434 | 5590.701 |
| 2020-10-28 | 24789.23 | 25578.07 | 5770.167 | 11986.84 | 26671.27 | 8604.793 | 12870.32 | 13827.12 | 1068.899 | 5656.529 |
| 2020-10-29 | 25080.13 | 25702.84 | 5805.08 | 12021.09 | 26945.95 | 8780.582 | 13362.16 | 14165.33 | 1069.442 | 5726.562 |
| 2020-10-30 | 25373.49 | 25816.65 | 5839.72 | 12052.93 | 27246.25 | 8978.618 | 13882.08 | 14523.21 | 1069.675 | 5799.463 |
| 2020-10-31 | 25587.17 | 25878.19 | 5873.424 | 12082.41 | 27512.58 | 9187.007 | 14273 | 14844.51 | 1070.062 | 5859.201 |
| 2020-11-01 | 25732.09 | 25925.8 | 5905.885 | 12105.24 | 27741.69 | 9376.646 | 14637.26 | 15185.44 | 1070.295 | 5914.428 |
| 2020-11-02 | 25942.54 | 25964.35 | 5933.378 | 12118.1 | 27990.54 | 9537.55 | 15249.37 | 15463.33 | 1070.644 | 5985.608 |
| 2020-11-03 | 26208.85 | 26032.49 | 5966.572 | 12138.77 | 28361.61 | 9747.273 | 15731.38 | 15757.32 | 1071.109 | 6056.01 |
| 2020-11-04 | 26442.41 | 26141.45 | 6002.606 | 12167.28 | 28663.85 | 9972.94 | 16073.83 | 16126.65 | 1071.42 | 6120.413 |
| 2020-11-05 | 26685.8 | 26252.88 | 6036.795 | 12198.36 | 29049.48 | 10206.18 | 16615.33 | 16480.72 | 1071.962 | 6199.084 |
| 2020-11-06 | 26944.24 | 26347.27 | 6072.933 | 12227.69 | 29424.84 | 10434.74 | 17171.69 | 16822.37 | 1072.273 | 6277.96 |
| 2020-11-07 | 27120.46 | 26430.19 | 6105.712 | 12256.52 | 29810.72 | 10688.57 | 17697.66 | 17188.27 | 1072.505 | 6353.336 |
| 2020-11-08 | 27237.36 | 26479.27 | 6138.655 | 12279.37 | 30164.86 | 10910.51 | 18060.85 | 17489.88 | 1072.777 | 6415.206 |
| 2020-11-09 | 27419.72 | 26541.95 | 6165.978 | 12300.14 | 30527.02 | 11115.44 | 18432.48 | 17803.33 | 1072.932 | 6479.221 |
| 2020-11-10 | 27682.34 | 26758.89 | 6197.757 | 12328.94 | 30933.36 | 11354.5 | 18842.79 | 18102.91 | 1073.009 | 6552.591 |
| 2020-11-11 | 27920.91 | 26881.54 | 6232.137 | 12364.58 | 31373.91 | 11604.78 | 19343.04 | 18439.59 | 1073.165 | 6632.14 |
| 2020-11-12 | 28165.68 | 27040.05 | 6264.345 | 12403.52 | 31852.38 | 11865.49 | 19812.42 | 18930.55 | 1073.203 | 6713.913 |
| 2020-11-13 | 28425.71 | 27220.98 | 6296.413 | 12440.38 | 32387.41 | 12142.78 | 20275.43 | 19330.81 | 1073.436 | 6797.688 |
| 2020-11-14 | 28611.39 | 27343.78 | 6325.909 | 12477.63 | 32883.78 | 12436.91 | 20655.33 | 19724.61 | 1074.561 | 6871.402 |
| 2020-11-15 | 28735.17 | 27408.76 | 6347.832 | 12508.31 | 33301.14 | 12688.6 | 20950.98 | 20090.59 | 1076.034 | 6931.887 |
| 2020-11-16 | 28908.24 | 27482.33 | 6368.762 | 12529.05 | 33770.78 | 12919.76 | 21309.02 | 20404.43 | 1076.383 | 6999.199 |
| 2020-11-17 | 29141.13 | 27648.18 | 6396.476 | 12562.14 | 34259.21 | 13199.91 | 21761.68 | 20698.4 | 1077.12 | 7076.709 |
| 2020-11-18 | 29367.68 | 27810.58 | 6429.184 | 12610.24 | 34776.48 | 13493.56 | 22183.24 | 20986.21 | 1077.43 | 7156.315 |
| 2020-11-19 | 29589.07 | 27978.16 | 6462.112 | 12652.11 | 35353.52 | 13807.13 | 22609.88 | 21322.34 | 1077.701 | 7239.442 |
| 2020-11-20 | 29799.75 | 28159.34 | 6495.291 | 12703.83 | 35957.13 | 14148.4 | 23033.46 | 21619.27 | 1078.244 | 7324.855 |
| 2020-11-21 | 29956.31 | 28300.56 | 6527.736 | 12747.89 | 36501.31 | 14490.36 | 23364.16 | 21910.87 | 1078.826 | 7399.481 |
| 2020-11-22 | 30048.05 | 28384.33 | 6559.355 | 12785.7 | 36940.02 | 14774.83 | 23600.95 | 22184.48 | 1079.33 | 7461.312 |
| 2020-11-23 | 30141.57 | 28469.19 | 6586.609 | 12820.34 | 37490.06 | 15033.12 | 23870.55 | 22410.99 | 1079.679 | 7529.335 |
| 2020-11-24 | 30298.65 | 28630.56 | 6618.456 | 12861.86 | 38008.64 | 15321.3 | 24227.99 | 22577.09 | 1080.067 | 7604.506 |
| 2020-11-25 | 30487.07 | 28839.11 | 6650.384 | 12915.99 | 38549.91 | 15647.15 | 24588.91 | 22844.45 | 1080.532 | 7684.639 |
| 2020-11-26 | 30685.36 | 29030.17 | 6681.302 | 12967.11 | 38900.16 | 16006.58 | 24946.53 | 23101.83 | 1080.842 | 7759.649 |
| 2020-11-27 | 30857.4 | 29186.75 | 6710.958 | 13023.24 | 39541.77 | 16386.04 | 25294.25 | 23318.96 | 1081.347 | 7847.624 |
| 2020-11-28 | 30991.11 | 29427.34 | 6740.963 | 13076.5 | 40053.4 | 16767.59 | 25562.64 | 23551.65 | 1081.579 | 7923.967 |
| 2020-11-29 | 31110.22 | 29525.53 | 6768.788 | 13119.18 | 40467.96 | 17072.91 | 25756.43 | 23729.86 | 1081.967 | 7985.342 |
| 2020-11-30 | 31235.77 | 29636.26 | 6791.121 | 13157.52 | 40940.73 | 17308.44 | 25970.9 | 23911.82 | 1082.355 | 8049.863 |
| 2020-12-01 | 31412 | 29879.19 | 6817.39 | 13195.75 | 41529.44 | 17602.32 | 26261.59 | 24109.32 | 1082.781 | 8128.972 |
| 2020-12-02 | 31577.17 | 30105.71 | 6842.904 | 13265.25 | 42121.56 | 17911.59 | 26568.38 | 24347.21 | 1083.402 | 8211.032 |
| 2020-12-03 | 31744.46 | 30340.01 | 6869.167 | 13338.53 | 42781.18 | 18252.36 | 26899.51 | 24566.24 | 1083.79 | 8298.875 |
| 2020-12-04 | 31895.73 | 30562.78 | 6895.471 | 13420.67 | 43486.08 | 18607.94 | 27206.45 | 24806.18 | 1084.061 | 8386.234 |
| 2020-12-05 | 32009.77 | 30751.02 | 6921.314 | 13498.04 | 44158.73 | 18933.64 | 27489.44 | 25034.34 | 1084.41 | 8468.572 |
| 2020-12-06 | 32081.65 | 30874.53 | 6944.984 | 13566.59 | 44694.28 | 19208.05 | 27697.09 | 25288.96 | 1084.681 | 8536.694 |
| 2020-12-07 | 32151.79 | 31000.82 | 6964.05 | 13621.77 | 45284.72 | 19414.76 | 27872.13 | 25505.2 | 1085.263 | 8603.806 |
| 2020-12-08 | 32230.95 | 31224.17 | 6987.073 | 13688.57 | 45959.13 | 19671.42 | 28166.12 | 25686.31 | 1085.496 | 8685.408 |
| 2020-12-09 | 32347.23 | 31479.27 | 7009.694 | 13800.31 | 46621.88 | 19969.05 | 28478.91 | 25929.92 | 1085.767 | 8770.824 |
| 2020-12-10 | 32500.59 | 31729.52 | 7030.774 | 13936.31 | 47325.21 | 20285.06 | 28829.79 | 26238.13 | 1086.194 | 8961.714 |
| 2020-12-11 | 32656.53 | 31975.08 | 7052.326 | 14074.87 | 48047.49 | 20604.68 | 29167.29 | 26557.07 | 1086.737 | 9051.48 |
| 2020-12-12 | 32772.18 | 32172.87 | 7074.038 | 14206.14 | 48715.72 | 20908.8 | 29454.31 | 26873.05 | 1086.931 | 9132.814 |
| 2020-12-13 | 32850.19 | 32275.81 | 7093.466 | 14339.36 | 49291.98 | 21129.77 | 29684.96 | 27144.84 | 1087.202 | 9200.812 |
| 2020-12-14 | 32961.19 | 32403.76 | 7109.302 | 14425.35 | 49928.94 | 21287.98 | 29872.68 | 27442.86 | 1087.629 | 9270.069 |
| 2020-12-15 | 33114.26 | 32628.21 | 7128.235 | 14551.13 | 50599.38 | 21491.36 | 30198.32 | 27714.24 | 1088.094 | 9353.726 |
| 2020-12-16 | 33264.31 | 32937.21 | 7145.466 | 14717.82 | 51310.7 | 21745.74 | 30566.98 | 28084.59 | 1088.559 | 9445.615 |
| 2020-12-17 | 33424.94 | 33258.24 | 7161.893 | 14869.81 | 52028.55 | 22032.71 | 30936.91 | 28604.77 | 1089.373 | 9539.857 |
| 2020-12-18 | 33578.48 | 33505.23 | 7179.944 | 15015.12 | 52780.99 | 22333.31 | 31298.2 | 29022.72 | 1090.653 | 9631.722 |
| 2020-12-19 | 33705.54 | 33730.67 | 7199.051 | 15197.31 | 53378.12 | 22613.5 | 31583.8 | 29421.35 | 1092.282 | 9710.294 |
| 2020-12-20 | 33795.79 | 33845.08 | 7216.517 | 15354.62 | 53959.11 | 22815.1 | 31785.17 | 29949.6 | 1093.445 | 9778.797 |
| 2020-12-21 | 33924.13 | 33974.27 | 7230.552 | 15501 | 54562.65 | 22975.64 | 32007.03 | 30440.36 | 1094.259 | 9849.159 |
| 2020-12-22 | 34102.64 | 34239.06 | 7247.74 | 15659.24 | 55173.76 | 23181.52 | 32354.57 | 30980.49 | 1094.957 | 9933.6 |
| 2020-12-23 | 34290.91 | 34452.52 | 7265.475 | 15893.18 | 55842.16 | 23424.74 | 32712.99 | 31556.94 | 1095.927 | 10021.47 |
| 2020-12-24 | 34290.91 | 34724.55 | 7282.029 | 16131.43 | 56477.54 | 23699.11 | 33068.41 | 32129.6 | 1096.392 | 10108.91 |
| 2020-12-25 | 34525.28 | 34823.54 | 7298.014 | 16377.85 | 56840.11 | 23962.71 | 33290.2 | 32609.38 | 1097.284 | 10171 |
| 2020-12-26 | 34606.7 | 34905.47 | 7311.457 | 16570.25 | 57521.93 | 24149.85 | 33423.86 | 33118.03 | 1097.711 | 10236.52 |
| 2020-12-27 | 34716.99 | 34987.78 | 7325.825 | 16728.51 | 57929.77 | 24300.49 | 33561.34 | 33593.34 | 1098.835 | 10288.97 |
| 2020-12-28 | 34875.22 | 35115.99 | 7337.618 | 16852.72 | 58451.3 | 24411.12 | 33744.29 | 34200.52 | 1099.301 | 10352.71 |
| 2020-12-29 | 35130.67 | 35379.42 | 7352.365 | 17012.28 | 59059.84 | 24581.42 | 34080.04 | 34979.77 | 1100.541 | 10437.9 |
| 2020-12-30 | 35388.64 | 35639.24 | 7368.026 | 17307.24 | 59720.4 | 24774.76 | 34496.79 | 35713.46 | 1101.472 | 10531.58 |
| 2020-12-31 | 35642.69 | 35893.77 | 7382.404 | 17607.03 | 60657.64 | 25007.51 | 34885.88 | 36533.4 | 1102.248 | 10636.85 |
| 2021-01-01 | 35732.15 | 36000.97 | 7396.097 | 17885.6 | 61175.46 | 25234.3 | 35179.81 | 37314.64 | 1103.605 | 10709.38 |
| 2021-01-02 | 35847.05 | 36072.71 | 7409.142 | 18135.46 | 62023.71 | 25358.56 | 35324.58 | 38160.95 | 1104.535 | 10784.5 |
| 2021-01-03 | 35976.06 | 36153.04 | 7420.986 | 18332.97 | 62636.53 | 25470.85 | 35499.56 | 38968.08 | 1105.311 | 10850.88 |
| 2021-01-04 | 36156.35 | 36270.05 | 7432.738 | 18542.84 | 63193.82 | 25574.24 | 35725.95 | 39830.23 | 1105.815 | 10921.92 |
| 2021-01-05 | 36458.72 | 36541.48 | 7445.719 | 18782.84 | 63884.18 | 25703.65 | 36128.01 | 40723.66 | 1106.552 | 11016.39 |
| 2021-01-06 | 36753.44 | 36833.57 | 7460.321 | 19146.45 | 64665.84 | 25869.89 | 36504.54 | 41637.81 | 1106.94 | 11117.19 |
| 2021-01-07 | 37056.8 | 37244.66 | 7473.338 | 19496.19 | 65530.55 | 26084.31 | 37008.93 | 42409.49 | 1107.909 | 11230.49 |
| 2021-01-08 | 37349.44 | 37523.1 | 7473.338 | 19862.26 | 66445.2 | 26222.3 | 37390.97 | 43407.42 | 1108.336 | 11335.32 |
| 2021-01-09 | 37591.89 | 37763.27 | 7499.796 | 20222.11 | 67236.54 | 26340.62 | 37700.94 | 44286.72 | 1108.84 | 11431.3 |
| 2021-01-10 | 37763.09 | 37898.91 | 7511.502 | 20512.26 | 67876.18 | 26463.06 | 37910.05 | 45092.45 | 1109.577 | 11506.15 |
| 2021-01-11 | 37953.95 | 38042.59 | 7520.533 | 20762.85 | 68487.85 | 26568.54 | 38195.14 | 45769.67 | 1110.352 | 11584.16 |
| 2021-01-12 | 38256.17 | 38343.65 | 7531.993 | 20981.12 | 69150.16 | 26692.61 | 38544.24 | 46437.4 | 1110.973 | 11673.45 |
| 2021-01-13 | 38535.19 | 38623.22 | 7544.154 | 21290.15 | 69840.4 | 26847.1 | 38940.91 | 47134.38 | 1111.36 | 11768.72 |
| 2021-01-14 | 38826.51 | 38942.53 | 7555.343 | 21598.32 | 70544.67 | 27040 | 39317.82 | 47848.31 | 1111.709 | 11864.83 |
| 2021-01-15 | 39096.92 | 39265.42 | 7566.221 | 21846.14 | 71289.38 | 27238.32 | 39653.18 | 48666.07 | 1112.485 | 11963.15 |
| 2021-01-16 | 39292.77 | 39538.53 | 7577.09 | 22078.86 | 71936.83 | 27425.84 | 39896.21 | 49272.24 | 1113.222 | 12045.9 |
| 2021-01-17 | 39452.05 | 39685.34 | 7586.985 | 22283.17 | 72473.12 | 27573.03 | 40082.62 | 49838.23 | 1113.726 | 12113.12 |
| 2021-01-18 | 39631.52 | 39820.55 | 7594.197 | 22433.23 | 72898.4 | 27650.84 | 40388.47 | 50388.64 | 1114.113 | 12179.15 |
| 2021-01-19 | 39897.73 | 40110.97 | 7604.112 | 22596.12 | 73362.43 | 27748.18 | 40698.52 | 50877.9 | 1114.462 | 12254.07 |
| 2021-01-20 | 40163.31 | 40412.28 | 7615.053 | 22807.8 | 73928.76 | 27856.97 | 41094.58 | 51448.48 | 1114.811 | 12343.08 |
| 2021-01-21 | 40413.2 | 40687.75 | 7625.491 | 22997.35 | 74503.55 | 27993.31 | 41435.4 | 52004.15 | 1115.044 | 12426.44 |
| 2021-01-22 | 40648.98 | 40952.27 | 7635.722 | 23193.23 | 75084.02 | 28123.96 | 41787.09 | 52594.53 | 1115.238 | 12510.82 |
| 2021-01-23 | 40832.33 | 41224.7 | 7646.379 | 23397.61 | 75618.22 | 28245.06 | 42026.51 | 53086.68 | 1115.471 | 12583.71 |
| 2021-01-24 | 40942.65 | 41357.97 | 7655.854 | 23533.3 | 76043.06 | 28342.65 | 42218.57 | 53527.06 | 1115.897 | 12642.44 |
| 2021-01-25 | 41108.81 | 41500.64 | 7662.386 | 23609.09 | 76431.83 | 28407.23 | 42540.42 | 53852.8 | 1116.014 | 12703.56 |

**Table 2.** Accumulated number of laboratory-confirmed COVID-19 cases per million in different regions for the period August 25, 2021 – January 20, 2022. [5].

|  | Argentina | Brazil | India | South Africa | United States | Ukraine | European Union | United Kingdom | Australia | World |
| --- | --- | --- | --- | --- | --- | --- | --- | --- | --- | --- |
| 2021-08-25 | 113035.5 | 96510.72 | 23366.1 | 45338.3 | 115383.4 | 54567.94 | 81364.06 | 96780.89 | 1855.111 | 27238.28 |
| 2021-08-26 | 113185.7 | 96655.22 | 23398.15 | 45551 | 115947.2 | 54616.65 | 81520.01 | 97338.04 | 1892.958 | 27332.1 |
| 2021-08-27 | 113313 | 96784.92 | 23431.7 | 45751.61 | 116663.6 | 54675.52 | 81615.48 | 97891.33 | 1936.349 | 27426.2 |
| 2021-08-28 | 113394.7 | 96892.16 | 23464.06 | 45921.04 | 116882.5 | 54735.06 | 81769.74 | 98361.76 | 1987.574 | 27496.97 |
| 2021-08-29 | 113440.1 | 96944.79 | 23494.85 | 46049.95 | 117019.6 | 54790.13 | 81859.27 | 98844.66 | 2040.118 | 27553.85 |
| 2021-08-30 | 113557.6 | 97004.61 | 23517.06 | 46143.95 | 117739.1 | 54818.11 | 81978.8 | 99229.18 | 2088.861 | 27638.82 |
| 2021-08-31 | 113705.2 | 97130.06 | 23547.17 | 46261.94 | 118280.6 | 54859.41 | 82106.18 | 99699.12 | 2136.363 | 27718.88 |
| 2021-09-01 | 113822 | 97250.38 | 23580.97 | 46420.89 | 118871.4 | 54916.64 | 82296.2 | 100220.1 | 2193.25 | 27811.04 |
| 2021-09-02 | 113924.1 | 97376.66 | 23613.52 | 46574.15 | 119399.7 | 54982.7 | 82441.89 | 100775.3 | 2257.155 | 27896.49 |
| 2021-09-03 | 114018.8 | 97486.97 | 23644.1 | 46727.36 | 120112.5 | 55053.53 | 82563.05 | 101392.8 | 2324.666 | 27987.65 |
| 2021-09-04 | 114073.3 | 97581.94 | 23674.8 | 46867.43 | 120344.3 | 55113.67 | 82666.3 | 101930 | 2389.425 | 28050 |
| 2021-09-05 | 114103.9 | 97641.11 | 23702.75 | 46966.21 | 120501.2 | 55162.95 | 82751.7 | 102465.3 | 2448.987 | 28106.91 |
| 2021-09-06 | 114189.3 | 97688.64 | 23725.15 | 47034.8 | 120743.1 | 55189.22 | 82838.14 | 103065.5 | 2505.834 | 28163.29 |
| 2021-09-07 | 114279.3 | 97752.22 | 23752.34 | 47130.91 | 121470.8 | 55248.16 | 82991.4 | 103611.3 | 2571.601 | 28252.39 |
| 2021-09-08 | 114356.7 | 97818.49 | 23783.38 | 47246.48 | 122005.3 | 55320.01 | 83101.22 | 104175.9 | 2638.492 | 28332.01 |
| 2021-09-09 | 114437 | 97971.87 | 23808.48 | 47350.89 | 122502.2 | 55412.26 | 83271.92 | 104726 | 2711.006 | 28413.9 |
| 2021-09-10 | 114498.7 | 98048.77 | 23832.44 | 47448.87 | 123200 | 55503.09 | 83379.92 | 105265.2 | 2791.469 | 28494.99 |
| 2021-09-11 | 114538.1 | 98113.08 | 23852.95 | 47537.29 | 123434.7 | 55599.63 | 83475.44 | 105688.3 | 2854.133 | 28554.03 |
| 2021-09-12 | 114558.5 | 98149.81 | 23872.51 | 47603.26 | 123569.6 | 55657.49 | 83549.68 | 106107.8 | 2920.83 | 28601.1 |
| 2021-09-13 | 114608.9 | 98190.22 | 23890.74 | 47647.23 | 124276.2 | 55692.5 | 83638.01 | 106553.3 | 2982.021 | 28675.56 |
| 2021-09-14 | 114675 | 98253.64 | 23910.25 | 47708.84 | 124704.8 | 55776.24 | 83768.79 | 106938.7 | 3045.732 | 28745.16 |
| 2021-09-15 | 114730 | 98315.68 | 23932.19 | 47786.57 | 125234.5 | 55889.85 | 83878.46 | 107376.7 | 3117.703 | 28817.82 |
| 2021-09-16 | 114784.7 | 98481.33 | 23956.88 | 47856.75 | 125705.9 | 56029.03 | 84021.44 | 107763.4 | 3187.502 | 28891.35 |
| 2021-09-17 | 114835.3 | 98646.46 | 23982.47 | 47917.51 | 126338.6 | 56188.7 | 84139.47 | 108237.5 | 3259.473 | 28970 |
| 2021-09-18 | 114867.1 | 99230.02 | 24004.55 | 47972.24 | 126552.8 | 56339.71 | 84233.74 | 108669.1 | 3320.431 | 29036.37 |
| 2021-09-19 | 114880.8 | 99272.89 | 24026.27 | 48010.23 | 126682.5 | 56439.37 | 84304.25 | 109094.4 | 3378.597 | 29082.62 |
| 2021-09-20 | 114928.2 | 99316.28 | 24045.01 | 48035.28 | 127221 | 56499.94 | 84386.06 | 109618.8 | 3439.943 | 29148.64 |
| 2021-09-21 | 114968.5 | 99388.15 | 24064.36 | 48071.87 | 127572.6 | 56627.44 | 84500.88 | 110075.2 | 3504.275 | 29209.66 |
| 2021-09-22 | 115013.1 | 99540.42 | 24087.27 | 48121.29 | 127983 | 56791.98 | 84623.42 | 110567.6 | 3574.423 | 29277.95 |
| 2021-09-23 | 115051.1 | 99656.85 | 24109.79 | 48167.64 | 128366.2 | 56982.5 | 84743.21 | 111092.4 | 3642.826 | 29346.2 |
| 2021-09-24 | 115091.6 | 99744.08 | 24131.05 | 48205.29 | 128896.5 | 57200.94 | 84856.31 | 111615.2 | 3715.146 | 29415.59 |
| 2021-09-25 | 115113.4 | 99808.86 | 24151.38 | 48232.51 | 129073.7 | 57401.67 | 84946.09 | 112051.3 | 3783.084 | 29463.01 |
| 2021-09-26 | 115125.7 | 99850.24 | 24170.06 | 48248.61 | 129230.7 | 57519.69 | 85020.1 | 112535.1 | 3840.165 | 29507.8 |
| 2021-09-27 | 115159.4 | 99893.91 | 24183.55 | 48258.24 | 129710.6 | 57600.48 | 85103.33 | 113085.1 | 3913.066 | 29565.95 |
| 2021-09-28 | 115199.4 | 99972.81 | 24197.1 | 48281.01 | 130049.2 | 57763.16 | 85226.12 | 113592.4 | 3983.331 | 29623.39 |
| 2021-09-29 | 115232.2 | 100051.7 | 24213.98 | 48281.01 | 130412.6 | 57997.94 | 85364.39 | 114106.9 | 4076.397 | 29686.84 |
| 2021-09-30 | 115268.2 | 100154.3 | 24233.16 | 48344.03 | 130747 | 58281.26 | 85498.6 | 114631.3 | 4156.2 | 29747.86 |
| 2021-10-01 | 115302.5 | 100238.7 | 24250.64 | 48371.26 | 131211.4 | 58571.2 | 85623.33 | 115138.8 | 4246.746 | 29813.41 |
| 2021-10-02 | 115321.9 | 100296.6 | 24267.03 | 48393.01 | 131352.6 | 58856.39 | 85727.41 | 115569.6 | 4319.337 | 29857.86 |
| 2021-10-03 | 115330.4 | 100339.4 | 24281.96 | 48406.49 | 131483.6 | 59053.66 | 85806.99 | 116004.6 | 4397.784 | 29897.17 |
| 2021-10-04 | 115351.9 | 100393.2 | 24295.13 | 48413.63 | 131920.6 | 59179.09 | 85899.42 | 116507.5 | 4490.423 | 29952.55 |
| 2021-10-05 | 115378.6 | 100496.7 | 24308.64 | 48426.42 | 132207 | 59420.45 | 86029.04 | 116992.5 | 4568.986 | 30005.86 |
| 2021-10-06 | 115406.7 | 100583.3 | 24324.74 | 48445.56 | 132546.4 | 59726.8 | 86179.31 | 117560.1 | 4654.917 | 30069.96 |
| 2021-10-07 | 115430.5 | 100651.6 | 24340 | 48462.03 | 132851.2 | 60090.15 | 86325.36 | 118143.1 | 4752.791 | 30126.34 |
| 2021-10-08 | 115447.1 | 100738.2 | 24354.16 | 48477.42 | 133246.9 | 60482.34 | 86467.37 | 118655.4 | 4850.316 | 30187.31 |
| 2021-10-09 | 115457.4 | 100807.4 | 24367.2 | 48491.01 | 133360.1 | 60864.74 | 86584.66 | 119233.3 | 4942.994 | 30230.21 |
| 2021-10-10 | 115464.6 | 100846.7 | 24380.21 | 48501.8 | 133477 | 61143.16 | 86674.11 | 119725.8 | 5024.271 | 30270.52 |
| 2021-10-11 | 115473.7 | 100879.4 | 24390.48 | 48505.15 | 133769.8 | 61364.14 | 86780.88 | 120306.2 | 5095.932 | 30318.16 |
| 2021-10-12 | 115497.1 | 100913.8 | 24401.84 | 48515.01 | 134103.6 | 61657.97 | 86930.76 | 120859.2 | 5174.611 | 30373.78 |
| 2021-10-13 | 115525.9 | 100952.9 | 24415.47 | 48530.7 | 134430.6 | 62051.37 | 87095.46 | 121467.6 | 5281.017 | 30431.06 |
| 2021-10-14 | 115555.5 | 101024.1 | 24427.57 | 48546.47 | 134691.9 | 62504.39 | 87267.02 | 122121.8 | 5379.201 | 30487.71 |
| 2021-10-15 | 115585.3 | 101090.9 | 24439.04 | 48558.68 | 135021.8 | 62837.01 | 87438.26 | 122770.6 | 5468.815 | 30545.64 |
| 2021-10-16 | 115602.6 | 101136.5 | 24449.19 | 48568.99 | 135139.7 | 63155.41 | 87579.53 | 123398.3 | 5553.079 | 30588.87 |
| 2021-10-17 | 115611.4 | 101165.4 | 24458.95 | 48575.88 | 135233.1 | 63435.23 | 87689.15 | 124053.6 | 5632.921 | 30628.71 |
| 2021-10-18 | 115631.4 | 101209 | 24468.32 | 48579.38 | 135533.5 | 63674.84 | 87827.76 | 124769.9 | 5711.95 | 30681.02 |
| 2021-10-19 | 115659.9 | 101270.8 | 24478.81 | 48586.91 | 135777.7 | 64054.07 | 88043.82 | 125406.3 | 5793.266 | 30737.23 |
| 2021-10-20 | 115686.6 | 101344.2 | 24492.06 | 48596.75 | 136053.9 | 64510.12 | 88276.78 | 126118.9 | 5891.916 | 30797.14 |
| 2021-10-21 | 115720.4 | 101423.6 | 24503.38 | 48605.41 | 136288 | 65047.04 | 88505.87 | 126874.6 | 5990.682 | 30855.07 |
| 2021-10-22 | 115750.8 | 101486.2 | 24515.1 | 48614.02 | 136577.5 | 65615.77 | 88736.28 | 127590 | 6063.7 | 30916.75 |
| 2021-10-23 | 115770.7 | 101541.8 | 24526.52 | 48621.5 | 136673.1 | 66172.03 | 88911.56 | 128249.5 | 6148.002 | 30962.91 |
| 2021-10-24 | 115782.5 | 101563 | 24536.78 | 48626.5 | 136728.9 | 66672.67 | 89066.29 | 128817.5 | 6213.303 | 31002.98 |
| 2021-10-25 | 115809.4 | 101599.6 | 24545.7 | 48628.93 | 137062.7 | 67031.95 | 89256.5 | 129349.4 | 6282.559 | 31057.3 |
| 2021-10-26 | 115840.5 | 101662 | 24555.36 | 48634.44 | 137270.7 | 67494.65 | 89531.04 | 129987.2 | 6354.297 | 31113.58 |
| 2021-10-27 | 115873 | 101742 | 24566.95 | 48642.3 | 137578.2 | 68037.16 | 89832.51 | 130632.3 | 6433.326 | 31178.84 |
| 2021-10-28 | 115907.9 | 101805.9 | 24577.25 | 48651.18 | 137805.8 | 68660.55 | 90079.8 | 131205.4 | 6506.46 | 31236.71 |
| 2021-10-29 | 115938 | 101865.2 | 24587.52 | 48659.09 | 138091.5 | 69302.74 | 90394.59 | 131838.2 | 6568.077 | 31301.1 |
| 2021-10-30 | 115955.8 | 101912 | 24596.73 | 48664.04 | 138193.3 | 69929.57 | 90632.22 | 132435.3 | 6614.028 | 31349.8 |
| 2021-10-31 | 115967.8 | 101940.6 | 24605.71 | 48667.87 | 138289.5 | 70354.99 | 90785.62 | 132987.6 | 6670.876 | 31390.6 |
| 2021-11-01 | 115992.8 | 101962.2 | 24613.19 | 48669.64 | 138624.3 | 70700.48 | 90968.48 | 133573.1 | 6714.888 | 31443.89 |
| 2021-11-02 | 116022.1 | 101989.9 | 24621.73 | 48672.45 | 138837.5 | 71172.93 | 91222.44 | 134065.9 | 6757.97 | 31497.38 |
| 2021-11-03 | 116049.9 | 102066.1 | 24630.98 | 48678.18 | 139093.9 | 71736.09 | 91618.25 | 134664.1 | 6817.571 | 31564.85 |
| 2021-11-04 | 116081.4 | 102128 | 24640.11 | 48683.49 | 139339.5 | 72391.24 | 92005.88 | 135202.8 | 6878.839 | 31630.81 |
| 2021-11-05 | 116109.3 | 102188.8 | 24647.96 | 48689.14 | 139629.9 | 73026.34 | 92362.62 | 135698.5 | 6938.324 | 31696.24 |
| 2021-11-06 | 116129.6 | 102241 | 24655.72 | 48695.1 | 139733.9 | 73628.82 | 92693.52 | 136140.5 | 6992.108 | 31750.08 |
| 2021-11-07 | 116142.6 | 102267.4 | 24663.96 | 48698.51 | 139842.9 | 74055.73 | 92918.84 | 136578 | 7041.123 | 31793.98 |
| 2021-11-08 | 116170.9 | 102299.6 | 24671.23 | 48700.45 | 140170.5 | 74382.66 | 93243.89 | 137051.1 | 7091.223 | 31854.48 |
| 2021-11-09 | 116200.5 | 102354.8 | 24679.46 | 48704.53 | 140410.7 | 74846.21 | 93605.87 | 137531.8 | 7137.795 | 31915.89 |
| 2021-11-10 | 116234.8 | 102424.8 | 24688.85 | 48709.61 | 140698.2 | 75408.6 | 94104.55 | 138111.6 | 7198.132 | 31989.27 |
| 2021-11-11 | 116266.8 | 102497.4 | 24697.84 | 48715.54 | 140868.4 | 76004.96 | 94537.92 | 138736 | 7252.964 | 32054.98 |
| 2021-11-12 | 116302.2 | 102565.6 | 24706.34 | 48722.08 | 141303.5 | 76585.63 | 94974.87 | 139308.4 | 7309.268 | 32130.28 |
| 2021-11-13 | 116326.2 | 102609.3 | 24714.43 | 48727.18 | 141434.2 | 77155.26 | 95330.29 | 139860.6 | 7352.505 | 32184.37 |
| 2021-11-14 | 116339.1 | 102629.4 | 24721.77 | 48731.54 | 141528 | 77516.3 | 95597.9 | 140390.3 | 7391.089 | 32229.17 |
| 2021-11-15 | 116370.2 | 102642.3 | 24728.13 | 48733.81 | 141932.9 | 77792.64 | 96010.93 | 140973.8 | 7430.409 | 32297.82 |
| 2021-11-16 | 116405.8 | 102672.9 | 24735.45 | 48738.35 | 142194.5 | 78195.57 | 96491.03 | 141515.6 | 7478.028 | 32364.23 |
| 2021-11-17 | 116439.8 | 102734.2 | 24744 | 48747.78 | 142533 | 78652.91 | 97094.02 | 142073.8 | 7527.159 | 32443.89 |
| 2021-11-18 | 116478.3 | 102787.5 | 24751.97 | 48757.52 | 142864 | 79154.67 | 97659.17 | 142762.7 | 7585.519 | 32521.83 |
| 2021-11-19 | 116511.6 | 102845.3 | 24759.37 | 48770.66 | 143245.3 | 79644.03 | 98211.91 | 143413.9 | 7638.256 | 32600.03 |
| 2021-11-20 | 116535.6 | 102883.7 | 24766.89 | 48785.44 | 143379.8 | 80092.05 | 98683.1 | 143998.6 | 7695.065 | 32660.92 |
| 2021-11-21 | 116549.8 | 102907.5 | 24772.99 | 48796.88 | 143497.7 | 80364.81 | 99039.83 | 144577.1 | 7741.869 | 32711.43 |
| 2021-11-22 | 116563.8 | 102926.8 | 24778.42 | 48802.08 | 143935.2 | 80562.94 | 99611.32 | 145237.1 | 7780.724 | 32790.35 |
| 2021-11-23 | 116599.9 | 102985 | 24785.09 | 49111.63 | 144209.5 | 80881.02 | 100150.3 | 145865.1 | 7837.494 | 32867.63 |
| 2021-11-24 | 116648.9 | 103042.2 | 24791.63 | 49132.86 | 144544.5 | 81233.66 | 100858 | 146490 | 7896.707 | 32952.87 |
| 2021-11-25 | 116698.4 | 103099.6 | 24799.2 | 49173.92 | 144661.8 | 81644.85 | 101526.6 | 147178.4 | 7959.798 | 33028.56 |
| 2021-11-26 | 116740.3 | 103150.5 | 24805.17 | 49221.02 | 144820.2 | 82031.39 | 102189.3 | 147904.9 | 8017.189 | 33104.19 |
| 2021-11-27 | 116773.7 | 103189.3 | 24811.47 | 49274.64 | 144898.3 | 82377.11 | 102679.5 | 148485 | 8065.079 | 33162.26 |
| 2021-11-28 | 116793.2 | 103205.8 | 24817.43 | 49322.24 | 145026 | 82566.79 | 103073.2 | 149020.2 | 8109.867 | 33213.95 |
| 2021-11-29 | 116836.3 | 103226.8 | 24822.45 | 49360.1 | 145589.9 | 82716.19 | 103628.2 | 149646.3 | 8152.483 | 33297.5 |
| 2021-11-30 | 116887.4 | 103274.9 | 24828.87 | 49432.93 | 145942.4 | 82974.14 | 104256.2 | 150232.9 | 8207.431 | 33376.93 |
| 2021-12-01 | 116928.7 | 103332.3 | 24835.88 | 49575.52 | 146353.9 | 83264.04 | 104959.5 | 150928.7 | 8273.43 | 33466.6 |
| 2021-12-02 | 116987.5 | 103388 | 24842.5 | 49767.63 | 146772.1 | 83589.87 | 105661.8 | 151710.4 | 8332.449 | 33556.26 |
| 2021-12-03 | 117039.7 | 103437.6 | 24848.67 | 50035.03 | 147248.6 | 83920.95 | 106331.6 | 152455.4 | 8398.681 | 33646.94 |
| 2021-12-04 | 117076.8 | 103472.3 | 24848.67 | 50307.61 | 147445.7 | 84238.57 | 106845.5 | 153063.2 | 8447.114 | 33711.69 |
| 2021-12-05 | 117105.1 | 103494.2 | 24861.01 | 50492.89 | 147608.7 | 84404.47 | 107234.9 | 153697.8 | 8496.943 | 33767.47 |
| 2021-12-06 | 117159.4 | 103522.3 | 24865.91 | 50599.17 | 148147.8 | 84520.53 | 107640.6 | 154451.8 | 8552.549 | 33842.76 |
| 2021-12-07 | 117227.2 | 103569.7 | 24871.97 | 50818.13 | 148488 | 84732.31 | 108397 | 155116.5 | 8618.665 | 33931.14 |
| 2021-12-08 | 117268.4 | 103617 | 24878.73 | 51148.6 | 148937.5 | 84959.95 | 108968.7 | 155861.4 | 8682.803 | 34015.89 |
| 2021-12-09 | 117328.6 | 103657.9 | 24884.76 | 51521.47 | 149305.3 | 85256.36 | 109725.1 | 156598.6 | 8749.151 | 34108.35 |
| 2021-12-10 | 117406.9 | 103690.9 | 24890.49 | 51838.2 | 149788 | 85528.22 | 110335.2 | 157454.1 | 8817.283 | 34195.62 |
| 2021-12-11 | 117460.5 | 103703.5 | 24896.14 | 52123.88 | 149955.7 | 85772.22 | 110789.3 | 158220.6 | 8877.116 | 34258.67 |
| 2021-12-12 | 117495 | 103711 | 24901.42 | 52754.69 | 150109.8 | 85903.84 | 111146.2 | 158925.4 | 8948.583 | 34314.64 |
| 2021-12-13 | 117572 | 103724 | 24905.57 | 52976 | 150671.8 | 86005.6 | 111636.9 | 159716.4 | 9024.974 | 34394.79 |
| 2021-12-14 | 117671.9 | 103744.4 | 24910.58 | 53373.34 | 151016.7 | 86183.11 | 112248.3 | 160592.4 | 9133.9 | 34478.23 |
| 2021-12-15 | 117777.5 | 103768.2 | 24916.3 | 53812.85 | 151448 | 86377.77 | 112917.9 | 161735.4 | 9266.635 | 34571.97 |
| 2021-12-16 | 117893.8 | 103785.3 | 24921.65 | 54225.65 | 151873.3 | 86605.94 | 113535.2 | 163022.7 | 9414.067 | 34665.66 |
| 2021-12-17 | 118017.6 | 103805.7 | 24921.65 | 54570.62 | 152452.3 | 86817.94 | 114086.1 | 164382 | 9569.836 | 34758.62 |
| 2021-12-18 | 118108.9 | 103816.8 | 24926.78 | 54838.43 | 152686 | 86997.53 | 114617.6 | 165687.9 | 9719.594 | 34831.88 |
| 2021-12-19 | 118180.2 | 103823.6 | 24936.57 | 55096 | 152949.7 | 87087 | 114932.4 | 166889.5 | 9875.906 | 34892.69 |
| 2021-12-20 | 118297.3 | 103835.9 | 24940.39 | 55237.75 | 153677.9 | 87151.74 | 115431.9 | 168243.8 | 10050.09 | 34988.31 |
| 2021-12-21 | 118502 | 103853.4 | 24944.92 | 55494.62 | 154213.7 | 87296.24 | 116138.9 | 169553 | 10264.57 | 35088.64 |
| 2021-12-22 | 118745.8 | 103868.5 | 24950.3 | 55846.01 | 154936.8 | 87447.82 | 116866.6 | 171101.6 | 10588.63 | 35203.06 |
| 2021-12-23 | 119040.9 | 103886.1 | 24955.07 | 56198.37 | 155747 | 87621.13 | 117657.1 | 172901.5 | 10907.31 | 35328.95 |
| 2021-12-24 | 119397.8 | 103903.3 | 24960.23 | 56512.26 | 156448.3 | 87778.21 | 118215.8 | 174687.3 | 11289.46 | 35436.18 |
| 2021-12-25 | 119555.3 | 103920.5 | 24965.25 | 56759.22 | 156667 | 87903.62 | 118842.2 | 176474 | 11656.25 | 35521.41 |
| 2021-12-26 | 119722.5 | 103943.7 | 24969.94 | 56852.54 | 157254.8 | 87972.36 | 119082.5 | 178217.8 | 12039.53 | 35595.63 |
| 2021-12-27 | 120166.8 | 103976.2 | 24974.5 | 56915.46 | 158782.6 | 88019.25 | 120008.9 | 179796.8 | 12496.99 | 35758.42 |
| 2021-12-28 | 120910.2 | 104018.5 | 24981.1 | 57035.65 | 159851.9 | 88074.9 | 121205.7 | 181820.5 | 13182.49 | 35924.41 |
| 2021-12-29 | 121831.8 | 104064.1 | 24990.54 | 57185.87 | 161348.9 | 88204.1 | 122708 | 184525.1 | 14011.13 | 36145.88 |
| 2021-12-30 | 122939.2 | 104123 | 25002.57 | 57402.02 | 163116 | 88344.19 | 124379.7 | 187295 | 15322.23 | 36391.52 |
| 2021-12-31 | 123984.3 | 104170.7 | 25018.91 | 57597.79 | 164587.3 | 88509.48 | 125735.3 | 190088.6 | 16499.63 | 36608.88 |
| 2022-01-01 | 124423.3 | 104188.3 | 25038.69 | 57760.89 | 165086.8 | 88628.45 | 126889.7 | 192472.1 | 17952.19 | 36755.39 |
| 2022-01-02 | 124872.9 | 104196.8 | 25062.91 | 57833.45 | 166000.3 | 88674.65 | 127354.9 | 194450.5 | 19100.31 | 36872.88 |
| 2022-01-03 | 125846.3 | 104251.2 | 25089.73 | 57884.69 | 169233.7 | 88719.6 | 129015.3 | 197198.2 | 20835.41 | 37181.37 |
| 2022-01-04 | 127627 | 104340.8 | 25131.43 | 58019.22 | 171653.4 | 88763.06 | 131020.2 | 200441.6 | 23607.8 | 37505.14 |
| 2022-01-05 | 129713.6 | 104447.6 | 25196.68 | 58204.19 | 173589.2 | 88871.44 | 133295.8 | 203294.9 | 26532.62 | 37829.87 |
| 2022-01-06 | 132117 | 104615 | 25280.72 | 58368.38 | 176036.2 | 89027.17 | 135091.5 | 205942.9 | 29566.14 | 38162.66 |
| 2022-01-07 | 134540.6 | 104910.8 | 25382.62 | 58522.59 | 178609.8 | 89195.2 | 137375 | 208565.3 | 33369.89 | 38535.24 |
| 2022-01-08 | 136770.4 | 105141.2 | 25497.18 | 58651.81 | 179768.5 | 89272.02 | 139020.7 | 210626.2 | 37709.35 | 38804.52 |
| 2022-01-09 | 138378 | 105279.8 | 25626.16 | 58726.46 | 181197.5 | 89339.77 | 140464 | 212699.2 | 40533.67 | 39059.36 |
| 2022-01-10 | 140315.3 | 105438.3 | 25746.78 | 58766.58 | 185307.8 | 89388.09 | 142149.1 | 214808.5 | 44171.22 | 39463.87 |
| 2022-01-11 | 143263.2 | 105780.6 | 25746.78 | 58860.99 | 187618.4 | 89515.95 | 144798.8 | 216506.5 | 47444.34 | 39828.93 |
| 2022-01-12 | 146137.4 | 106191.3 | 26064.08 | 58973.57 | 190350.5 | 89683.12 | 147384 | 218412.2 | 54240.9 | 40300.24 |
| 2022-01-13 | 148952.9 | 106649 | 26064.08 | 59072.12 | 192874 | 89918.44 | 149643.3 | 219967.5 | 59211.58 | 40666.37 |
| 2022-01-14 | 152019.4 | 107168.2 | 26446.62 | 59159.31 | 195454.1 | 90164.2 | 152118.6 | 221418.2 | 62557.6 | 41120.78 |
| 2022-01-15 | 154138.7 | 107395.1 | 26641.25 | 59235.76 | 196636.3 | 90412.64 | 153866.6 | 222601.8 | 66934.29 | 41426.84 |
| 2022-01-16 | 155569.3 | 107550.6 | 26826.48 | 59280.04 | 198040.2 | 90565.21 | 155343.1 | 223630.4 | 69256.71 | 41694.83 |
| 2022-01-17 | 157815.9 | 107898.2 | 26997.29 | 59307.17 | 200021.4 | 90688.09 | 157325.3 | 224936.4 | 72527.78 | 42040.97 |
| 2022-01-18 | 160468.7 | 108554 | 27200.37 | 59368.08 | 203335.2 | 90891.72 | 160167.3 | 226318.7 | 75808.93 | 42510.65 |
| 2022-01-19 | 163282.4 | 109467.8 | 27428.25 | 59368.08 | 206312.2 | 91194.08 | 163298.5 | 227899.4 | 78430.43 | 43048.11 |
| 2022-01-20 | 166126.5 | 110261.2 | 27677.46 | 59506.02 | 208250.9 | 91627.08 | 166315.3 | 229466.3 | 80909.9 | 43502.64 |

**Table 3.** Accumulated number of deaths per million in different regions for the period August 25, 2020 - January 25, 2021, [5].

|  | Argentina | Brazil | India | South Africa | United States | Ukraine | European Union | United Kingdom | Australia | World |
| --- | --- | --- | --- | --- | --- | --- | --- | --- | --- | --- |
| 2020-08-25 | 165.834 | 545.624 | 42.598 | 221.645 | 535.341 | 54.34 | 311.635 | 608.397 | 21.289 | 109.528 |
| 2020-08-26 | 171.886 | 550.666 | 43.399 | 224.876 | 538.879 | 55.192 | 311.74 | 608.632 | 22.181 | 110.361 |
| 2020-08-27 | 176.513 | 555.269 | 44.157 | 226.974 | 542.228 | 56.342 | 312.098 | 608.807 | 22.607 | 111.151 |
| 2020-08-28 | 181.358 | 559.237 | 44.89 | 228.89 | 545.055 | 57.492 | 312.384 | 608.939 | 23.266 | 111.892 |
| 2020-08-29 | 183.156 | 563.428 | 45.57 | 232.854 | 548.008 | 58.435 | 312.567 | 609.115 | 23.693 | 112.631 |
| 2020-08-30 | 185.437 | 565.302 | 46.267 | 233.636 | 549.356 | 59.241 | 312.717 | 609.13 | 25.283 | 113.173 |
| 2020-08-31 | 189.888 | 568.326 | 46.855 | 235.652 | 550.84 | 59.931 | 313.167 | 609.159 | 25.477 | 113.735 |
| 2020-09-01 | 195.567 | 573.7 | 47.605 | 237.55 | 554.027 | 61.058 | 313.621 | 609.203 | 25.709 | 114.573 |
| 2020-09-02 | 199.931 | 579.326 | 48.353 | 239.649 | 557.187 | 62.231 | 314.03 | 609.35 | 26.291 | 115.371 |
| 2020-09-03 | 205.259 | 583.378 | 49.14 | 242.547 | 560.284 | 63.474 | 314.399 | 609.541 | 28.579 | 116.146 |
| 2020-09-04 | 211.004 | 587.345 | 49.921 | 244.462 | 563.225 | 64.693 | 314.983 | 609.687 | 29.005 | 116.891 |
| 2020-09-05 | 213.547 | 590.168 | 50.686 | 246.144 | 565.688 | 65.866 | 315.231 | 609.863 | 29.199 | 117.541 |
| 2020-09-06 | 216.179 | 592.308 | 51.415 | 247.976 | 567.009 | 66.672 | 315.416 | 609.892 | 29.548 | 118.065 |
| 2020-09-07 | 222.099 | 593.803 | 52.228 | 249.892 | 567.971 | 67.408 | 315.859 | 609.936 | 29.859 | 119.285 |
| 2020-09-08 | 228.151 | 596.257 | 53.028 | 251.257 | 569.376 | 68.742 | 316.349 | 610.406 | 30.285 | 119.938 |
| 2020-09-09 | 233.698 | 601.663 | 53.869 | 252.623 | 572.696 | 69.8 | 316.767 | 610.523 | 30.557 | 120.732 |
| 2020-09-10 | 239.158 | 606.215 | 54.737 | 254.239 | 575.471 | 70.836 | 317.185 | 610.728 | 30.906 | 121.496 |
| 2020-09-11 | 244.442 | 610.178 | 55.599 | 256.121 | 578.943 | 72.055 | 317.67 | 610.816 | 31.138 | 122.258 |
| 2020-09-12 | 246.964 | 613.762 | 56.398 | 256.937 | 581.193 | 73.757 | 317.903 | 610.948 | 31.41 | 122.9 |
| 2020-09-13 | 248.916 | 615.654 | 57.214 | 257.27 | 582.395 | 74.517 | 318.104 | 611.021 | 31.642 | 123.401 |
| 2020-09-14 | 255.823 | 617.795 | 57.97 | 258.136 | 583.665 | 75.299 | 318.621 | 611.153 | 31.642 | 123.985 |
| 2020-09-15 | 259.879 | 622.851 | 58.896 | 260.501 | 587.276 | 76.518 | 319.39 | 611.549 | 31.953 | 124.82 |
| 2020-09-16 | 265.668 | 627.346 | 59.708 | 261.567 | 590.145 | 78.313 | 320.32 | 611.842 | 32.263 | 125.567 |
| 2020-09-17 | 273.211 | 631.407 | 60.551 | 262.683 | 592.746 | 79.716 | 321.063 | 612.15 | 32.457 | 126.283 |
| 2020-09-18 | 277.508 | 635.276 | 61.446 | 264.098 | 595.629 | 81.326 | 321.917 | 612.546 | 32.728 | 127.027 |
| 2020-09-19 | 280.644 | 638.459 | 62.259 | 265.481 | 597.765 | 82.477 | 322.23 | 612.942 | 32.922 | 127.702 |
| 2020-09-20 | 286.213 | 640.099 | 63.07 | 265.697 | 598.576 | 83.42 | 322.516 | 613.206 | 33 | 128.206 |
| 2020-09-21 | 295.62 | 642.277 | 63.825 | 266.347 | 599.817 | 84.018 | 323.239 | 613.367 | 33.116 | 128.766 |
| 2020-09-22 | 305.926 | 645.987 | 64.604 | 268.445 | 602.926 | 85.49 | 324.283 | 613.91 | 33.31 | 129.55 |
| 2020-09-23 | 315.223 | 650.342 | 65.414 | 269.911 | 606.125 | 87.055 | 325.128 | 614.452 | 33.387 | 130.381 |
| 2020-09-24 | 323.774 | 654.057 | 66.233 | 271.194 | 608.849 | 88.297 | 325.837 | 615.038 | 33.698 | 131.102 |
| 2020-09-25 | 333.466 | 657.899 | 67.015 | 271.677 | 611.597 | 89.954 | 326.805 | 615.537 | 33.736 | 131.866 |
| 2020-09-26 | 340.812 | 661.273 | 67.821 | 272.742 | 613.949 | 91.748 | 327.355 | 616.05 | 33.814 | 132.556 |
| 2020-09-27 | 345.329 | 662.857 | 68.567 | 273.109 | 614.887 | 93.036 | 327.689 | 616.299 | 33.93 | 133.044 |
| 2020-09-28 | 353.31 | 664.684 | 69.124 | 276.24 | 615.911 | 93.911 | 328.57 | 616.49 | 34.202 | 133.562 |
| 2020-09-29 | 362.213 | 668.693 | 69.97 | 277.589 | 618.62 | 95.567 | 329.187 | 617.531 | 34.357 | 134.337 |
| 2020-09-30 | 371.378 | 673.282 | 70.818 | 278.705 | 621.42 | 97.109 | 330.658 | 618.572 | 34.434 | 135.156 |
| 2020-10-01 | 444.855 | 676.895 | 71.604 | 280.903 | 624.171 | 98.65 | 331.609 | 619.437 | 34.512 | 136.287 |
| 2020-10-02 | 451.675 | 679.951 | 72.371 | 281.62 | 626.697 | 100.237 | 332.648 | 620.404 | 34.628 | 137.008 |
| 2020-10-03 | 455.972 | 682.698 | 73.045 | 282.103 | 628.758 | 102.4 | 333.165 | 621.123 | 34.667 | 137.609 |
| 2020-10-04 | 460.862 | 684.371 | 73.693 | 282.735 | 629.866 | 103.412 | 333.652 | 621.607 | 34.667 | 138.12 |
| 2020-10-05 | 470.729 | 686.208 | 74.328 | 283.402 | 631.314 | 104.217 | 334.594 | 621.885 | 34.706 | 139.018 |
| 2020-10-06 | 478.601 | 689.993 | 75.035 | 284.851 | 633.42 | 106.38 | 336.076 | 623 | 34.783 | 139.761 |
| 2020-10-07 | 487.35 | 693.381 | 75.732 | 287.266 | 636.18 | 108.289 | 337.219 | 624.026 | 34.783 | 140.509 |
| 2020-10-08 | 497.963 | 696.816 | 76.424 | 289.93 | 639.13 | 110.59 | 338.304 | 625.155 | 34.783 | 141.317 |
| 2020-10-09 | 509.255 | 699.872 | 77.089 | 292.245 | 642.053 | 112.707 | 339.686 | 626.43 | 34.783 | 142.103 |
| 2020-10-10 | 517.061 | 702.367 | 77.747 | 294.344 | 644.035 | 115.26 | 340.464 | 627.618 | 34.822 | 142.724 |
| 2020-10-11 | 523.354 | 703.666 | 78.333 | 296.126 | 645.438 | 117.285 | 341.126 | 628.571 | 34.822 | 143.247 |
| 2020-10-12 | 530.327 | 704.573 | 78.84 | 297.508 | 646.633 | 118.32 | 342.476 | 629.304 | 34.861 | 143.763 |
| 2020-10-13 | 538.791 | 706.339 | 79.364 | 300.257 | 648.967 | 120.874 | 343.872 | 631.4 | 35.055 | 144.452 |
| 2020-10-14 | 546.443 | 709.76 | 79.852 | 302.305 | 651.968 | 123.427 | 345.661 | 633.409 | 35.055 | 145.237 |
| 2020-10-15 | 555.675 | 713.564 | 80.494 | 304.937 | 654.506 | 125.222 | 347.291 | 635.432 | 35.055 | 146.023 |
| 2020-10-16 | 564.029 | 716.569 | 81.095 | 305.953 | 657.339 | 127.707 | 349.301 | 637.426 | 35.055 | 146.812 |
| 2020-10-17 | 572.449 | 718.508 | 81.836 | 306.585 | 659.802 | 130.421 | 350.444 | 639.625 | 35.055 | 147.535 |
| 2020-10-18 | 575.957 | 719.606 | 82.252 | 307.635 | 661.193 | 132.561 | 351.537 | 640.608 | 35.094 | 148.026 |
| 2020-10-19 | 585.802 | 721.13 | 82.673 | 307.984 | 662.664 | 134.217 | 353.409 | 641.781 | 35.094 | 148.627 |
| 2020-10-20 | 594.222 | 724.158 | 83.187 | 310.716 | 665.479 | 136.84 | 355.918 | 645.314 | 35.094 | 149.47 |
| 2020-10-21 | 603.41 | 726.826 | 83.691 | 312.132 | 669.005 | 140.176 | 358.286 | 648.114 | 35.094 | 150.341 |
| 2020-10-22 | 613.014 | 729.186 | 84.186 | 313.83 | 671.58 | 142.937 | 360.813 | 650.885 | 35.094 | 151.093 |
| 2020-10-23 | 621.368 | 731.817 | 84.653 | 314.63 | 674.433 | 145.766 | 363.667 | 654.169 | 35.094 | 151.989 |
| 2020-10-24 | 627.398 | 733.625 | 85.068 | 315.512 | 677.326 | 148.688 | 365.887 | 656.72 | 35.094 | 152.733 |
| 2020-10-25 | 633.603 | 734.714 | 85.412 | 315.912 | 678.702 | 151.058 | 367.296 | 658.934 | 35.094 | 153.274 |
| 2020-10-26 | 642.484 | 736.125 | 85.762 | 316.578 | 680.27 | 152.783 | 370.487 | 660.43 | 35.094 | 153.989 |
| 2020-10-27 | 651.89 | 738.583 | 86.127 | 317.328 | 683.306 | 155.751 | 374.673 | 665.81 | 35.171 | 154.936 |
| 2020-10-28 | 659.368 | 740.939 | 86.498 | 318.294 | 686.412 | 159.616 | 378.204 | 670.355 | 35.171 | 155.851 |
| 2020-10-29 | 667.502 | 743.513 | 86.902 | 319.177 | 689.425 | 162.377 | 382.017 | 674.46 | 35.171 | 156.766 |
| 2020-10-30 | 675.177 | 746.191 | 87.297 | 320.276 | 692.621 | 166.449 | 386.657 | 678.478 | 35.171 | 157.743 |
| 2020-10-31 | 679.782 | 747.556 | 87.635 | 321.042 | 695.418 | 170.222 | 390.232 | 683.257 | 35.171 | 158.58 |
| 2020-11-01 | 682.808 | 748.504 | 87.991 | 323.29 | 696.928 | 172.89 | 393.046 | 685.632 | 35.171 | 159.232 |
| 2020-11-02 | 693.398 | 749.275 | 88.342 | 324.19 | 698.602 | 174.478 | 397.565 | 687.626 | 35.171 | 160.016 |
| 2020-11-03 | 702.805 | 750.621 | 88.711 | 325.422 | 703.399 | 178.274 | 404.819 | 693.447 | 35.171 | 161.22 |
| 2020-11-04 | 713.067 | 753.509 | 89.216 | 326.188 | 706.763 | 182.944 | 412.972 | 700.66 | 35.171 | 162.51 |
| 2020-11-05 | 718.461 | 756.327 | 89.697 | 327.721 | 710.262 | 187.453 | 418.876 | 706.202 | 35.171 | 163.616 |
| 2020-11-06 | 726.574 | 757.593 | 90.111 | 328.92 | 713.909 | 192.285 | 426.081 | 711.407 | 35.171 | 164.772 |
| 2020-11-07 | 731.222 | 758.659 | 90.513 | 329.586 | 717.264 | 196.771 | 430.897 | 717.462 | 35.171 | 165.747 |
| 2020-11-08 | 735.871 | 759.547 | 90.864 | 329.919 | 718.955 | 200.038 | 434.645 | 719.749 | 35.171 | 166.521 |
| 2020-11-09 | 743.48 | 760.416 | 91.186 | 330.519 | 721.235 | 202.729 | 440.334 | 722.593 | 35.171 | 167.432 |
| 2020-11-10 | 749.531 | 761.341 | 91.553 | 332.284 | 725.488 | 207.469 | 449.024 | 730.393 | 35.171 | 168.71 |
| 2020-11-11 | 757.162 | 764.023 | 91.948 | 333.283 | 729.823 | 211.978 | 457.059 | 739.116 | 35.171 | 170.046 |
| 2020-11-12 | 762.666 | 768.383 | 92.34 | 334.366 | 733.439 | 216.763 | 463.671 | 747.371 | 35.171 | 171.289 |
| 2020-11-13 | 768.433 | 771.075 | 92.714 | 335.648 | 737.044 | 220.95 | 471.63 | 752.883 | 35.171 | 172.528 |
| 2020-11-14 | 774.177 | 774.505 | 93.034 | 336.531 | 741.168 | 225.597 | 477.327 | 759.657 | 35.171 | 173.678 |
| 2020-11-15 | 777.006 | 775.159 | 93.347 | 337.114 | 743.535 | 227.852 | 482.052 | 762.12 | 35.171 | 174.515 |
| 2020-11-16 | 783.387 | 776.36 | 93.669 | 338.33 | 745.98 | 230.107 | 488.935 | 765.243 | 35.171 | 175.523 |
| 2020-11-17 | 791.697 | 779.529 | 94.009 | 340.295 | 751.113 | 233.926 | 498.479 | 774.01 | 35.171 | 176.93 |
| 2020-11-18 | 796.982 | 783.052 | 94.429 | 342.36 | 756.947 | 239.976 | 506.941 | 781.766 | 35.171 | 178.384 |
| 2020-11-19 | 801.038 | 786.09 | 94.848 | 344.276 | 763.08 | 246.027 | 514.394 | 789.111 | 35.171 | 179.8 |
| 2020-11-20 | 806.695 | 788.487 | 95.253 | 345.741 | 768.935 | 251.433 | 523.938 | 796.603 | 35.171 | 181.294 |
| 2020-11-21 | 809.151 | 790.155 | 95.612 | 347.174 | 773.855 | 256.494 | 529.969 | 801.588 | 35.171 | 182.439 |
| 2020-11-22 | 811.344 | 790.987 | 95.979 | 348.14 | 777.042 | 259.784 | 534.652 | 807.423 | 35.171 | 183.382 |
| 2020-11-23 | 813.975 | 792.646 | 96.323 | 349.222 | 780.265 | 262.798 | 541.962 | 810.443 | 35.171 | 184.465 |
| 2020-11-24 | 820.772 | 795.576 | 96.669 | 351.138 | 786.681 | 267.307 | 552.425 | 819.357 | 35.171 | 186.081 |
| 2020-11-25 | 826.956 | 798.487 | 97.045 | 353.103 | 793.449 | 272.783 | 561.035 | 829.547 | 35.171 | 187.619 |
| 2020-11-26 | 831.933 | 801.805 | 97.398 | 354.568 | 797.663 | 278.166 | 569.653 | 836.848 | 35.171 | 189.017 |
| 2020-11-27 | 837.963 | 804.417 | 97.746 | 356.051 | 802.325 | 282.79 | 579.476 | 844.472 | 35.171 | 190.472 |
| 2020-11-28 | 840.287 | 806.959 | 98.102 | 357.067 | 806.404 | 287.231 | 585.805 | 851.495 | 35.171 | 191.646 |
| 2020-11-29 | 843.598 | 808.048 | 98.42 | 357.7 | 809.561 | 290.175 | 590.543 | 854.647 | 35.21 | 192.572 |
| 2020-11-30 | 849.234 | 809.688 | 98.766 | 358.666 | 813.502 | 292.89 | 597.815 | 857.623 | 35.21 | 193.706 |
| 2020-12-01 | 853.575 | 812.889 | 99.125 | 360.481 | 821.167 | 298.204 | 607.444 | 866.464 | 35.21 | 195.322 |
| 2020-12-02 | 858.575 | 816.062 | 99.503 | 361.564 | 829.668 | 302.3 | 616.177 | 875.964 | 35.21 | 196.93 |
| 2020-12-03 | 861.842 | 819.619 | 99.89 | 363.129 | 838.586 | 308.143 | 625.054 | 882.034 | 35.21 | 198.527 |
| 2020-12-04 | 866.381 | 822.787 | 100.258 | 365.794 | 846.432 | 313.826 | 634.048 | 889.423 | 35.21 | 200.107 |
| 2020-12-05 | 869.012 | 825.81 | 100.604 | 367.526 | 853.611 | 319.255 | 640.243 | 895.244 | 35.21 | 201.418 |
| 2020-12-06 | 872.038 | 827.399 | 100.884 | 369.841 | 857.825 | 323.327 | 644.959 | 898.631 | 35.21 | 202.397 |
| 2020-12-07 | 874.625 | 829.385 | 101.161 | 370.557 | 862.508 | 326.87 | 651.258 | 901.402 | 35.21 | 203.503 |
| 2020-12-08 | 877.278 | 833.114 | 101.449 | 373.605 | 870.267 | 331.586 | 659.677 | 910.184 | 35.21 | 205.063 |
| 2020-12-09 | 881.949 | 837.1 | 101.745 | 375.97 | 879.822 | 338.235 | 667.622 | 917.998 | 35.21 | 206.673 |
| 2020-12-10 | 886.532 | 840.694 | 102.041 | 378.851 | 888.791 | 344.654 | 676.104 | 925.563 | 35.21 | 208.281 |
| 2020-12-11 | 890.369 | 843.783 | 102.359 | 382.266 | 898.947 | 351.486 | 685.252 | 931.78 | 35.21 | 209.94 |
| 2020-12-12 | 891.728 | 846.947 | 102.64 | 384.831 | 906.556 | 357.238 | 691.545 | 939.403 | 35.21 | 211.301 |
| 2020-12-13 | 893.877 | 848.325 | 102.881 | 387.662 | 911.539 | 360.988 | 696.299 | 941.515 | 35.21 | 212.29 |
| 2020-12-14 | 899.907 | 850.722 | 103.135 | 390.577 | 916.492 | 363.312 | 702.867 | 944.916 | 35.21 | 213.454 |
| 2020-12-15 | 903.481 | 855.087 | 103.413 | 394.074 | 925.464 | 368.971 | 713.283 | 952.335 | 35.21 | 215.198 |
| 2020-12-16 | 907.011 | 859.484 | 103.667 | 396.839 | 936.677 | 375.321 | 722.592 | 961.307 | 35.21 | 216.959 |
| 2020-12-17 | 910.717 | 864.442 | 103.91 | 399.903 | 947.04 | 381.417 | 731.108 | 969.107 | 35.21 | 218.656 |
| 2020-12-18 | 913.743 | 868.26 | 104.159 | 404.467 | 955.877 | 387.606 | 740.256 | 976.291 | 35.21 | 220.294 |
| 2020-12-19 | 915.738 | 871.34 | 104.404 | 408.697 | 964.15 | 392.759 | 746.473 | 984.164 | 35.21 | 221.684 |
| 2020-12-20 | 916.835 | 873.298 | 104.643 | 411.229 | 969.463 | 395.727 | 750.679 | 988.944 | 35.21 | 222.711 |
| 2020-12-21 | 920.869 | 875.919 | 104.859 | 414.826 | 974.861 | 397.844 | 757.39 | 992.096 | 35.21 | 223.923 |
| 2020-12-22 | 926.504 | 880.448 | 105.098 | 420.472 | 984.768 | 403.411 | 767.021 | 1002.227 | 35.21 | 225.724 |
| 2020-12-23 | 927.82 | 884.957 | 105.322 | 427.318 | 994.917 | 410.037 | 775.623 | 1013.193 | 35.21 | 227.479 |
| 2020-12-24 | 927.82 | 888.509 | 105.563 | 432.747 | 1003.694 | 415.42 | 782.057 | 1021.624 | 35.21 | 228.98 |
| 2020-12-25 | 930.188 | 890.761 | 105.743 | 437.627 | 1008.215 | 419.975 | 786.444 | 1029.981 | 35.21 | 230.083 |
| 2020-12-26 | 931.92 | 892.144 | 105.943 | 441.708 | 1013.961 | 423.058 | 790.203 | 1033.059 | 35.21 | 231.041 |
| 2020-12-27 | 935.188 | 893.719 | 106.143 | 445.272 | 1018.281 | 424.968 | 794.032 | 1038.147 | 35.249 | 232.02 |
| 2020-12-28 | 939.968 | 896.233 | 106.324 | 450.868 | 1023.919 | 426.877 | 800.758 | 1043.381 | 35.249 | 233.249 |
| 2020-12-29 | 943.257 | 901.144 | 106.529 | 459.145 | 1034.78 | 432.537 | 811.512 | 1050.096 | 35.249 | 235.165 |
| 2020-12-30 | 946.436 | 906.832 | 106.744 | 466.89 | 1046.594 | 438.449 | 821.329 | 1064.493 | 35.249 | 237.149 |
| 2020-12-31 | 948.234 | 911.579 | 106.928 | 474.151 | 1056.588 | 443.58 | 828.44 | 1078.626 | 35.249 | 238.831 |
| 2021-01-01 | 949.857 | 913.771 | 107.088 | 481.113 | 1063.178 | 447.169 | 833.576 | 1087.643 | 35.249 | 240.06 |
| 2021-01-02 | 951.085 | 915.14 | 107.244 | 485.91 | 1070.985 | 448.572 | 837.465 | 1094.167 | 35.249 | 241.134 |
| 2021-01-03 | 953.431 | 916.594 | 107.398 | 492.605 | 1075.301 | 451.609 | 841.226 | 1100.824 | 35.249 | 242.102 |
| 2021-01-04 | 956.764 | 919.383 | 107.542 | 499.833 | 1081.249 | 453.495 | 848.096 | 1106.835 | 35.249 | 243.379 |
| 2021-01-05 | 960.075 | 924.935 | 107.731 | 508.378 | 1092.074 | 458.488 | 858.62 | 1119.707 | 35.249 | 245.337 |
| 2021-01-06 | 964.263 | 930.687 | 107.891 | 522.434 | 1103.861 | 464.055 | 866.723 | 1134.97 | 35.249 | 247.254 |
| 2021-01-07 | 967.464 | 937.781 | 108.059 | 529.779 | 1115.96 | 467.805 | 874.87 | 1152.006 | 35.249 | 249.178 |
| 2021-01-08 | 970.775 | 942.833 | 108.059 | 540.039 | 1128.543 | 470.06 | 883.893 | 1171.52 | 35.249 | 251.132 |
| 2021-01-09 | 973.933 | 947.548 | 108.367 | 546.684 | 1138.323 | 472.222 | 889.81 | 1186.694 | 35.249 | 252.759 |
| 2021-01-10 | 975.643 | 949.838 | 108.482 | 552.33 | 1144.643 | 474.868 | 893.969 | 1194.949 | 35.249 | 253.875 |
| 2021-01-11 | 979.129 | 952.132 | 108.602 | 559.259 | 1150.161 | 476.662 | 900.794 | 1202.704 | 35.249 | 255.152 |
| 2021-01-12 | 983.383 | 957.389 | 108.747 | 571.833 | 1163.345 | 481.172 | 910.897 | 1220.958 | 35.249 | 257.329 |
| 2021-01-13 | 986.343 | 963.296 | 108.889 | 585.257 | 1175.414 | 485.911 | 920.148 | 1243.903 | 35.249 | 259.473 |
| 2021-01-14 | 989.457 | 968.689 | 109.026 | 597.115 | 1186.804 | 490.029 | 928.91 | 1262.2 | 35.249 | 261.441 |
| 2021-01-15 | 991.694 | 973.992 | 109.152 | 607.358 | 1198.255 | 494.147 | 936.797 | 1280.966 | 35.249 | 263.351 |
| 2021-01-16 | 993.185 | 978.731 | 109.282 | 613.754 | 1208.999 | 497.782 | 943.078 | 1299.952 | 35.249 | 265.094 |
| 2021-01-17 | 995.64 | 981.306 | 109.386 | 617.984 | 1214.98 | 500.773 | 947.266 | 1309.819 | 35.249 | 266.25 |
| 2021-01-18 | 1004.959 | 983.53 | 109.484 | 623.713 | 1219.68 | 502.613 | 952.812 | 1318.601 | 35.249 | 267.486 |
| 2021-01-19 | 1010.09 | 988.951 | 109.6 | 637.687 | 1226.892 | 506.984 | 964.036 | 1342.206 | 35.249 | 269.596 |
| 2021-01-20 | 1013.379 | 995.47 | 109.709 | 647.114 | 1240.235 | 512.207 | 973.767 | 1368.889 | 35.249 | 271.889 |
| 2021-01-21 | 1016.427 | 1001.764 | 109.826 | 657.89 | 1252.662 | 518.119 | 982.263 | 1387.802 | 35.249 | 274.066 |
| 2021-01-22 | 1021.251 | 1006.769 | 109.935 | 667.466 | 1264.007 | 522.191 | 991.051 | 1408.343 | 35.249 | 276.113 |
| 2021-01-23 | 1024.803 | 1012.12 | 110.046 | 675.76 | 1274.688 | 525.228 | 996.308 | 1428.121 | 35.249 | 277.881 |
| 2021-01-24 | 1026.777 | 1014.9 | 110.14 | 680.757 | 1280.269 | 527.437 | 1000.535 | 1437.064 | 35.249 | 279.049 |
| 2021-01-25 | 1031.316 | 1018.125 | 110.224 | 684.804 | 1285.586 | 529.162 | 1007.881 | 1445.744 | 35.249 | 280.452 |

**Table 4.** Accumulated number of deaths per million for the period August 25, 2021 - January 20, 2022. [5].

|  | Argentina | Brazil | India | South Africa | United States | Ukraine | European Union | United Kingdom | Australia | World |
| --- | --- | --- | --- | --- | --- | --- | --- | --- | --- | --- |
| 2021-08-25 | 2433.154 | 2696.199 | 313.164 | 1340.212 | 1900.893 | 1302.511 | 1680.514 | 1936.704 | 38.351 | 566.954 |
| 2021-08-26 | 2436.465 | 2700.27 | 313.52 | 1346.158 | 1907.565 | 1304.029 | 1681.646 | 1938.771 | 38.428 | 568.428 |
| 2021-08-27 | 2439.82 | 2704.092 | 313.885 | 1352.17 | 1912.74 | 1306.031 | 1682.665 | 1940.237 | 38.506 | 569.736 |
| 2021-08-28 | 2441.004 | 2707.032 | 314.215 | 1356.734 | 1914.641 | 1307.825 | 1683.262 | 1942.187 | 38.739 | 570.811 |
| 2021-08-29 | 2442.298 | 2708.331 | 314.488 | 1358.965 | 1915.51 | 1309.044 | 1683.745 | 1943.082 | 38.894 | 571.663 |
| 2021-08-30 | 2447.209 | 2709.817 | 314.739 | 1362.879 | 1920.715 | 1309.827 | 1685 | 1943.786 | 39.01 | 572.873 |
| 2021-08-31 | 2451.704 | 2713.934 | 315.069 | 1370.058 | 1925.05 | 1311.437 | 1686.384 | 1944.519 | 39.243 | 574.039 |
| 2021-09-01 | 2455.936 | 2717.191 | 315.434 | 1373.972 | 1931.159 | 1312.863 | 1687.569 | 1947.553 | 39.514 | 575.622 |
| 2021-09-02 | 2460.103 | 2720.845 | 315.697 | 1380.933 | 1940.029 | 1314.313 | 1688.837 | 1950.163 | 40.018 | 577.049 |
| 2021-09-03 | 2463.633 | 2724.303 | 315.934 | 1385.047 | 1945.941 | 1315.762 | 1689.989 | 1951.937 | 40.173 | 578.405 |
| 2021-09-04 | 2465.562 | 2727.285 | 316.155 | 1388.078 | 1947.848 | 1316.774 | 1690.678 | 1953.697 | 40.29 | 579.383 |
| 2021-09-05 | 2467.031 | 2728.551 | 316.312 | 1389.344 | 1949.038 | 1317.948 | 1691.223 | 1954.693 | 40.484 | 580.21 |
| 2021-09-06 | 2470.584 | 2729.579 | 316.52 | 1392.642 | 1950.837 | 1318.684 | 1692.473 | 1955.353 | 40.794 | 581.23 |
| 2021-09-07 | 2474.487 | 2731.107 | 316.785 | 1397.339 | 1957.082 | 1320.363 | 1693.947 | 1958.417 | 41.104 | 582.559 |
| 2021-09-08 | 2476.921 | 2732.271 | 317.028 | 1401.552 | 1963.927 | 1322.204 | 1695.215 | 1961.218 | 41.337 | 583.834 |
| 2021-09-09 | 2479.925 | 2736.121 | 317.214 | 1404.467 | 1973.792 | 1324.044 | 1696.51 | 1963.666 | 41.724 | 585.212 |
| 2021-09-10 | 2483.937 | 2739.093 | 317.435 | 1409.147 | 1981.091 | 1326.253 | 1697.664 | 1965.821 | 42.035 | 586.375 |
| 2021-09-11 | 2485.56 | 2742.22 | 317.678 | 1411.529 | 1983.47 | 1328.254 | 1698.285 | 1968.108 | 42.306 | 587.433 |
| 2021-09-12 | 2486.568 | 2743.617 | 317.835 | 1413.627 | 1984.479 | 1329.106 | 1698.82 | 1968.93 | 42.578 | 588.138 |
| 2021-09-13 | 2491.787 | 2744.944 | 318.078 | 1415.709 | 1990.952 | 1329.681 | 1700.224 | 1969.839 | 42.733 | 589.292 |
| 2021-09-14 | 2495.646 | 2748.112 | 318.282 | 1420.706 | 1996.74 | 1332.35 | 1702.111 | 1972.551 | 43.276 | 590.551 |
| 2021-09-15 | 2499.001 | 2751.855 | 318.591 | 1423.47 | 2005.049 | 1334.926 | 1703.424 | 1975.498 | 43.741 | 591.878 |
| 2021-09-16 | 2501.895 | 2754.954 | 318.821 | 1428.65 | 2015.445 | 1337.71 | 1704.672 | 1977.814 | 44.245 | 593.212 |
| 2021-09-17 | 2505.952 | 2757.935 | 319.023 | 1431.531 | 2022.792 | 1340.448 | 1705.855 | 1980.439 | 44.516 | 594.354 |
| 2021-09-18 | 2507.728 | 2760.673 | 319.244 | 1434.263 | 2025.688 | 1342.702 | 1706.678 | 1982.843 | 45.059 | 595.177 |
| 2021-09-19 | 2509.066 | 2761.832 | 319.456 | 1435.229 | 2026.73 | 1344.06 | 1707.248 | 1983.664 | 45.253 | 595.947 |
| 2021-09-20 | 2511.039 | 2763.071 | 319.637 | 1435.928 | 2033.594 | 1345.325 | 1708.726 | 1984.382 | 45.68 | 596.987 |
| 2021-09-21 | 2512.377 | 2766.216 | 319.912 | 1438.593 | 2040.668 | 1348.822 | 1710.383 | 1987.359 | 45.99 | 598.169 |
| 2021-09-22 | 2514.679 | 2769.323 | 320.114 | 1440.658 | 2049.216 | 1351.698 | 1711.939 | 1989.792 | 46.378 | 599.429 |
| 2021-09-23 | 2516.608 | 2772.263 | 320.342 | 1443.24 | 2059.03 | 1354.941 | 1713.393 | 1992.461 | 46.843 | 600.669 |
| 2021-09-24 | 2517.836 | 2775.44 | 320.551 | 1448.436 | 2066.491 | 1358.576 | 1714.757 | 1995.1 | 47.308 | 601.784 |
| 2021-09-25 | 2518.297 | 2777.964 | 320.737 | 1449.002 | 2068.864 | 1362.004 | 1715.468 | 1996.888 | 47.735 | 602.473 |
| 2021-09-26 | 2518.582 | 2779.207 | 320.935 | 1449.852 | 2070.513 | 1363.822 | 1716.148 | 1997.812 | 48.278 | 603.162 |
| 2021-09-27 | 2520.599 | 2780.113 | 321.064 | 1452.583 | 2077.434 | 1366.191 | 1717.715 | 1998.399 | 48.704 | 604.138 |
| 2021-09-28 | 2522.441 | 2783.931 | 321.335 | 1455.931 | 2085.123 | 1369.78 | 1719.372 | 2000.847 | 49.596 | 605.333 |
| 2021-09-29 | 2524.458 | 2787.113 | 321.558 | 1455.931 | 2092.972 | 1375.026 | 1720.989 | 2003.046 | 50.062 | 606.524 |
| 2021-09-30 | 2525.533 | 2789.955 | 321.757 | 1459.412 | 2101.377 | 1379.903 | 1722.416 | 2005.055 | 50.837 | 607.644 |
| 2021-10-01 | 2526.541 | 2792.244 | 321.925 | 1460.728 | 2107.793 | 1384.136 | 1723.818 | 2006.917 | 51.225 | 608.735 |
| 2021-10-02 | 2526.848 | 2794.366 | 322.1 | 1461.527 | 2110.178 | 1389.105 | 1724.737 | 2008.691 | 51.729 | 609.48 |
| 2021-10-03 | 2526.98 | 2795.422 | 322.229 | 1461.977 | 2111.677 | 1392.188 | 1725.555 | 2009.321 | 52.194 | 610.022 |
| 2021-10-04 | 2527.813 | 2796.375 | 322.418 | 1462.626 | 2117.561 | 1394.949 | 1727.179 | 2009.805 | 52.621 | 610.933 |
| 2021-10-05 | 2528.69 | 2799.572 | 322.617 | 1464.342 | 2123.289 | 1402.679 | 1729.138 | 2012.239 | 53.474 | 611.876 |
| 2021-10-06 | 2529.918 | 2802.235 | 322.846 | 1465.324 | 2131.192 | 1410.478 | 1731.061 | 2014.365 | 53.862 | 613.038 |
| 2021-10-07 | 2530.729 | 2804.235 | 323.04 | 1467.373 | 2138.639 | 1418.024 | 1732.758 | 2016.153 | 54.482 | 614.176 |
| 2021-10-08 | 2531.343 | 2807.179 | 323.218 | 1469.571 | 2144.147 | 1423.914 | 1734.69 | 2017.986 | 55.103 | 615.138 |
| 2021-10-09 | 2531.65 | 2809.123 | 323.372 | 1470.504 | 2145.415 | 1429.987 | 1735.911 | 2020.273 | 55.529 | 615.751 |
| 2021-10-10 | 2531.979 | 2809.829 | 323.51 | 1470.92 | 2146.821 | 1434.013 | 1736.786 | 2020.83 | 56.15 | 616.343 |
| 2021-10-11 | 2532.374 | 2810.796 | 323.64 | 1471.403 | 2150.131 | 1439.029 | 1738.449 | 2021.241 | 56.654 | 617.074 |
| 2021-10-12 | 2533.602 | 2811.614 | 323.802 | 1472.786 | 2157.199 | 1447.564 | 1740.672 | 2023.894 | 57.313 | 618.107 |
| 2021-10-13 | 2534.369 | 2812.577 | 323.979 | 1473.402 | 2166.892 | 1458.906 | 1742.64 | 2025.888 | 58.011 | 619.225 |
| 2021-10-14 | 2535.488 | 2815.25 | 324.251 | 1474.068 | 2173.329 | 1468.867 | 1744.666 | 2028.19 | 58.438 | 620.244 |
| 2021-10-15 | 2536.08 | 2817.619 | 324.37 | 1475.001 | 2178.817 | 1473.952 | 1746.714 | 2030.272 | 58.748 | 621.163 |
| 2021-10-16 | 2536.145 | 2819.792 | 324.473 | 1475.417 | 2180.397 | 1480.831 | 1748.107 | 2032.442 | 59.407 | 621.83 |
| 2021-10-17 | 2536.211 | 2820.39 | 324.592 | 1475.834 | 2181.755 | 1486.26 | 1749.315 | 2033.278 | 59.834 | 622.372 |
| 2021-10-18 | 2537.044 | 2821.334 | 324.71 | 1475.95 | 2186.993 | 1490.723 | 1751.386 | 2033.937 | 60.415 | 623.192 |
| 2021-10-19 | 2537.768 | 2823.105 | 324.851 | 1476.866 | 2193.412 | 1503.653 | 1754.098 | 2037.207 | 61.152 | 624.242 |
| 2021-10-20 | 2538.492 | 2824.97 | 324.966 | 1478.199 | 2203.033 | 1515.547 | 1756.5 | 2039.831 | 61.656 | 625.347 |
| 2021-10-21 | 2539.062 | 2827.147 | 325.132 | 1479.548 | 2209.005 | 1528.683 | 1758.803 | 2041.547 | 62.47 | 626.325 |
| 2021-10-22 | 2539.566 | 2829.236 | 325.61 | 1480.48 | 2214.7 | 1543.407 | 1760.907 | 2044.186 | 62.897 | 627.366 |
| 2021-10-23 | 2539.654 | 2830.872 | 326.013 | 1480.863 | 2216.319 | 1555.14 | 1762.848 | 2046.165 | 63.479 | 628.103 |
| 2021-10-24 | 2539.72 | 2831.428 | 326.331 | 1481.047 | 2217.052 | 1564.596 | 1764.248 | 2047.22 | 63.905 | 628.677 |
| 2021-10-25 | 2540.268 | 2832.395 | 326.586 | 1481.197 | 2221.299 | 1572.809 | 1766.525 | 2047.778 | 64.099 | 629.522 |
| 2021-10-26 | 2540.597 | 2834.278 | 327.006 | 1482.079 | 2226.159 | 1590.408 | 1769.635 | 2051.633 | 64.719 | 630.526 |
| 2021-10-27 | 2541.101 | 2836.311 | 327.532 | 1483.112 | 2232.395 | 1607.065 | 1772.882 | 2054.668 | 65.766 | 631.692 |
| 2021-10-28 | 2541.693 | 2838.054 | 328.11 | 1484.028 | 2237.928 | 1621.352 | 1775.434 | 2057.102 | 66.232 | 632.779 |
| 2021-10-29 | 2542.11 | 2839.961 | 328.504 | 1484.811 | 2243.143 | 1636.674 | 1778.39 | 2059.829 | 66.775 | 633.805 |
| 2021-10-30 | 2542.263 | 2840.97 | 328.824 | 1485.011 | 2244.23 | 1649.764 | 1780.431 | 2062.263 | 67.279 | 634.565 |
| 2021-10-31 | 2542.438 | 2841.592 | 329.004 | 1485.244 | 2244.93 | 1658.115 | 1782.131 | 2063.348 | 67.589 | 635.134 |
| 2021-11-01 | 2543.294 | 2842.073 | 329.322 | 1485.277 | 2248.685 | 1665.684 | 1784.727 | 2063.934 | 68.093 | 635.932 |
| 2021-11-02 | 2543.754 | 2842.783 | 329.545 | 1485.577 | 2252.544 | 1682.571 | 1788.019 | 2068.215 | 68.558 | 636.855 |
| 2021-11-03 | 2544.171 | 2843.671 | 329.876 | 1485.96 | 2258.453 | 1699.871 | 1791.559 | 2071.397 | 69.063 | 637.926 |
| 2021-11-04 | 2544.741 | 2845.699 | 330.034 | 1486.476 | 2262.081 | 1716.643 | 1795.193 | 2074.534 | 69.605 | 638.894 |
| 2021-11-05 | 2545.355 | 2847.484 | 330.316 | 1487.209 | 2268.458 | 1733.414 | 1798.52 | 2077.364 | 69.993 | 639.986 |
| 2021-11-06 | 2545.53 | 2848.9 | 330.693 | 1487.609 | 2269.843 | 1752.486 | 1801.031 | 2079.636 | 70.381 | 640.826 |
| 2021-11-07 | 2545.815 | 2849.228 | 330.884 | 1487.825 | 2270.405 | 1763.575 | 1802.621 | 2080.545 | 70.846 | 641.394 |
| 2021-11-08 | 2546.67 | 2849.788 | 331.122 | 1488.158 | 2274.051 | 1775.055 | 1805.913 | 2081.381 | 71.389 | 642.242 |
| 2021-11-09 | 2547.153 | 2850.76 | 331.453 | 1488.741 | 2278.518 | 1794.863 | 1809.945 | 2085.222 | 72.048 | 643.289 |
| 2021-11-10 | 2547.569 | 2852.017 | 331.697 | 1489.541 | 2283.57 | 1814.372 | 1814.22 | 2088.36 | 72.204 | 644.325 |
| 2021-11-11 | 2548.118 | 2853.139 | 332.056 | 1489.824 | 2286.154 | 1830.085 | 1817.876 | 2091.219 | 72.63 | 645.303 |
| 2021-11-12 | 2548.403 | 2856.041 | 332.454 | 1490.107 | 2293.636 | 1848.076 | 1821.126 | 2093.345 | 72.785 | 646.448 |
| 2021-11-13 | 2548.534 | 2857.475 | 332.659 | 1490.224 | 2295.306 | 1864.733 | 1823.865 | 2095.647 | 72.979 | 647.242 |
| 2021-11-14 | 2548.622 | 2857.779 | 332.749 | 1490.357 | 2295.877 | 1874.579 | 1825.674 | 2096.57 | 73.212 | 647.826 |
| 2021-11-15 | 2549.017 | 2858.106 | 332.89 | 1490.44 | 2299.695 | 1885.3 | 1829.458 | 2097.259 | 73.6 | 648.713 |
| 2021-11-16 | 2549.981 | 2858.775 | 333.106 | 1490.69 | 2303.72 | 1905.2 | 1833.778 | 2100.397 | 73.949 | 649.69 |
| 2021-11-17 | 2550.398 | 2860.536 | 333.443 | 1490.873 | 2308.778 | 1923.513 | 1838.651 | 2103.344 | 74.53 | 650.858 |
| 2021-11-18 | 2551.012 | 2861.807 | 333.773 | 1491.539 | 2312.764 | 1941.55 | 1842.712 | 2106.261 | 74.957 | 651.91 |
| 2021-11-19 | 2551.429 | 2862.92 | 333.964 | 1491.656 | 2318.189 | 1958.988 | 1846.831 | 2108.563 | 75.151 | 652.93 |
| 2021-11-20 | 2551.736 | 2863.887 | 334.189 | 1491.822 | 2319.535 | 1975.001 | 1849.722 | 2110.762 | 75.383 | 653.7 |
| 2021-11-21 | 2551.801 | 2864.335 | 334.368 | 1491.856 | 2320.132 | 1984.502 | 1851.336 | 2111.657 | 75.538 | 654.274 |
| 2021-11-22 | 2551.911 | 2864.924 | 334.537 | 1492.022 | 2323.845 | 1992.669 | 1855.887 | 2112.316 | 76.314 | 655.194 |
| 2021-11-23 | 2552.635 | 2866.303 | 334.851 | 1492.872 | 2327.975 | 2010.062 | 1860.487 | 2114.735 | 76.508 | 656.217 |
| 2021-11-24 | 2553.577 | 2867.569 | 335.135 | 1493.238 | 2333.229 | 2024.51 | 1865.523 | 2116.92 | 76.702 | 657.318 |
| 2021-11-25 | 2554.06 | 2868.915 | 335.485 | 1495.137 | 2334.625 | 2039.809 | 1870.23 | 2119.075 | 76.973 | 658.302 |
| 2021-11-26 | 2554.608 | 2870.34 | 335.819 | 1495.337 | 2335.728 | 2054.602 | 1874.472 | 2121.421 | 77.167 | 659.166 |
| 2021-11-27 | 2554.871 | 2871.401 | 336.265 | 1495.47 | 2336.659 | 2068.497 | 1877.464 | 2123.342 | 77.322 | 659.916 |
| 2021-11-28 | 2555.134 | 2871.775 | 336.434 | 1495.57 | 2337.323 | 2078.482 | 1879.51 | 2124.089 | 77.438 | 660.52 |
| 2021-11-29 | 2555.682 | 2872.326 | 336.57 | 1495.986 | 2343.093 | 2086.028 | 1884.045 | 2124.602 | 77.787 | 661.507 |
| 2021-11-30 | 2556.45 | 2873.85 | 336.762 | 1496.336 | 2347.671 | 2099.716 | 1889.233 | 2126.948 | 77.981 | 662.517 |
| 2021-12-01 | 2556.625 | 2875.102 | 337.104 | 1496.802 | 2353.994 | 2113.336 | 1894.269 | 2129.455 | 78.369 | 663.693 |
| 2021-12-02 | 2557.064 | 2876.018 | 337.385 | 1497.535 | 2365.705 | 2126.173 | 1899.171 | 2131.523 | 78.834 | 664.976 |
| 2021-12-03 | 2557.546 | 2877.112 | 337.683 | 1498.018 | 2370.611 | 2138.643 | 1904.204 | 2133.619 | 79.183 | 666.053 |
| 2021-12-04 | 2557.634 | 2877.794 | 337.683 | 1498.368 | 2372.605 | 2149.363 | 1907.42 | 2135.481 | 79.494 | 666.749 |
| 2021-12-05 | 2557.7 | 2878.126 | 339.841 | 1498.385 | 2373.608 | 2156.449 | 1909.12 | 2136.273 | 79.726 | 667.678 |
| 2021-12-06 | 2558.445 | 2878.742 | 339.999 | 1498.534 | 2377.651 | 2162.615 | 1913.274 | 2136.889 | 80.075 | 668.567 |
| 2021-12-07 | 2558.95 | 2879.962 | 340.138 | 1498.984 | 2382.755 | 2174.049 | 1919.509 | 2139.528 | 80.347 | 669.621 |
| 2021-12-08 | 2559.059 | 2881.084 | 340.253 | 1499.584 | 2388.21 | 2185.092 | 1924.274 | 2141.917 | 80.735 | 670.641 |
| 2021-12-09 | 2559.3 | 2882.018 | 340.517 | 1499.95 | 2392.802 | 2196.572 | 1930.162 | 2144.087 | 80.812 | 671.762 |
| 2021-12-10 | 2559.936 | 2883.084 | 340.799 | 1500.283 | 2398.296 | 2207.477 | 1935.493 | 2145.847 | 81.433 | 672.788 |
| 2021-12-11 | 2560.199 | 2883.331 | 341.202 | 1500.883 | 2400.06 | 2218.405 | 1938.512 | 2147.782 | 81.588 | 673.544 |
| 2021-12-12 | 2560.441 | 2883.715 | 341.347 | 1501.233 | 2401.003 | 2224.57 | 1940.516 | 2148.544 | 81.665 | 674.124 |
| 2021-12-13 | 2560.901 | 2883.925 | 341.528 | 1501.416 | 2405.181 | 2229.517 | 1945.122 | 2149.101 | 81.937 | 674.957 |
| 2021-12-14 | 2561.647 | 2884.453 | 341.705 | 1501.815 | 2409.72 | 2239.133 | 1950.889 | 2151.301 | 82.092 | 675.939 |
| 2021-12-15 | 2562.326 | 2885.845 | 341.951 | 1502.715 | 2416.562 | 2248.013 | 1956.473 | 2153.705 | 82.441 | 677.103 |
| 2021-12-16 | 2562.699 | 2886.42 | 342.232 | 1503.314 | 2420.329 | 2256.825 | 1961.54 | 2155.86 | 82.751 | 678.029 |
| 2021-12-17 | 2563.094 | 2887.346 | 342.232 | 1503.897 | 2425.892 | 2265.015 | 1965.874 | 2157.517 | 83.061 | 678.917 |
| 2021-12-18 | 2563.247 | 2887.99 | 342.439 | 1504.697 | 2427.793 | 2272.285 | 1970.067 | 2159.35 | 83.216 | 679.683 |
| 2021-12-19 | 2563.335 | 2888.247 | 342.723 | 1504.747 | 2428.553 | 2276.955 | 1972.059 | 2160.009 | 83.216 | 680.229 |
| 2021-12-20 | 2563.927 | 2888.635 | 343.049 | 1506.496 | 2433.215 | 2281.119 | 1976.605 | 2160.64 | 83.527 | 681.109 |
| 2021-12-21 | 2564.431 | 2888.967 | 343.277 | 1507.078 | 2439.072 | 2289.746 | 1981.635 | 2163.176 | 83.837 | 682.09 |
| 2021-12-22 | 2564.672 | 2889.649 | 343.588 | 1508.727 | 2445.425 | 2297.292 | 1986.704 | 2165.243 | 84.263 | 683.132 |
| 2021-12-23 | 2565.001 | 2890.238 | 343.857 | 1509.976 | 2455.834 | 2304.263 | 1991.362 | 2167.399 | 84.612 | 684.307 |
| 2021-12-24 | 2565.637 | 2891 | 344.134 | 1511.326 | 2457.837 | 2310.567 | 1994.743 | 2169.407 | 84.845 | 685.051 |
| 2021-12-25 | 2565.9 | 2891.145 | 344.251 | 1511.825 | 2458.378 | 2317.331 | 1997.684 | 2169.554 | 84.923 | 685.585 |
| 2021-12-26 | 2566.229 | 2891.36 | 344.477 | 1512.508 | 2458.9 | 2321.104 | 1999.104 | 2169.598 | 85.155 | 686.037 |
| 2021-12-27 | 2566.909 | 2891.663 | 344.687 | 1512.758 | 2464.478 | 2324.739 | 2003.265 | 2171.694 | 85.388 | 686.886 |
| 2021-12-28 | 2567.326 | 2892.509 | 344.904 | 1513.174 | 2471.597 | 2328.489 | 2007.954 | 2171.973 | 85.698 | 687.824 |
| 2021-12-29 | 2567.896 | 2893.056 | 345.096 | 1514.523 | 2478.422 | 2336.219 | 2013.769 | 2172.823 | 86.241 | 688.839 |
| 2021-12-30 | 2568.663 | 2893.776 | 345.254 | 1516.622 | 2482.894 | 2343.097 | 2018.368 | 2177.691 | 86.823 | 689.792 |
| 2021-12-31 | 2569.168 | 2894.173 | 345.545 | 1518.021 | 2484.88 | 2348.642 | 2022.273 | 2180.667 | 87.365 | 690.622 |
| 2022-01-01 | 2569.431 | 2894.327 | 345.749 | 1518.904 | 2485.871 | 2353.519 | 2024.753 | 2182.925 | 87.521 | 691.153 |
| 2022-01-02 | 2569.935 | 2894.486 | 345.837 | 1519.403 | 2486.796 | 2356.303 | 2026.005 | 2183.995 | 87.87 | 691.549 |
| 2022-01-03 | 2570.834 | 2894.822 | 345.926 | 1520.802 | 2492.116 | 2359.455 | 2029.798 | 2184.611 | 88.025 | 692.309 |
| 2022-01-04 | 2571.908 | 2895.668 | 346.31 | 1523.117 | 2498.832 | 2363.113 | 2034.699 | 2185.329 | 88.8 | 693.306 |
| 2022-01-05 | 2573.049 | 2895.668 | 346.543 | 1524.949 | 2505.008 | 2369.946 | 2039.921 | 2190.373 | 89.266 | 694.305 |
| 2022-01-06 | 2573.926 | 2895.668 | 346.76 | 1534.126 | 2510.79 | 2374.8 | 2044.044 | 2193.759 | 90.002 | 695.231 |
| 2022-01-07 | 2574.847 | 2896.453 | 346.964 | 1536.458 | 2517.618 | 2379.608 | 2047.309 | 2197.132 | 90.623 | 696.132 |
| 2022-01-08 | 2575.658 | 2896.991 | 347.199 | 1538.44 | 2519.763 | 2381.656 | 2049.697 | 2201.72 | 91.747 | 696.792 |
| 2022-01-09 | 2576.25 | 2898.458 | 347.304 | 1539.806 | 2521.096 | 2383.772 | 2051.086 | 2203.143 | 92.562 | 697.27 |
| 2022-01-10 | 2577.368 | 2898.995 | 347.502 | 1541.088 | 2526.74 | 2386.096 | 2054.358 | 2204.286 | 93.647 | 698.069 |
| 2022-01-11 | 2578.508 | 2899.654 | 347.502 | 1543.07 | 2533.709 | 2391.479 | 2060.194 | 2209.872 | 95.586 | 699.147 |
| 2022-01-12 | 2580.153 | 2900.28 | 348.092 | 1546.085 | 2541.201 | 2396.287 | 2065.588 | 2215.737 | 97.797 | 700.323 |
| 2022-01-13 | 2583.179 | 2901.164 | 348.161 | 1548.733 | 2546.974 | 2401.073 | 2069.872 | 2220.648 | 99.735 | 701.244 |
| 2022-01-14 | 2585.218 | 2902.253 | 348.607 | 1550.864 | 2553.979 | 2404.616 | 2074.226 | 2224.607 | 101.636 | 702.3 |
| 2022-01-15 | 2587.148 | 2903.047 | 348.832 | 1553.546 | 2556.478 | 2407.882 | 2077.209 | 2228.814 | 103.652 | 703.062 |
| 2022-01-16 | 2588.266 | 2903.486 | 349.109 | 1554.978 | 2558.175 | 2410.229 | 2078.913 | 2230.105 | 104.389 | 703.587 |
| 2022-01-17 | 2592.454 | 2904.192 | 349.331 | 1556.427 | 2561.62 | 2412.323 | 2082.979 | 2231.395 | 107.53 | 704.395 |
| 2022-01-18 | 2596.598 | 2905.711 | 349.648 | 1558.426 | 2567.321 | 2416.993 | 2088.395 | 2237.846 | 110.244 | 705.467 |
| 2022-01-19 | 2601.159 | 2907.215 | 350 | 1558.426 | 2578.766 | 2421.065 | 2092.648 | 2243.109 | 112.067 | 706.789 |
| 2022-01-20 | 2605.128 | 2908.856 | 350.504 | 1563.006 | 2586.212 | 2424.378 | 2097.286 | 2247.947 | 115.479 | 707.947 |

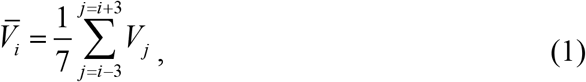

and averaged new daily numbers DCC and DDC as follows:

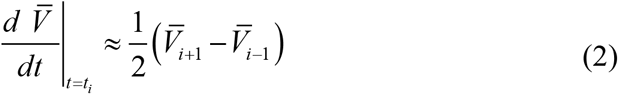

Formulas (1)- (2) can yield the averaged DCC and DDC values which are closer to the characteristics at the moments of time *t*_*i*_ in comparison with ones calculated by JHU [5] with the use of *V*_*j*_ corresponding only to previous moments of time.

Tables 5 and 6 represent the accumulated numbers of fully vaccinated people and boosters per hundred for the period of August 25, 2021 to January 20, 2022 for the same countries and regions according to [5]. Since the booster figures in Ukraine, India and South Africa for this period were negligible, they are not listed in JHU statistics [5] and in Table 6. We will use datasets from Tables 5 and 6 without any smoothing. Since JHU often changes its data sets, we have to clarify that the data in Tables 1-6 correspond to the version of the file published on January 22, 2022.

**Table 5.**
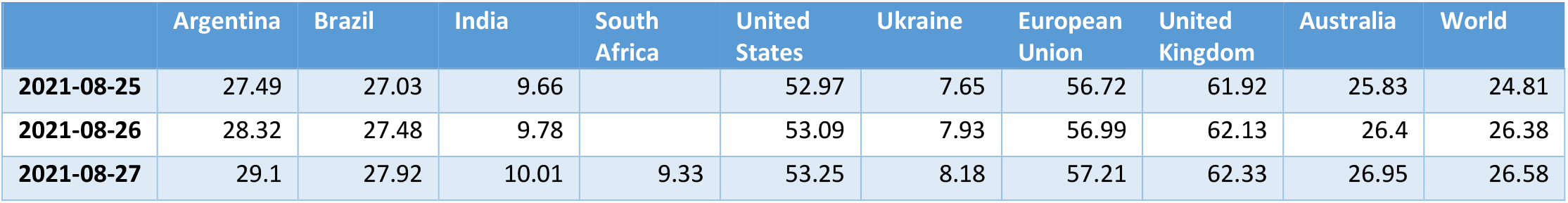
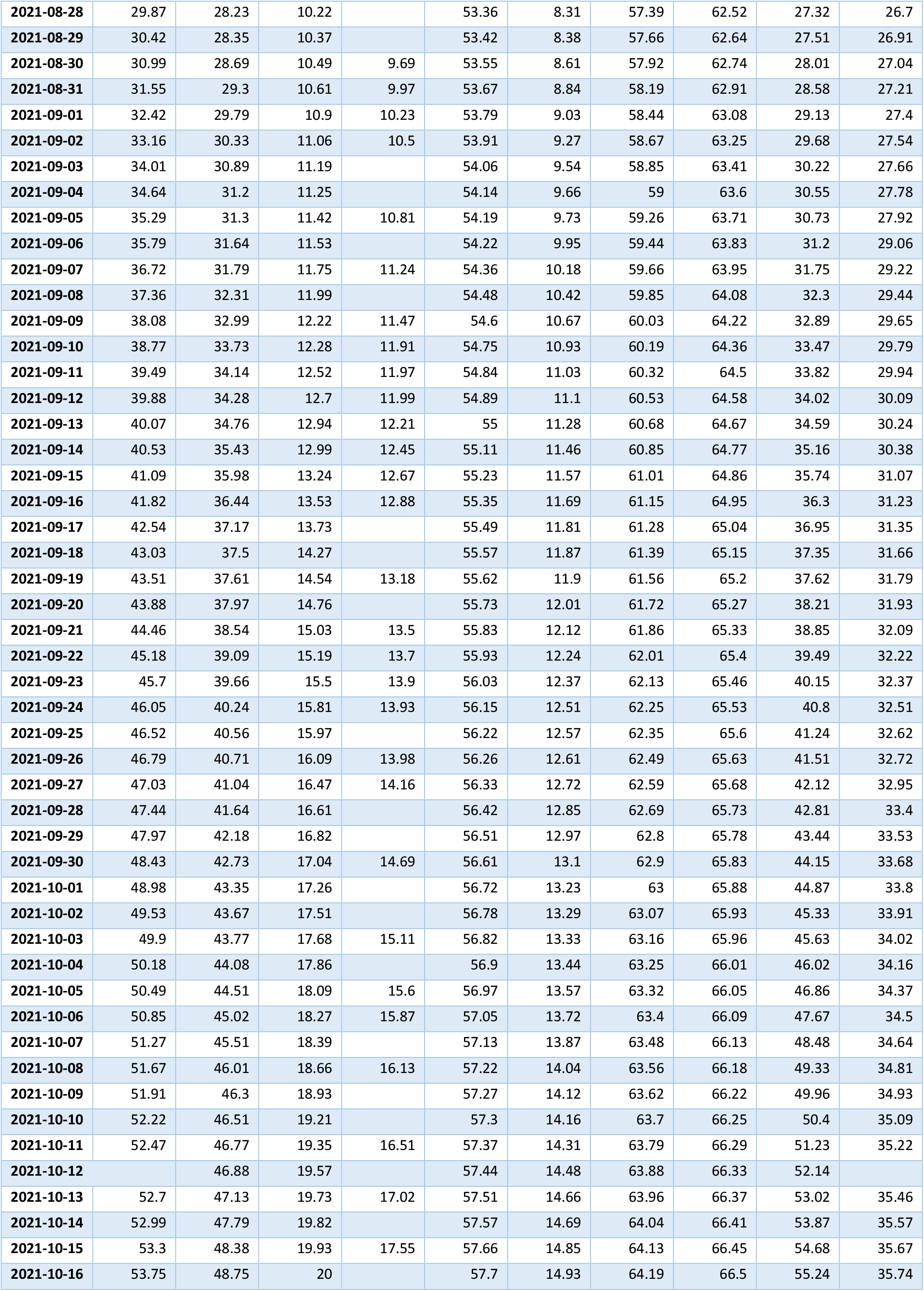
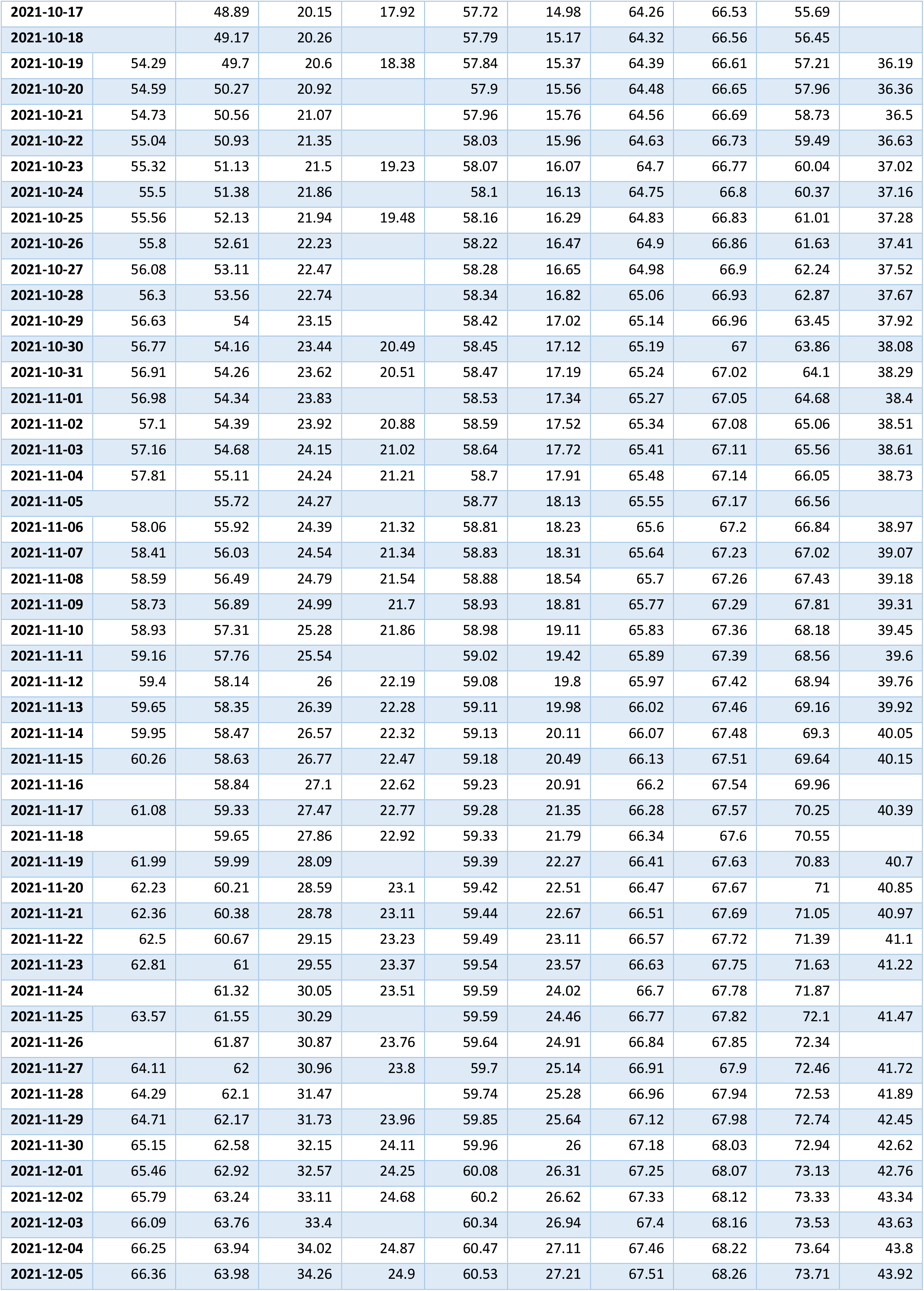
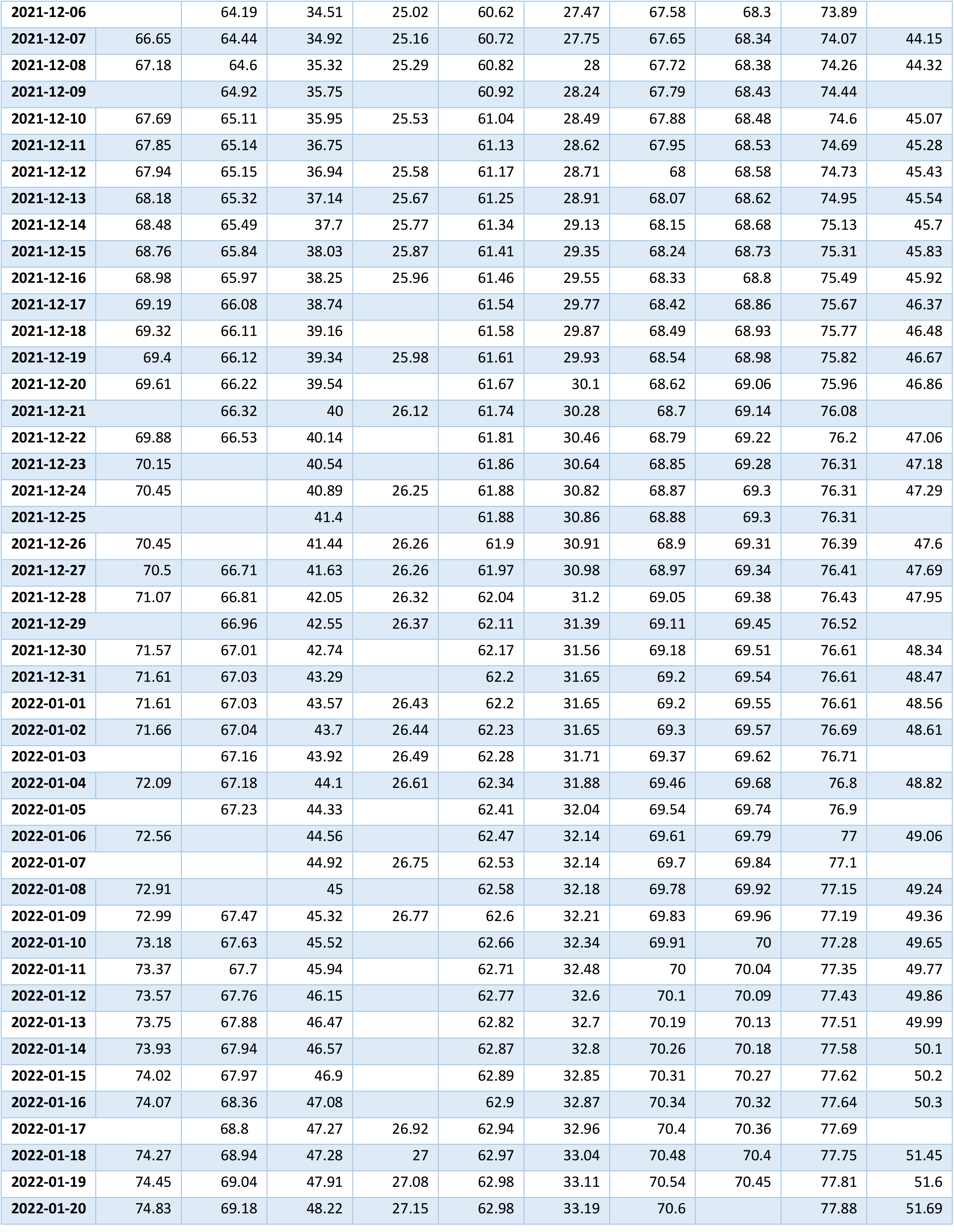
Numbers of fully vaccinated persons per hundred for the period August 25, 2021 - January 20, 2022, [5].

|  | Argentina | Brazil | India | South Africa | United States | Ukraine | European Union | United Kingdom | Australia | World |
| --- | --- | --- | --- | --- | --- | --- | --- | --- | --- | --- |
| 2021-08-25 | 27.49 | 27.03 | 9.66 |  | 52.97 | 7.65 | 56.72 | 61.92 | 25.83 | 24.81 |
| 2021-08-26 | 28.32 | 27.48 | 9.78 |  | 53.09 | 7.93 | 56.99 | 62.13 | 26.4 | 26.38 |
| 2021-08-27 | 29.1 | 27.92 | 10.01 | 9.33 | 53.25 | 8.18 | 57.21 | 62.33 | 26.95 | 26.58 |
| 2021-08-28 | 29.87 | 28.23 | 10.22 |  | 53.36 | 8.31 | 57.39 | 62.52 | 27.32 | 26.7 |
| 2021-08-29 | 30.42 | 28.35 | 10.37 |  | 53.42 | 8.38 | 57.66 | 62.64 | 27.51 | 26.91 |
| 2021-08-30 | 30.99 | 28.69 | 10.49 | 9.69 | 53.55 | 8.61 | 57.92 | 62.74 | 28.01 | 27.04 |
| 2021-08-31 | 31.55 | 29.3 | 10.61 | 9.97 | 53.67 | 8.84 | 58.19 | 62.91 | 28.58 | 27.21 |
| 2021-09-01 | 32.42 | 29.79 | 10.9 | 10.23 | 53.79 | 9.03 | 58.44 | 63.08 | 29.13 | 27.4 |
| 2021-09-02 | 33.16 | 30.33 | 11.06 | 10.5 | 53.91 | 9.27 | 58.67 | 63.25 | 29.68 | 27.54 |
| 2021-09-03 | 34.01 | 30.89 | 11.19 |  | 54.06 | 9.54 | 58.85 | 63.41 | 30.22 | 27.66 |
| 2021-09-04 | 34.64 | 31.2 | 11.25 |  | 54.14 | 9.66 | 59 | 63.6 | 30.55 | 27.78 |
| 2021-09-05 | 35.29 | 31.3 | 11.42 | 10.81 | 54.19 | 9.73 | 59.26 | 63.71 | 30.73 | 27.92 |
| 2021-09-06 | 35.79 | 31.64 | 11.53 |  | 54.22 | 9.95 | 59.44 | 63.83 | 31.2 | 29.06 |
| 2021-09-07 | 36.72 | 31.79 | 11.75 | 11.24 | 54.36 | 10.18 | 59.66 | 63.95 | 31.75 | 29.22 |
| 2021-09-08 | 37.36 | 32.31 | 11.99 |  | 54.48 | 10.42 | 59.85 | 64.08 | 32.3 | 29.44 |
| 2021-09-09 | 38.08 | 32.99 | 12.22 | 11.47 | 54.6 | 10.67 | 60.03 | 64.22 | 32.89 | 29.65 |
| 2021-09-10 | 38.77 | 33.73 | 12.28 | 11.91 | 54.75 | 10.93 | 60.19 | 64.36 | 33.47 | 29.79 |
| 2021-09-11 | 39.49 | 34.14 | 12.52 | 11.97 | 54.84 | 11.03 | 60.32 | 64.5 | 33.82 | 29.94 |
| 2021-09-12 | 39.88 | 34.28 | 12.7 | 11.99 | 54.89 | 11.1 | 60.53 | 64.58 | 34.02 | 30.09 |
| 2021-09-13 | 40.07 | 34.76 | 12.94 | 12.21 | 55 | 11.28 | 60.68 | 64.67 | 34.59 | 30.24 |
| 2021-09-14 | 40.53 | 35.43 | 12.99 | 12.45 | 55.11 | 11.46 | 60.85 | 64.77 | 35.16 | 30.38 |
| 2021-09-15 | 41.09 | 35.98 | 13.24 | 12.67 | 55.23 | 11.57 | 61.01 | 64.86 | 35.74 | 31.07 |
| 2021-09-16 | 41.82 | 36.44 | 13.53 | 12.88 | 55.35 | 11.69 | 61.15 | 64.95 | 36.3 | 31.23 |
| 2021-09-17 | 42.54 | 37.17 | 13.73 |  | 55.49 | 11.81 | 61.28 | 65.04 | 36.95 | 31.35 |
| 2021-09-18 | 43.03 | 37.5 | 14.27 |  | 55.57 | 11.87 | 61.39 | 65.15 | 37.35 | 31.66 |
| 2021-09-19 | 43.51 | 37.61 | 14.54 | 13.18 | 55.62 | 11.9 | 61.56 | 65.2 | 37.62 | 31.79 |
| 2021-09-20 | 43.88 | 37.97 | 14.76 |  | 55.73 | 12.01 | 61.72 | 65.27 | 38.21 | 31.93 |
| 2021-09-21 | 44.46 | 38.54 | 15.03 | 13.5 | 55.83 | 12.12 | 61.86 | 65.33 | 38.85 | 32.09 |
| 2021-09-22 | 45.18 | 39.09 | 15.19 | 13.7 | 55.93 | 12.24 | 62.01 | 65.4 | 39.49 | 32.22 |
| 2021-09-23 | 45.7 | 39.66 | 15.5 | 13.9 | 56.03 | 12.37 | 62.13 | 65.46 | 40.15 | 32.37 |
| 2021-09-24 | 46.05 | 40.24 | 15.81 | 13.93 | 56.15 | 12.51 | 62.25 | 65.53 | 40.8 | 32.51 |
| 2021-09-25 | 46.52 | 40.56 | 15.97 |  | 56.22 | 12.57 | 62.35 | 65.6 | 41.24 | 32.62 |
| 2021-09-26 | 46.79 | 40.71 | 16.09 | 13.98 | 56.26 | 12.61 | 62.49 | 65.63 | 41.51 | 32.72 |
| 2021-09-27 | 47.03 | 41.04 | 16.47 | 14.16 | 56.33 | 12.72 | 62.59 | 65.68 | 42.12 | 32.95 |
| 2021-09-28 | 47.44 | 41.64 | 16.61 |  | 56.42 | 12.85 | 62.69 | 65.73 | 42.81 | 33.4 |
| 2021-09-29 | 47.97 | 42.18 | 16.82 |  | 56.51 | 12.97 | 62.8 | 65.78 | 43.44 | 33.53 |
| 2021-09-30 | 48.43 | 42.73 | 17.04 | 14.69 | 56.61 | 13.1 | 62.9 | 65.83 | 44.15 | 33.68 |
| 2021-10-01 | 48.98 | 43.35 | 17.26 |  | 56.72 | 13.23 | 63 | 65.88 | 44.87 | 33.8 |
| 2021-10-02 | 49.53 | 43.67 | 17.51 |  | 56.78 | 13.29 | 63.07 | 65.93 | 45.33 | 33.91 |
| 2021-10-03 | 49.9 | 43.77 | 17.68 | 15.11 | 56.82 | 13.33 | 63.16 | 65.96 | 45.63 | 34.02 |
| 2021-10-04 | 50.18 | 44.08 | 17.86 |  | 56.9 | 13.44 | 63.25 | 66.01 | 46.02 | 34.16 |
| 2021-10-05 | 50.49 | 44.51 | 18.09 | 15.6 | 56.97 | 13.57 | 63.32 | 66.05 | 46.86 | 34.37 |
| 2021-10-06 | 50.85 | 45.02 | 18.27 | 15.87 | 57.05 | 13.72 | 63.4 | 66.09 | 47.67 | 34.5 |
| 2021-10-07 | 51.27 | 45.51 | 18.39 |  | 57.13 | 13.87 | 63.48 | 66.13 | 48.48 | 34.64 |
| 2021-10-08 | 51.67 | 46.01 | 18.66 | 16.13 | 57.22 | 14.04 | 63.56 | 66.18 | 49.33 | 34.81 |
| 2021-10-09 | 51.91 | 46.3 | 18.93 |  | 57.27 | 14.12 | 63.62 | 66.22 | 49.96 | 34.93 |
| 2021-10-10 | 52.22 | 46.51 | 19.21 |  | 57.3 | 14.16 | 63.7 | 66.25 | 50.4 | 35.09 |
| 2021-10-11 | 52.47 | 46.77 | 19.35 | 16.51 | 57.37 | 14.31 | 63.79 | 66.29 | 51.23 | 35.22 |
| 2021-10-12 |  | 46.88 | 19.57 |  | 57.44 | 14.48 | 63.88 | 66.33 | 52.14 |  |
| 2021-10-13 | 52.7 | 47.13 | 19.73 | 17.02 | 57.51 | 14.66 | 63.96 | 66.37 | 53.02 | 35.46 |
| 2021-10-14 | 52.99 | 47.79 | 19.82 |  | 57.57 | 14.69 | 64.04 | 66.41 | 53.87 | 35.57 |
| 2021-10-15 | 53.3 | 48.38 | 19.93 | 17.55 | 57.66 | 14.85 | 64.13 | 66.45 | 54.68 | 35.67 |
| 2021-10-16 | 53.75 | 48.75 | 20 |  | 57.7 | 14.93 | 64.19 | 66.5 | 55.24 | 35.74 |
| 2021-10-17 |  | 48.89 | 20.15 | 17.92 | 57.72 | 14.98 | 64.26 | 66.53 | 55.69 |  |
| 2021-10-18 |  | 49.17 | 20.26 |  | 57.79 | 15.17 | 64.32 | 66.56 | 56.45 |  |
| 2021-10-19 | 54.29 | 49.7 | 20.6 | 18.38 | 57.84 | 15.37 | 64.39 | 66.61 | 57.21 | 36.19 |
| 2021-10-20 | 54.59 | 50.27 | 20.92 |  | 57.9 | 15.56 | 64.48 | 66.65 | 57.96 | 36.36 |
| 2021-10-21 | 54.73 | 50.56 | 21.07 |  | 57.96 | 15.76 | 64.56 | 66.69 | 58.73 | 36.5 |
| 2021-10-22 | 55.04 | 50.93 | 21.35 |  | 58.03 | 15.96 | 64.63 | 66.73 | 59.49 | 36.63 |
| 2021-10-23 | 55.32 | 51.13 | 21.5 | 19.23 | 58.07 | 16.07 | 64.7 | 66.77 | 60.04 | 37.02 |
| 2021-10-24 | 55.5 | 51.38 | 21.86 |  | 58.1 | 16.13 | 64.75 | 66.8 | 60.37 | 37.16 |
| 2021-10-25 | 55.56 | 52.13 | 21.94 | 19.48 | 58.16 | 16.29 | 64.83 | 66.83 | 61.01 | 37.28 |
| 2021-10-26 | 55.8 | 52.61 | 22.23 |  | 58.22 | 16.47 | 64.9 | 66.86 | 61.63 | 37.41 |
| 2021-10-27 | 56.08 | 53.11 | 22.47 |  | 58.28 | 16.65 | 64.98 | 66.9 | 62.24 | 37.52 |
| 2021-10-28 | 56.3 | 53.56 | 22.74 |  | 58.34 | 16.82 | 65.06 | 66.93 | 62.87 | 37.67 |
| 2021-10-29 | 56.63 | 54 | 23.15 |  | 58.42 | 17.02 | 65.14 | 66.96 | 63.45 | 37.92 |
| 2021-10-30 | 56.77 | 54.16 | 23.44 | 20.49 | 58.45 | 17.12 | 65.19 | 67 | 63.86 | 38.08 |
| 2021-10-31 | 56.91 | 54.26 | 23.62 | 20.51 | 58.47 | 17.19 | 65.24 | 67.02 | 64.1 | 38.29 |
| 2021-11-01 | 56.98 | 54.34 | 23.83 |  | 58.53 | 17.34 | 65.27 | 67.05 | 64.68 | 38.4 |
| 2021-11-02 | 57.1 | 54.39 | 23.92 | 20.88 | 58.59 | 17.52 | 65.34 | 67.08 | 65.06 | 38.51 |
| 2021-11-03 | 57.16 | 54.68 | 24.15 | 21.02 | 58.64 | 17.72 | 65.41 | 67.11 | 65.56 | 38.61 |
| 2021-11-04 | 57.81 | 55.11 | 24.24 | 21.21 | 58.7 | 17.91 | 65.48 | 67.14 | 66.05 | 38.73 |
| 2021-11-05 |  | 55.72 | 24.27 |  | 58.77 | 18.13 | 65.55 | 67.17 | 66.56 |  |
| 2021-11-06 | 58.06 | 55.92 | 24.39 | 21.32 | 58.81 | 18.23 | 65.6 | 67.2 | 66.84 | 38.97 |
| 2021-11-07 | 58.41 | 56.03 | 24.54 | 21.34 | 58.83 | 18.31 | 65.64 | 67.23 | 67.02 | 39.07 |
| 2021-11-08 | 58.59 | 56.49 | 24.79 | 21.54 | 58.88 | 18.54 | 65.7 | 67.26 | 67.43 | 39.18 |
| 2021-11-09 | 58.73 | 56.89 | 24.99 | 21.7 | 58.93 | 18.81 | 65.77 | 67.29 | 67.81 | 39.31 |
| 2021-11-10 | 58.93 | 57.31 | 25.28 | 21.86 | 58.98 | 19.11 | 65.83 | 67.36 | 68.18 | 39.45 |
| 2021-11-11 | 59.16 | 57.76 | 25.54 |  | 59.02 | 19.42 | 65.89 | 67.39 | 68.56 | 39.6 |
| 2021-11-12 | 59.4 | 58.14 | 26 | 22.19 | 59.08 | 19.8 | 65.97 | 67.42 | 68.94 | 39.76 |
| 2021-11-13 | 59.65 | 58.35 | 26.39 | 22.28 | 59.11 | 19.98 | 66.02 | 67.46 | 69.16 | 39.92 |
| 2021-11-14 | 59.95 | 58.47 | 26.57 | 22.32 | 59.13 | 20.11 | 66.07 | 67.48 | 69.3 | 40.05 |
| 2021-11-15 | 60.26 | 58.63 | 26.77 | 22.47 | 59.18 | 20.49 | 66.13 | 67.51 | 69.64 | 40.15 |
| 2021-11-16 |  | 58.84 | 27.1 | 22.62 | 59.23 | 20.91 | 66.2 | 67.54 | 69.96 |  |
| 2021-11-17 | 61.08 | 59.33 | 27.47 | 22.77 | 59.28 | 21.35 | 66.28 | 67.57 | 70.25 | 40.39 |
| 2021-11-18 |  | 59.65 | 27.86 | 22.92 | 59.33 | 21.79 | 66.34 | 67.6 | 70.55 |  |
| 2021-11-19 | 61.99 | 59.99 | 28.09 |  | 59.39 | 22.27 | 66.41 | 67.63 | 70.83 | 40.7 |
| 2021-11-20 | 62.23 | 60.21 | 28.59 | 23.1 | 59.42 | 22.51 | 66.47 | 67.67 | 71 | 40.85 |
| 2021-11-21 | 62.36 | 60.38 | 28.78 | 23.11 | 59.44 | 22.67 | 66.51 | 67.69 | 71.05 | 40.97 |
| 2021-11-22 | 62.5 | 60.67 | 29.15 | 23.23 | 59.49 | 23.11 | 66.57 | 67.72 | 71.39 | 41.1 |
| 2021-11-23 | 62.81 | 61 | 29.55 | 23.37 | 59.54 | 23.57 | 66.63 | 67.75 | 71.63 | 41.22 |
| 2021-11-24 |  | 61.32 | 30.05 | 23.51 | 59.59 | 24.02 | 66.7 | 67.78 | 71.87 |  |
| 2021-11-25 | 63.57 | 61.55 | 30.29 |  | 59.59 | 24.46 | 66.77 | 67.82 | 72.1 | 41.47 |
| 2021-11-26 |  | 61.87 | 30.87 | 23.76 | 59.64 | 24.91 | 66.84 | 67.85 | 72.34 |  |
| 2021-11-27 | 64.11 | 62 | 30.96 | 23.8 | 59.7 | 25.14 | 66.91 | 67.9 | 72.46 | 41.72 |
| 2021-11-28 | 64.29 | 62.1 | 31.47 |  | 59.74 | 25.28 | 66.96 | 67.94 | 72.53 | 41.89 |
| 2021-11-29 | 64.71 | 62.17 | 31.73 | 23.96 | 59.85 | 25.64 | 67.12 | 67.98 | 72.74 | 42.45 |
| 2021-11-30 | 65.15 | 62.58 | 32.15 | 24.11 | 59.96 | 26 | 67.18 | 68.03 | 72.94 | 42.62 |
| 2021-12-01 | 65.46 | 62.92 | 32.57 | 24.25 | 60.08 | 26.31 | 67.25 | 68.07 | 73.13 | 42.76 |
| 2021-12-02 | 65.79 | 63.24 | 33.11 | 24.68 | 60.2 | 26.62 | 67.33 | 68.12 | 73.33 | 43.34 |
| 2021-12-03 | 66.09 | 63.76 | 33.4 |  | 60.34 | 26.94 | 67.4 | 68.16 | 73.53 | 43.63 |
| 2021-12-04 | 66.25 | 63.94 | 34.02 | 24.87 | 60.47 | 27.11 | 67.46 | 68.22 | 73.64 | 43.8 |
| 2021-12-05 | 66.36 | 63.98 | 34.26 | 24.9 | 60.53 | 27.21 | 67.51 | 68.26 | 73.71 | 43.92 |
| 2021-12-06 |  | 64.19 | 34.51 | 25.02 | 60.62 | 27.47 | 67.58 | 68.3 | 73.89 |  |
| 2021-12-07 | 66.65 | 64.44 | 34.92 | 25.16 | 60.72 | 27.75 | 67.65 | 68.34 | 74.07 | 44.15 |
| 2021-12-08 | 67.18 | 64.6 | 35.32 | 25.29 | 60.82 | 28 | 67.72 | 68.38 | 74.26 | 44.32 |
| 2021-12-09 |  | 64.92 | 35.75 |  | 60.92 | 28.24 | 67.79 | 68.43 | 74.44 |  |
| 2021-12-10 | 67.69 | 65.11 | 35.95 | 25.53 | 61.04 | 28.49 | 67.88 | 68.48 | 74.6 | 45.07 |
| 2021-12-11 | 67.85 | 65.14 | 36.75 |  | 61.13 | 28.62 | 67.95 | 68.53 | 74.69 | 45.28 |
| 2021-12-12 | 67.94 | 65.15 | 36.94 | 25.58 | 61.17 | 28.71 | 68 | 68.58 | 74.73 | 45.43 |
| 2021-12-13 | 68.18 | 65.32 | 37.14 | 25.67 | 61.25 | 28.91 | 68.07 | 68.62 | 74.95 | 45.54 |
| 2021-12-14 | 68.48 | 65.49 | 37.7 | 25.77 | 61.34 | 29.13 | 68.15 | 68.68 | 75.13 | 45.7 |
| 2021-12-15 | 68.76 | 65.84 | 38.03 | 25.87 | 61.41 | 29.35 | 68.24 | 68.73 | 75.31 | 45.83 |
| 2021-12-16 | 68.98 | 65.97 | 38.25 | 25.96 | 61.46 | 29.55 | 68.33 | 68.8 | 75.49 | 45.92 |
| 2021-12-17 | 69.19 | 66.08 | 38.74 |  | 61.54 | 29.77 | 68.42 | 68.86 | 75.67 | 46.37 |
| 2021-12-18 | 69.32 | 66.11 | 39.16 |  | 61.58 | 29.87 | 68.49 | 68.93 | 75.77 | 46.48 |
| 2021-12-19 | 69.4 | 66.12 | 39.34 | 25.98 | 61.61 | 29.93 | 68.54 | 68.98 | 75.82 | 46.67 |
| 2021-12-20 | 69.61 | 66.22 | 39.54 |  | 61.67 | 30.1 | 68.62 | 69.06 | 75.96 | 46.86 |
| 2021-12-21 |  | 66.32 | 40 | 26.12 | 61.74 | 30.28 | 68.7 | 69.14 | 76.08 |  |
| 2021-12-22 | 69.88 | 66.53 | 40.14 |  | 61.81 | 30.46 | 68.79 | 69.22 | 76.2 | 47.06 |
| 2021-12-23 | 70.15 |  | 40.54 |  | 61.86 | 30.64 | 68.85 | 69.28 | 76.31 | 47.18 |
| 2021-12-24 | 70.45 |  | 40.89 | 26.25 | 61.88 | 30.82 | 68.87 | 69.3 | 76.31 | 47.29 |
| 2021-12-25 |  |  | 41.4 |  | 61.88 | 30.86 | 68.88 | 69.3 | 76.31 |  |
| 2021-12-26 | 70.45 |  | 41.44 | 26.26 | 61.9 | 30.91 | 68.9 | 69.31 | 76.39 | 47.6 |
| 2021-12-27 | 70.5 | 66.71 | 41.63 | 26.26 | 61.97 | 30.98 | 68.97 | 69.34 | 76.41 | 47.69 |
| 2021-12-28 | 71.07 | 66.81 | 42.05 | 26.32 | 62.04 | 31.2 | 69.05 | 69.38 | 76.43 | 47.95 |
| 2021-12-29 |  | 66.96 | 42.55 | 26.37 | 62.11 | 31.39 | 69.11 | 69.45 | 76.52 |  |
| 2021-12-30 | 71.57 | 67.01 | 42.74 |  | 62.17 | 31.56 | 69.18 | 69.51 | 76.61 | 48.34 |
| 2021-12-31 | 71.61 | 67.03 | 43.29 |  | 62.2 | 31.65 | 69.2 | 69.54 | 76.61 | 48.47 |
| 2022-01-01 | 71.61 | 67.03 | 43.57 | 26.43 | 62.2 | 31.65 | 69.2 | 69.55 | 76.61 | 48.56 |
| 2022-01-02 | 71.66 | 67.04 | 43.7 | 26.44 | 62.23 | 31.65 | 69.3 | 69.57 | 76.69 | 48.61 |
| 2022-01-03 |  | 67.16 | 43.92 | 26.49 | 62.28 | 31.71 | 69.37 | 69.62 | 76.71 |  |
| 2022-01-04 | 72.09 | 67.18 | 44.1 | 26.61 | 62.34 | 31.88 | 69.46 | 69.68 | 76.8 | 48.82 |
| 2022-01-05 |  | 67.23 | 44.33 |  | 62.41 | 32.04 | 69.54 | 69.74 | 76.9 |  |
| 2022-01-06 | 72.56 |  | 44.56 |  | 62.47 | 32.14 | 69.61 | 69.79 | 77 | 49.06 |
| 2022-01-07 |  |  | 44.92 | 26.75 | 62.53 | 32.14 | 69.7 | 69.84 | 77.1 |  |
| 2022-01-08 | 72.91 |  | 45 |  | 62.58 | 32.18 | 69.78 | 69.92 | 77.15 | 49.24 |
| 2022-01-09 | 72.99 | 67.47 | 45.32 | 26.77 | 62.6 | 32.21 | 69.83 | 69.96 | 77.19 | 49.36 |
| 2022-01-10 | 73.18 | 67.63 | 45.52 |  | 62.66 | 32.34 | 69.91 | 70 | 77.28 | 49.65 |
| 2022-01-11 | 73.37 | 67.7 | 45.94 |  | 62.71 | 32.48 | 70 | 70.04 | 77.35 | 49.77 |
| 2022-01-12 | 73.57 | 67.76 | 46.15 |  | 62.77 | 32.6 | 70.1 | 70.09 | 77.43 | 49.86 |
| 2022-01-13 | 73.75 | 67.88 | 46.47 |  | 62.82 | 32.7 | 70.19 | 70.13 | 77.51 | 49.99 |
| 2022-01-14 | 73.93 | 67.94 | 46.57 |  | 62.87 | 32.8 | 70.26 | 70.18 | 77.58 | 50.1 |
| 2022-01-15 | 74.02 | 67.97 | 46.9 |  | 62.89 | 32.85 | 70.31 | 70.27 | 77.62 | 50.2 |
| 2022-01-16 | 74.07 | 68.36 | 47.08 |  | 62.9 | 32.87 | 70.34 | 70.32 | 77.64 | 50.3 |
| 2022-01-17 |  | 68.8 | 47.27 | 26.92 | 62.94 | 32.96 | 70.4 | 70.36 | 77.69 |  |
| 2022-01-18 | 74.27 | 68.94 | 47.28 | 27 | 62.97 | 33.04 | 70.48 | 70.4 | 77.75 | 51.45 |
| 2022-01-19 | 74.45 | 69.04 | 47.91 | 27.08 | 62.98 | 33.11 | 70.54 | 70.45 | 77.81 | 51.6 |
| 2022-01-20 | 74.83 | 69.18 | 48.22 | 27.15 | 62.98 | 33.19 | 70.6 |  | 77.88 | 51.69 |

**Table 6.** Numbers of boosters per hundred in different regions for the period August 25. 2021 - January 20, 2022. [5].

|  | Argentina | Brazil | United States | European Union | United Kingdom | Australia | World |
| --- | --- | --- | --- | --- | --- | --- | --- |
| 2021-08-25 |  |  | 0.3 | 0.04 |  |  | 0.21 |
| 2021-08-26 |  |  | 0.33 | 0.04 |  |  | 0.22 |
| 2021-08-27 |  |  | 0.37 | 0.04 |  |  | 0.22 |
| 2021-08-28 |  |  | 0.39 | 0.04 |  |  | 0.22 |
| 2021-08-29 |  |  | 0.4 | 0.04 |  |  | 0.23 |
| 2021-08-30 |  |  | 0.43 | 0.04 |  |  | 0.23 |
| 2021-08-31 |  |  | 0.46 | 0.12 |  |  | 0.24 |
| 2021-09-01 |  |  | 0.49 | 0.13 |  |  | 0.25 |
| 2021-09-02 |  | 0 | 0.53 | 0.15 |  |  | 0.26 |
| 2021-09-03 |  | 0 | 0.56 | 0.16 |  |  | 0.27 |
| 2021-09-04 |  | 0 | 0.58 | 0.16 |  |  | 0.27 |
| 2021-09-05 |  | 0 | 0.58 | 0.17 |  |  | 0.27 |
| 2021-09-06 |  | 0.01 | 0.59 | 0.19 |  |  | 0.28 |
| 2021-09-07 |  | 0.01 | 0.62 | 0.2 |  |  | 0.28 |
| 2021-09-08 |  | 0.01 | 0.64 | 0.22 |  |  | 0.29 |
| 2021-09-09 |  | 0.02 | 0.67 | 0.24 |  |  | 0.3 |
| 2021-09-10 |  | 0.02 | 0.7 | 0.26 |  |  | 0.3 |
| 2021-09-11 |  | 0.03 | 0.72 | 0.26 |  |  | 0.31 |
| 2021-09-12 |  | 0.03 | 0.73 | 0.27 |  |  | 0.31 |
| 2021-09-13 |  | 0.04 | 0.75 | 0.29 |  |  | 0.31 |
| 2021-09-14 |  | 0.06 | 0.77 | 0.32 |  |  | 0.32 |
| 2021-09-15 |  | 0.09 | 0.79 | 0.35 |  |  | 0.33 |
| 2021-09-16 |  | 0.1 | 0.81 | 0.38 |  |  | 0.33 |
| 2021-09-17 |  | 0.12 | 0.84 | 0.4 |  |  | 0.34 |
| 2021-09-18 |  | 0.12 | 0.86 | 0.41 |  |  | 0.34 |
| 2021-09-19 |  | 0.12 | 0.86 | 0.42 |  |  | 0.35 |
| 2021-09-20 |  | 0.16 | 0.89 | 0.44 |  |  | 0.35 |
| 2021-09-21 |  | 0.18 | 0.91 | 0.48 |  |  | 0.36 |
| 2021-09-22 |  | 0.2 | 0.94 | 0.51 |  |  | 0.37 |
| 2021-09-23 |  | 0.21 | 0.97 | 0.55 |  |  | 0.37 |
| 2021-09-24 |  | 0.24 | 1.05 | 0.58 |  |  | 0.39 |
| 2021-09-25 |  | 0.25 | 1.12 | 0.59 |  |  | 0.4 |
| 2021-09-26 |  | 0.26 | 1.16 | 0.6 |  |  | 0.4 |
| 2021-09-27 |  | 0.38 | 1.31 | 0.63 |  |  | 0.42 |
| 2021-09-28 |  | 0.41 | 1.47 | 0.67 |  |  | 0.43 |
| 2021-09-29 |  | 0.44 | 1.63 | 0.71 |  |  | 0.44 |
| 2021-09-30 |  | 0.51 | 1.79 | 0.75 | 1.27 |  | 0.47 |
| 2021-10-01 |  | 0.61 | 1.97 | 0.78 | 1.5 |  | 0.49 |
| 2021-10-02 |  | 0.66 | 2.05 | 0.8 | 1.78 |  | 0.5 |
| 2021-10-03 |  | 0.68 | 2.08 | 0.8 | 1.91 |  | 0.5 |
| 2021-10-04 |  | 0.77 | 2.23 | 0.85 | 2.1 |  | 0.52 |
| 2021-10-05 |  | 0.88 | 2.38 | 0.95 | 2.33 |  | 0.55 |
| 2021-10-06 |  | 1.01 | 2.53 | 1.01 | 2.56 |  | 0.57 |
| 2021-10-07 |  | 1.17 | 2.68 | 1.08 | 2.82 | 0 | 0.59 |
| 2021-10-08 |  | 1.32 | 2.85 | 1.13 | 3.08 | 0 | 0.61 |
| 2021-10-09 |  | 1.42 | 2.91 | 1.15 | 3.45 | 0 | 0.63 |
| 2021-10-10 |  | 1.46 | 2.95 | 1.18 | 3.58 | 0 | 0.63 |
| 2021-10-11 |  | 1.53 | 3.07 | 1.22 | 3.77 | 0 | 0.65 |
| 2021-10-12 |  | 1.55 | 3.21 | 1.28 | 4.01 | 0 | 0.66 |
| 2021-10-13 |  | 1.66 | 3.34 | 1.36 | 4.68 | 0.01 | 0.69 |
| 2021-10-14 |  | 1.75 | 3.48 | 1.43 | 5.03 | 0.01 | 0.71 |
| 2021-10-15 |  | 1.91 | 3.63 | 1.49 | 5.36 | 0.01 | 0.73 |
| 2021-10-16 |  | 2.02 | 3.69 | 1.52 | 5.81 | 0.01 | 0.75 |
| 2021-10-17 |  | 2.07 | 3.72 | 1.54 | 5.98 | 0.01 | 0.75 |
| 2021-10-18 |  | 2.26 | 3.84 | 1.6 | 6.24 | 0.01 | 0.77 |
| 2021-10-19 |  | 2.4 | 3.97 | 1.67 | 6.58 | 0.01 | 0.8 |
| 2021-10-20 |  | 2.58 | 4.1 | 1.77 | 6.96 | 0.02 | 0.82 |
| 2021-10-21 |  | 2.7 | 4.23 | 1.85 | 7.85 | 0.02 | 0.85 |
| 2021-10-22 |  | 2.86 | 4.48 | 1.93 | 8.29 | 0.02 | 0.88 |
| 2021-10-23 |  | 2.91 | 4.63 | 1.97 | 8.84 | 0.03 | 0.9 |
| 2021-10-24 |  | 2.96 | 4.72 | 1.99 | 9.09 | 0.03 | 0.91 |
| 2021-10-25 |  | 3.21 | 5.01 | 2.27 | 9.44 | 0.03 | 0.95 |
| 2021-10-26 |  | 3.43 | 5.33 | 2.37 | 9.83 | 0.03 | 0.99 |
| 2021-10-27 |  | 3.61 | 5.65 | 2.55 | 10.26 | 0.04 | 1.03 |
| 2021-10-28 |  | 3.79 | 5.98 | 2.69 | 10.69 | 0.04 | 1.07 |
| 2021-10-29 |  | 3.99 | 6.34 | 2.79 | 11.11 | 0.05 | 1.1 |
| 2021-10-30 |  | 4.06 | 6.49 | 2.86 | 11.66 | 0.07 | 1.12 |
| 2021-10-31 |  | 4.11 | 6.57 | 2.89 | 11.9 | 0.08 | 1.13 |
| 2021-11-01 |  | 4.15 | 6.86 | 2.95 | 12.25 | 0.18 | 1.16 |
| 2021-11-02 |  | 4.18 | 7.16 | 3.08 | 12.69 | 0.28 | 1.2 |
| 2021-11-03 |  | 4.33 | 7.47 | 3.25 | 13.21 | 0.39 | 1.24 |
| 2021-11-04 |  | 4.45 | 7.79 | 3.44 | 13.7 | 0.49 | 1.28 |
| 2021-11-05 |  | 4.65 | 8.15 | 3.58 | 14.18 | 0.58 | 1.8 |
| 2021-11-06 |  | 4.74 | 8.31 | 3.65 | 14.8 | 0.67 | 1.82 |
| 2021-11-07 |  | 4.75 | 8.38 | 3.73 | 15.1 | 0.69 | 1.83 |
| 2021-11-08 |  | 4.93 | 8.62 | 3.9 | 15.51 | 0.74 | 1.88 |
| 2021-11-09 |  | 5.09 | 8.9 | 4.14 | 16.01 | 0.8 | 1.92 |
| 2021-11-10 |  | 5.25 | 9.2 | 4.38 | 16.79 | 0.86 | 1.97 |
| 2021-11-11 |  | 5.41 | 9.46 | 4.61 | 17.33 | 0.92 | 2.01 |
| 2021-11-12 |  | 5.58 | 9.8 | 4.8 | 17.86 | 0.97 | 2.2 |
| 2021-11-13 |  | 5.66 | 9.96 | 4.89 | 18.54 | 1 | 2.23 |
| 2021-11-14 |  | 5.68 | 10.04 | 5 | 18.86 | 1.01 | 2.24 |
| 2021-11-15 | 1.96 | 5.77 | 10.31 | 5.25 | 19.28 | 1.05 | 2.29 |
| 2021-11-16 |  | 5.86 | 10.61 | 5.52 | 19.79 | 1.1 | 2.34 |
| 2021-11-17 | 2.29 | 6.02 | 10.93 | 5.86 | 20.35 | 1.16 | 2.39 |
| 2021-11-18 |  | 6.2 | 11.25 | 6.19 | 20.92 | 1.21 | 2.45 |
| 2021-11-19 | 2.64 | 6.39 | 11.63 | 6.47 | 21.45 | 1.26 | 2.71 |
| 2021-11-20 | 2.77 | 6.49 | 11.83 | 6.62 | 22.13 | 1.29 | 2.74 |
| 2021-11-21 | 2.84 | 6.56 | 11.94 | 6.76 | 22.48 | 1.29 | 2.76 |
| 2021-11-22 | 2.91 | 6.73 | 12.27 | 7.13 | 22.93 | 1.34 | 2.82 |
| 2021-11-23 | 3.11 | 6.89 | 12.6 | 7.52 | 23.46 | 1.4 | 2.88 |
| 2021-11-24 |  | 7.05 | 12.84 | 7.94 | 24.02 | 1.46 | 2.94 |
| 2021-11-25 | 3.49 | 7.16 | 12.85 | 8.5 | 24.6 | 1.51 | 2.99 |
| 2021-11-26 |  | 7.32 | 13.06 | 8.9 | 25.14 | 1.57 | 3.05 |
| 2021-11-27 | 3.88 | 7.38 | 13.19 | 9.27 | 25.82 | 1.6 | 3.09 |
| 2021-11-28 | 3.94 | 7.43 | 13.28 | 9.46 | 26.24 | 1.61 | 3.12 |
| 2021-11-29 | 4.14 | 7.46 | 13.56 | 9.9 | 26.71 | 1.67 | 3.17 |
| 2021-11-30 | 4.36 | 7.73 | 13.89 | 10.49 | 27.28 | 1.74 | 3.25 |
| 2021-12-01 | 4.67 | 7.88 | 14.25 | 11.13 | 27.88 | 1.82 | 3.33 |
| 2021-12-02 | 4.99 | 8.05 | 14.64 | 11.77 | 28.5 | 1.92 | 3.41 |
| 2021-12-03 | 5.22 | 8.26 | 15.06 | 12.38 | 29.07 | 2.02 | 3.89 |
| 2021-12-04 | 5.38 | 8.4 | 15.27 | 12.88 | 29.75 | 2.06 | 3.95 |
| 2021-12-05 | 5.46 | 8.42 | 15.38 | 13.12 | 30.17 | 2.08 | 3.98 |
| 2021-12-06 |  | 8.63 | 15.7 | 13.74 | 30.66 | 2.17 | 4.06 |
| 2021-12-07 | 5.65 | 8.86 | 16.03 | 14.33 | 31.23 | 2.26 | 4.13 |
| 2021-12-08 | 5.96 | 9.03 | 16.37 | 14.99 | 31.84 | 2.37 | 4.21 |
| 2021-12-09 |  | 9.28 | 16.71 | 15.66 | 32.53 | 2.49 | 4.31 |
| 2021-12-10 | 6.43 | 9.49 | 17.08 | 16.4 | 33.15 | 2.6 | 4.71 |
| 2021-12-11 | 6.6 | 9.56 | 17.26 | 17.03 | 33.96 | 2.66 | 4.78 |
| 2021-12-12 | 6.71 | 9.6 | 17.37 | 17.93 | 34.54 | 2.7 | 4.86 |
| 2021-12-13 | 6.99 | 9.8 | 17.65 | 18.59 | 35.3 | 2.99 | 4.94 |
| 2021-12-14 | 7.28 | 10.01 | 17.96 | 19.43 | 36.26 | 3.37 | 5.05 |
| 2021-12-15 | 7.68 | 10.17 | 18.27 | 20.29 | 37.35 | 3.82 | 5.17 |
| 2021-12-16 | 8.03 | 10.41 | 18.59 | 21.13 | 38.62 | 4.33 | 5.28 |
| 2021-12-17 | 8.37 | 10.6 | 18.95 | 21.86 | 39.87 | 4.88 | 5.4 |
| 2021-12-18 | 8.56 | 10.66 | 19.14 | 22.56 | 41.24 | 5.15 | 5.52 |
| 2021-12-19 | 8.7 | 10.68 | 19.25 | 22.99 | 42.49 | 5.31 | 5.61 |
| 2021-12-20 | 9.05 | 10.9 | 19.58 | 23.69 | 43.8 | 5.86 | 5.74 |
| 2021-12-21 |  | 11.11 | 19.94 | 24.46 | 45.22 | 6.47 | 5.88 |
| 2021-12-22 | 9.39 | 11.34 | 20.28 | 25.2 | 46.45 | 7.12 | 6 |
| 2021-12-23 | 9.72 | 11.54 | 20.53 | 25.69 | 47.34 | 7.7 | 6.11 |
| 2021-12-24 | 9.98 |  | 20.6 | 25.84 | 47.59 | 7.7 | 6.15 |
| 2021-12-25 |  |  | 20.6 | 26.13 | 47.61 | 7.7 | 6.19 |
| 2021-12-26 | 10 |  | 20.67 | 26.31 | 47.64 | 8.12 | 6.22 |
| 2021-12-27 | 10.07 | 11.78 | 20.93 | 26.8 | 48.04 | 8.21 | 6.32 |
| 2021-12-28 | 11.01 | 12.04 | 21.21 | 27.44 | 48.52 | 8.32 | 6.42 |
| 2021-12-29 |  | 12.25 | 21.49 | 28 | 49.15 | 8.75 | 6.53 |
| 2021-12-30 | 12.1 | 12.39 | 21.75 | 28.48 | 49.74 | 9.23 | 6.63 |
| 2021-12-31 | 12.22 | 12.42 | 21.85 | 28.64 | 49.98 | 9.23 | 6.68 |
| 2022-01-01 | 12.26 | 12.42 | 21.86 | 29.03 | 50.01 | 9.23 | 6.71 |
| 2022-01-02 | 12.46 | 12.43 | 21.94 | 29.28 | 50.17 | 9.81 | 6.74 |
| 2022-01-03 |  | 12.63 | 22.14 | 29.82 | 50.38 | 9.95 | 6.82 |
| 2022-01-04 | 13.83 | 12.92 | 22.37 | 30.46 | 50.71 | 10.63 | 6.93 |
| 2022-01-05 |  | 13.18 | 22.6 | 31.03 | 51.07 | 11.5 | 7.03 |
| 2022-01-06 | 15.22 |  | 22.85 | 31.57 | 51.41 | 12.4 | 9.8 |
| 2022-01-07 |  |  | 23.12 | 32.14 | 51.74 | 13.28 | 9.9 |
| 2022-01-08 | 16.31 |  | 23.28 | 32.99 | 52.08 | 13.83 | 10 |
| 2022-01-09 | 16.62 | 13.87 | 23.36 | 33.44 | 52.29 | 14.16 | 10.08 |
| 2022-01-10 | 17.22 | 14.17 | 23.56 | 34.01 | 52.51 | 15 | 10.19 |
| 2022-01-11 | 17.91 | 14.47 | 23.77 | 34.63 | 52.71 | 15.94 | 10.29 |
| 2022-01-12 | 18.61 | 14.75 | 23.97 | 35.28 | 52.9 | 16.92 | 10.41 |
| 2022-01-13 | 19.31 | 15.01 | 24.17 | 36.54 | 53.06 | 17.91 | 10.69 |
| 2022-01-14 | 20.01 | 15.28 | 24.4 | 37.11 | 53.23 | 18.86 | 10.81 |
| 2022-01-15 | 20.38 | 15.36 | 24.53 | 37.74 | 53.38 | 19.38 | 10.9 |
| 2022-01-16 | 20.66 | 16.32 | 24.58 | 38.09 | 53.47 | 19.68 | 11 |
| 2022-01-17 |  | 17.63 | 24.69 | 38.54 | 53.58 | 20.62 | 11.12 |
| 2022-01-18 | 21.31 | 17.85 | 24.78 | 39.09 | 53.69 | 21.53 | 11.22 |
| 2022-01-19 | 22 | 18.31 | 24.79 | 39.54 | 53.79 | 22.48 | 11.32 |
| 2022-01-20 | 23.3 | 18.61 | 24.79 | 39.93 |  | 23.51 | 11.41 |

## Results and discussion

The DCC values calculated with the use of (1), (2) and datasets presented in Tables 1 and 2 are shown in Figs. 1 and 2 for northern and southern regions, respectively. Dashed and solid lines correspond to the years 2020-2021 and 2021-2022, respectively. The vaccination levels from Tables 5 and 6 are represented by “circles” and “crosses”, respectively. Fig. 1 illustrates that for the Nordic countries and the world as a whole, the DCC values in 2021 mostly exceeded the corresponding numbers in 2020, despite the relatively high levels of vaccinations (compare solid and dashed lines). The case of the United Kingdom is particularly significant, where the DCC values differed 1.5-25 times at the highest percentages of vaccinated individuals and persons who received a booster dose of a vaccine.

**Fig. 1.**
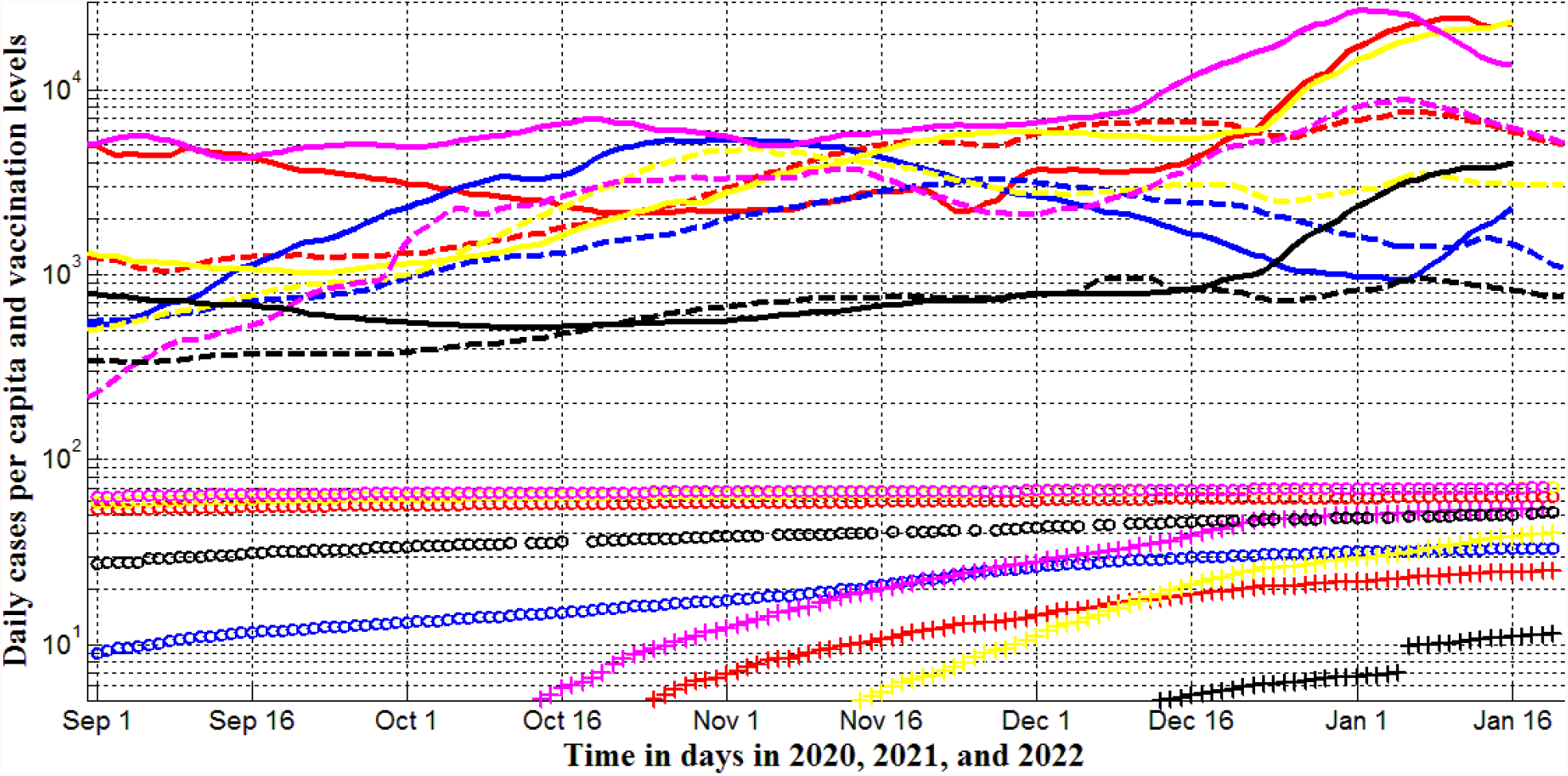
Averaged daily number of new cases per capita and vaccination levels for northern region in September-January, 2020, 2021, and 2022 (Ukraine – blue, EU-yellow, USA – red, UK – magenta, the whole world –black). Lines correspond to DCC*10 values calculated with the use of eqs. (1), (2), Table 1 (2020 and 2021, dashed), and Table 2 (2021 and 2022, solid). “Circles” and “crosses” show the numbers of fully vaccinated people and boosters per hundred, respectively (2021 and 2022, Tables 5 and 6).

**Fig. 2.**
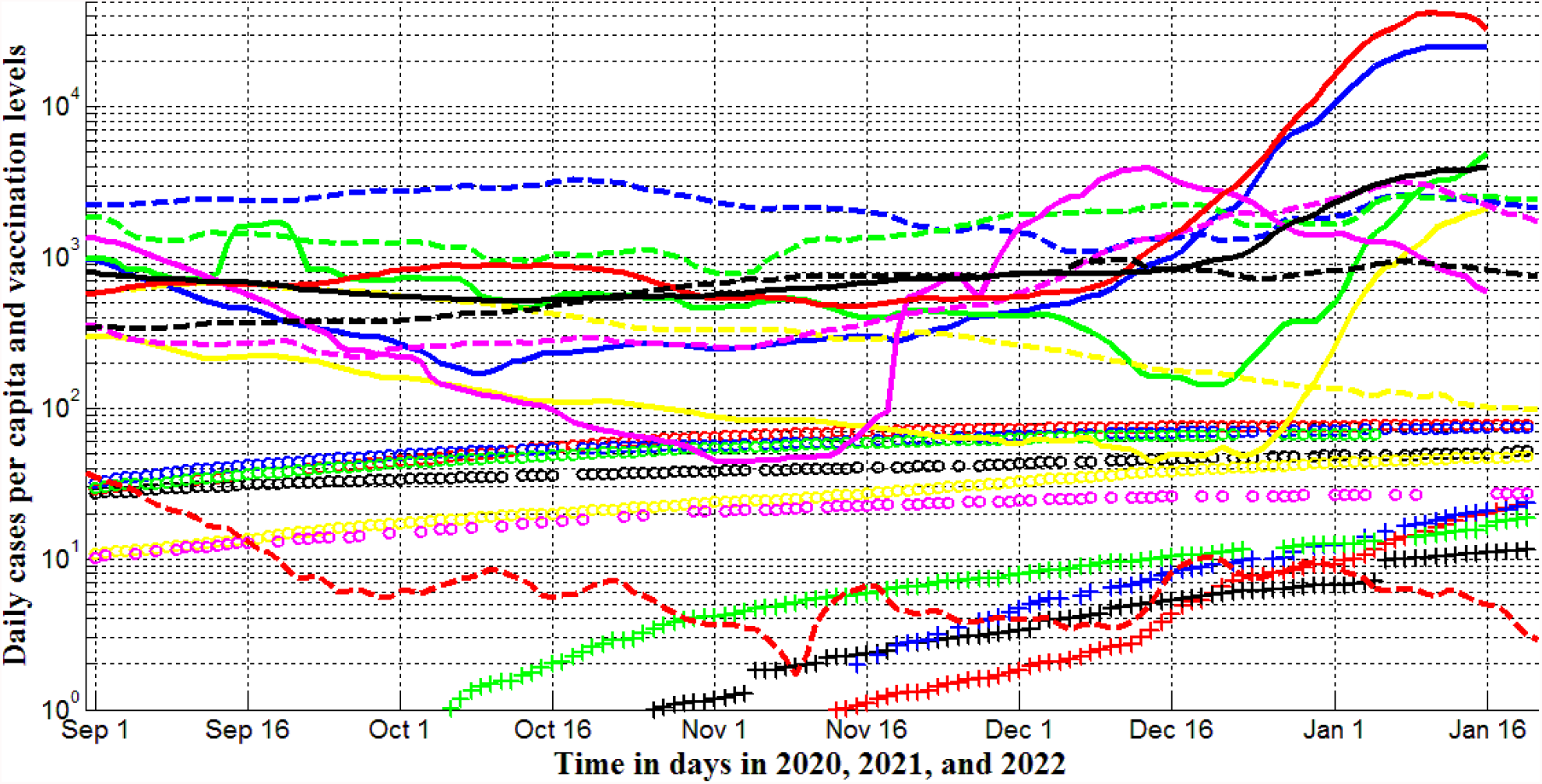
Averaged daily number of new cases per capita and vaccination levels for southern region in September-January, 2020, 2021, and 2022 (Argentina – blue, India - yellow, Brazil – green, South Africa – magenta, Australia-red, and the whole world –black). Lines correspond to DCC*10 values calculated with the use of eqs. (1), (2), Table 1 (2020 and 2021, dashed), and Table 2 (2021 and 2022, solid). “Circles” and “crosses” show the numbers of fully vaccinated people and boosters per hundred, respectively (2021 and 2022, Tables 5 and 6).

The fact that a rather high vaccination level does not prevent the appearance of new COVID-19 pandemic waves was revealed already in September, 2021 [7]. In particular, the DCC numbers in Israel in the summer of 2021 were much higher than in Ukraine, despite the huge difference in vaccination levels [7, 8]. The statistical analysis performed in [9] revealed no correlation between DCC and VC values. Thus, we can conclude that existing vaccines cannot prevent new infections and vaccinated people can spread the infection as intensively as non-vaccinated ones. Probably, it is connected with new strains of coronavirus and with limited in time effectiveness of antibodies. Many countries have introduced special passports (and/or QR codes) that remove restrictions for vaccinated persons to visit crowded places. Presented results illustrate that this does not make sense, since vaccinated and non-vaccinated persons spread the infection with more or less equal probability. Having fresh PCR (or other) tests can be a much more effective pass to crowded places in order to prevent the spread of infection.

Some interesting trends are visible in Fig. 2. For India, Brazil and Argentina (DCC values in 2020 and early 2021 mostly exceeded the corresponding numbers in 2021and early 2022 (compare solid and dashed yellow, green and blue lines, respectively). In June and July (cold season for southern countries) the DCC values were higher in 2021 in comparison with 2020 for these countries (including South Africa), [6]. Probably we see the influence of the seasonal factor, since the vaccination levels in Southern countries were much lower than in the Northern ones (compare corresponding markers in Figs. 1 and 2).

Probably, a new omicron strain is responsible for very sharp increase of DCC values in South Africa after November 12, 2021 (see the solid magenta line in Fig. 2). Later the omicron caused a sharp increase in DCC values in all the countries and regions taken for our analysis (see solid lines, in Figs. 1 and 2). We see two severe epidemic waves in Ukraine in 2021-2022 (the solid blue line in Fig. 1). Probably, the first one (started in September 2021) was not connected with the omicron. Approximately in one month the daily numbers of new cases start to decrease for the “omicron” waves (see solid magenta and red lines in Fig. 1 and solid magenta, red and blue lines in Fig. 2). In January 2022, we can expect diminishing the DCC numbers in Brazil, India, EU, and worldwide. The maximum of DCC values in Ukraine is expected around February 7, 2022 (see the solid blue line in Fig. 1). More exact and long-term predictions can be done with the use of the generalized SIR model [2, 3, 8, 10, 11]. Recent simulations for Ukraine, Poland, Germany and the whole world [10, 11] were based on the datasets corresponding to the periods before “omicron” waves and have to be updated.

The pandemic dynamics in Australia differs significantly from all other countries and regions considered in our study. The DCC values in 2020-2021 exceed 10-800 times corresponding values in 2021-2022 (compare red solid and dashed lines in Fig. 2). The DDC values were close to zero in 2020-2021 and close to the corresponding values for other countries in 2021-2022 (compare solid and dashed lines in Figs. 3 and 4). This is surprising, because in early 2022, Australia has reached a relatively high level of vaccination (see Tables 5 and 6, “circles” and “crosses” in Figs. 1-6). The explanation for these facts should be sought in the weakened quarantine measures and the reduction in the testing level.

**Fig. 3.**
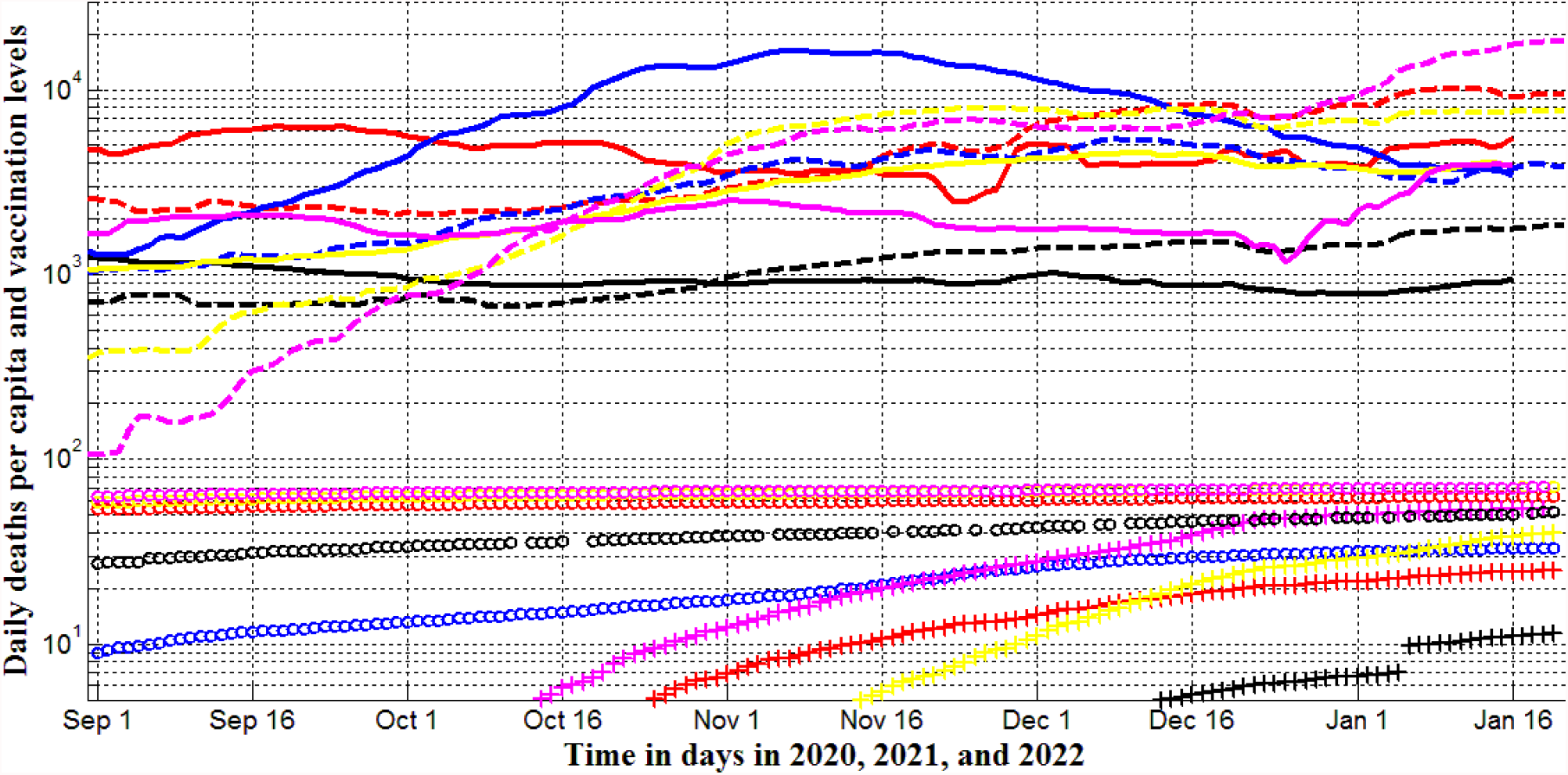
Averaged daily number of deaths per capita and vaccination levels for northern region in September-January, 2020, 2021, and 2022 (Ukraine – blue, EU-yellow, USA – red, UK – magenta, the whole world –black). Lines correspond to DDC*1000 values calculated with the use of eqs. (1), (2), Table 3 (2020 and 2021, dashed), and Table 4 (2021 and 2022, solid). “Circles” and “crosses” show the numbers of fully vaccinated people and boosters per hundred, respectively (2021 and 2022, Tables 5 and 6).

**Fig. 4.**
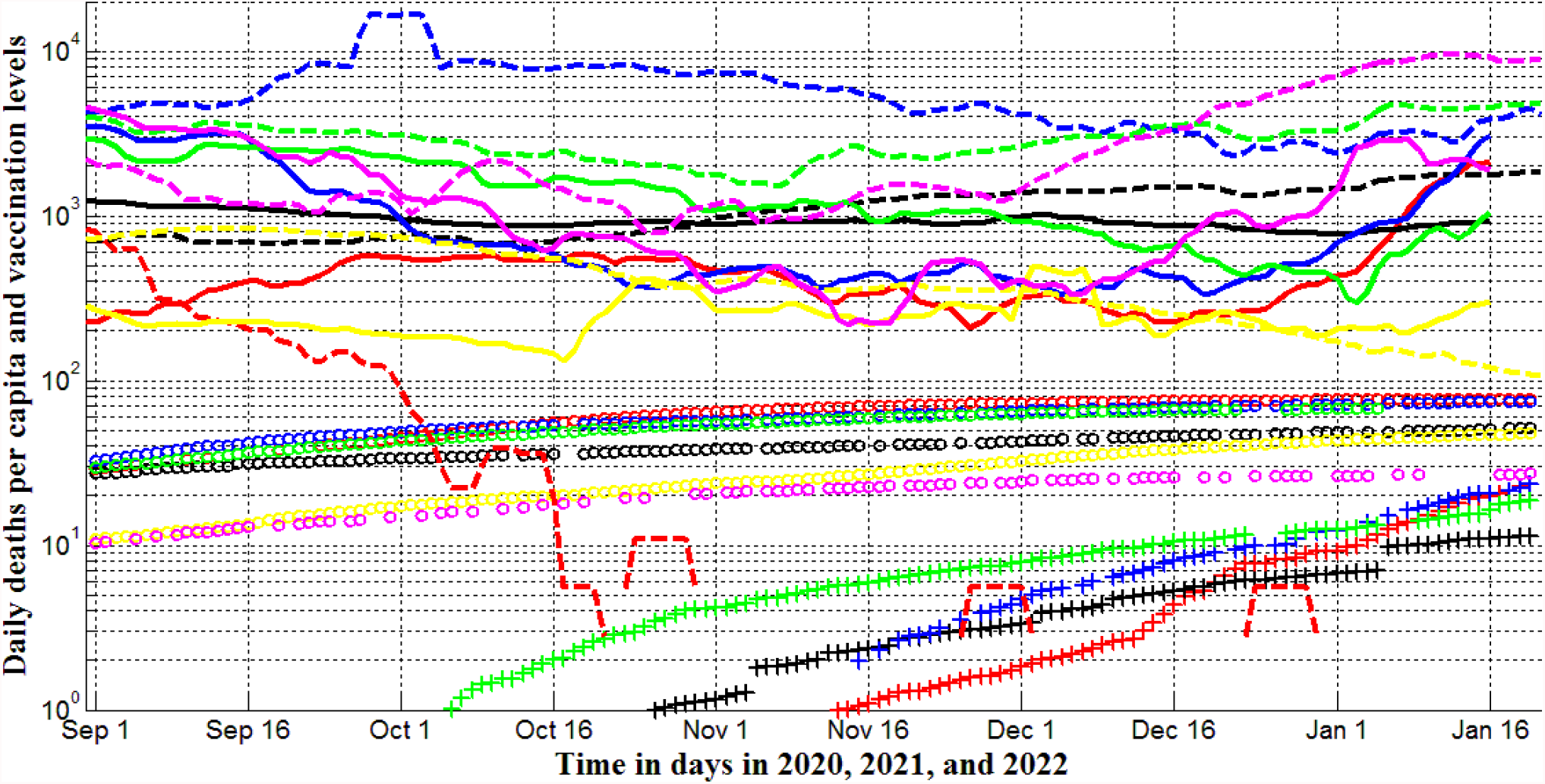
Averaged daily number of new deaths per capita and vaccination levels for southern region in September-January, 2020, 2021, and 2022 (Argentina – blue, India - yellow, Brazil – green, South Africa – magenta, Australia-red, and the whole world –black). Lines correspond to DDC*1000 values calculated with the use of eqs. (1), (2), Table 3 (2020 and 2021, dashed), and Table 4 (2021 and 2022 solid). “Circles” and “crosses” show the numbers of fully vaccinated people and boosters per hundred, respectively (2021 and 2022, Tables 5 and 6).

**Fig. 5.**
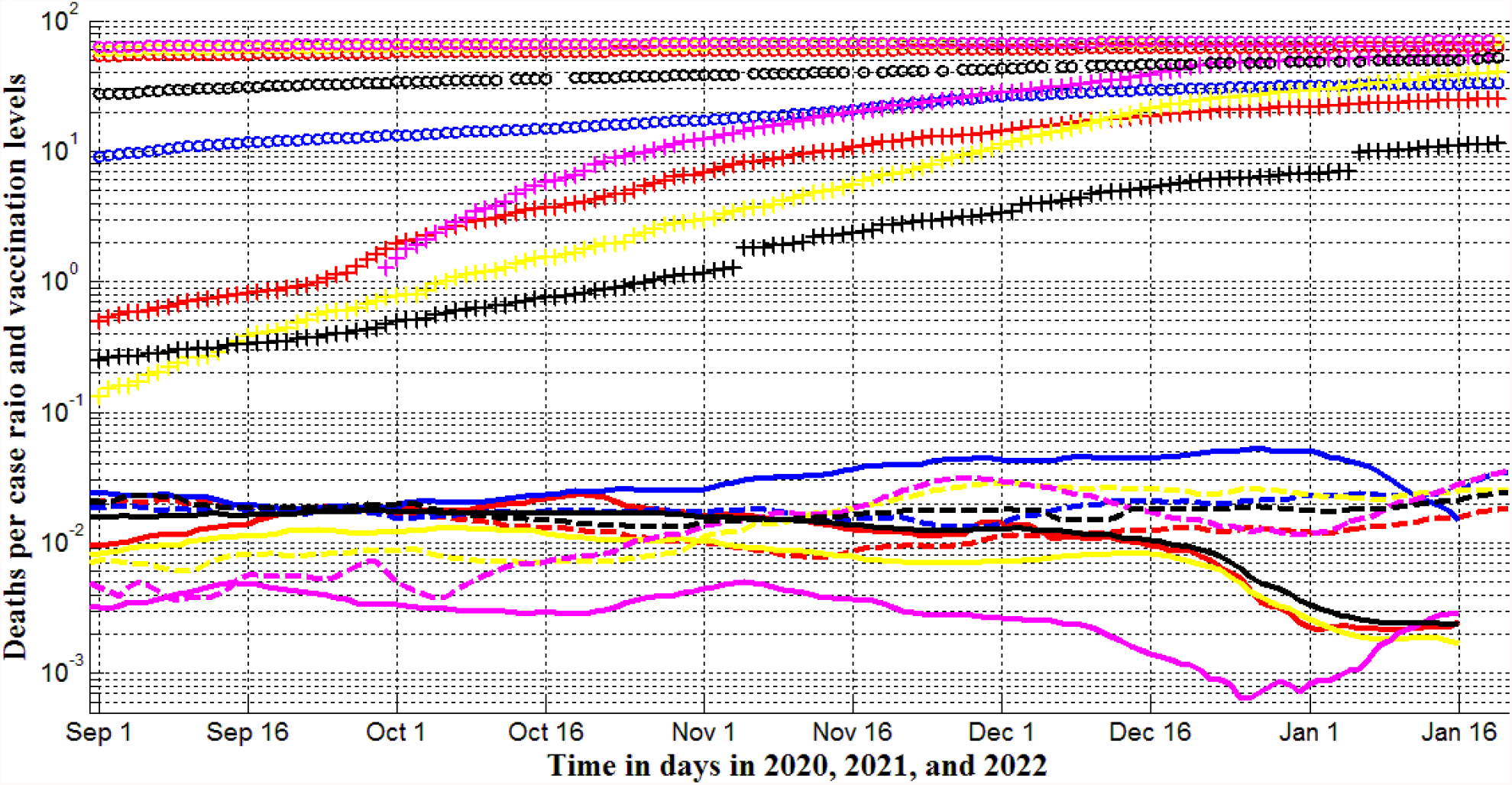
Averaged deaths per case ratios and vaccination levels for northern region in September-January, 2020, 2021, and 2022 (Ukraine – blue, EU-yellow, USA – red, UK – magenta, the whole world –black). Lines correspond to DDC*1000 values calculated with the use of eqs. (1), (2), Table 3 (2020 and 2021, dashed), and Table 4 (2021 and 2022, solid). “Circles” and “crosses” show the numbers of fully vaccinated people and boosters per hundred, respectively (2021and 2022, Tables 5 and 6).

**Fig. 6.**
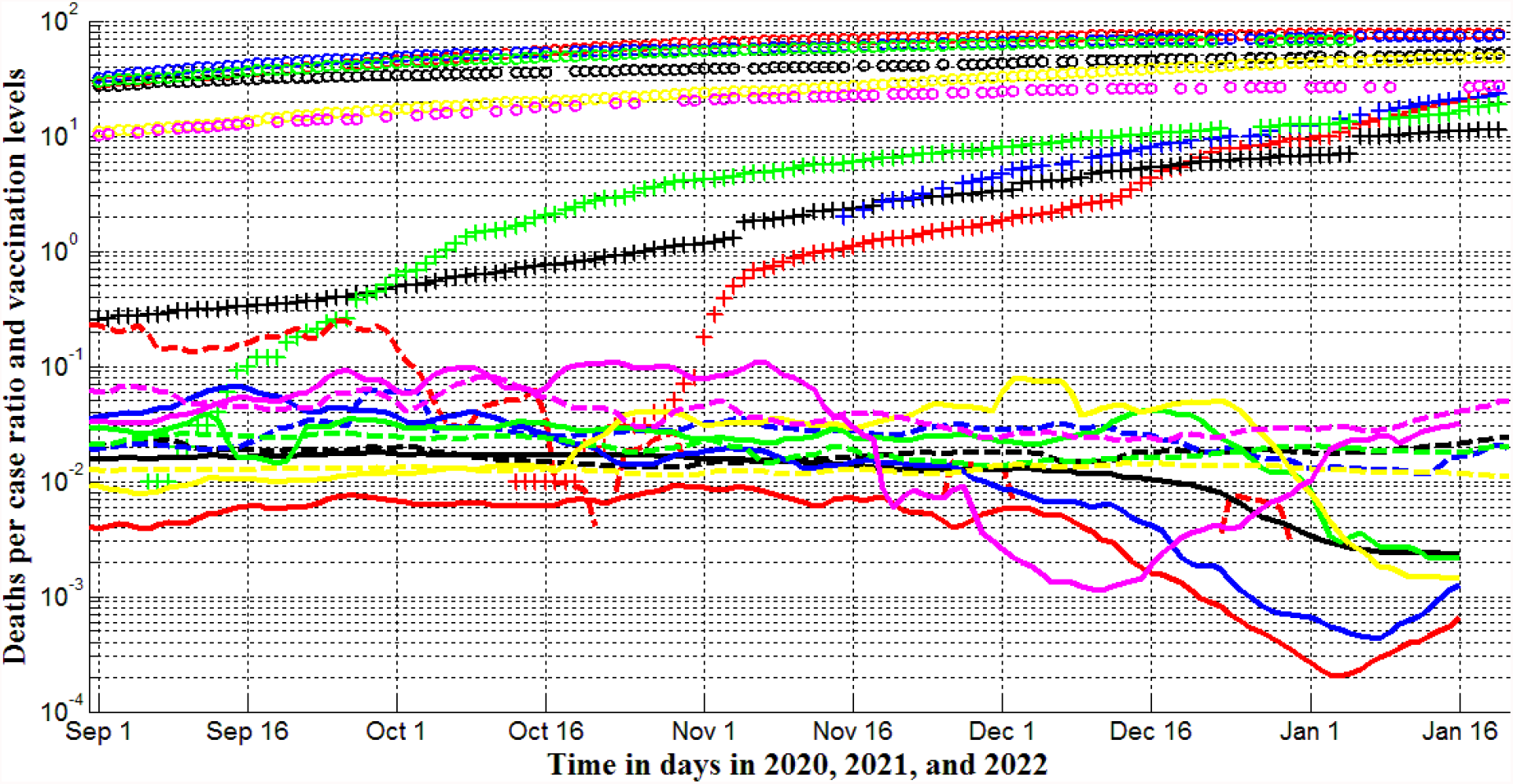
Averaged deaths per case ratios and vaccination levels for southern region in September-January, 2020, 2021, and 2022 (Argentina – blue, India - yellow, Brazil – green, South Africa – magenta, Australia-red, and the whole world –black). Lines correspond to DDC/DCC values calculated with the use of eqs. (1), (2), Table 3 (2020 and 2021, dashed), and Table 4 (2021 and 2022, solid). “Circles” and “crosses” show the numbers of fully vaccinated people and boosters per hundred, respectively (2021 and 2022, Tables 5 and 6).

According to the statistical analysis performed in [9], the numbers of tests per capita TC do not affect the CC, DC, and CC/DC values, but the increase in the number of tests per case TC/CC can reduce CC and DC values. In particular, CC figures can tend to zero when TC/CC exceeds some critical value 520 (in the case of DC, the critical values TC/CC are 460 and 198 for the whole world and Europe, respectively), [9]. In Australia, the TC/CC figures were close to the critical values before September 2021 (394.45, December 30, 2020 and 562.6, September 1, 2021), but later they start to decrease rapidly (238.83, November 14, 2021 and 28.54, January 20, 2022; all the values were calculated with the use of information available in [5]). Where the level of testing was consistently maintained above the critical level (for example, in Hong Kong TC/CC=1984.5 on August 31, 2021 and TC/CC=2386.1 on December 28, 2021; [5]), almost complete control of the pandemic was achieved (the smoothed daily numbers of new cases per million were 0.681 on September 1, 2021 and 1.778 on December 31, 2021 in Hong Kong, [5]). Thus, the significant increase of the testing levels can be recommended as a unique way to stop the pandemic. Unfortunately, existing vaccines cannot do this.

Nevertheless, vaccinations reduce mortality. Figs. 3 and 4 illustrate the influence of vaccination levels on the daily numbers of deaths per capita. In October-November 2021 the DDC values became smaller than corresponding values in 2020 for UK, USA, EU and the whole world (compare solid and dashed magenta, red, yellow, and black lines in Fig. 3) These countries and regions have much higher vaccination levels than Ukraine (see markers in Fig. 3), where DDC values in 2021 were much higher than in 2020 (compare blue solid and dashed lines in Fig. 3). In Argentina and Brazil the DDC values were smaller in 2021 in comparison with 2020 (compare solid and dashed blue and green lines in Fig. 4). In less vaccinated India and South Africa (see markers in Fig. 4), DDC values in 2021 were sometimes higher than in 2020 (compare solid and dashed yellow and magenta lines in Fig. 4). These facts support the conclusion of paper [7] that vaccinations can diminish the numbers of death per capita. The efficiency of boosters needs special statistical analysis.

The case of Australia (red lines and markers in Fig. 4) was already discussed before. Due to the very high testing level TC/CC, this country had almost zero level of mortality in 2020 (see the dashed red line in Fig. 4). After a rapid diminishing of the TC/CC ratio after September 2021, the DDC values increased drastically (see the solid red line) despite a fairly high level of vaccinations (see red markers in Fig. 4).

The number of deaths per case is an important characteristic indicating the probability of death in the case of infection. Knowing of this ratio is important for individuals in order to decide on their vaccination. Comparing these mortality rates in different countries with the same levels of vaccinations allows making conclusions about the efficiency of treatments. The average DDC/DCC values calculated with the of smoothed DDC and DCC numbers (according to eqs. (1) and (2) and Tables 1-4) are shown in Figs. 5 and 6 for northern and southern regions, respectively.

A positive impact of vaccinations is visible almost for all the countries and regions. Solid lines indicate decrease of mortality in 2021-2022 (compare with dashed lines corresponding to 2020-2021). Opposite trend is visible for Ukraine (compare blue dashed and solid lines in Fig. 5) and can be explained by the lower levels of vaccination (compare markers in Fig. 5). The increase of DCC/DCC values in South Africa (see the magenta solid line in Fig. 6) can be also explained by insufficient level of vaccination in this country (compare markers in Fig. 6).

The mortality rates DDC/DCC versus percentage of fully vaccinated people (“circles”) and boosters (“crosses”) are shown in Figs. 7 and 8 for northern and southern regions, respectively. To illustrate the “omicron” waves we use large markers for the period after November 12, 2021. The best fitting lines, linking the DDC/DCC values with the percentage of fully vaccinated people VC can be written as follows:

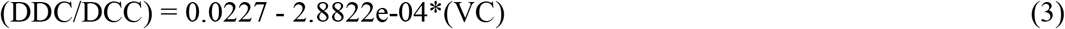

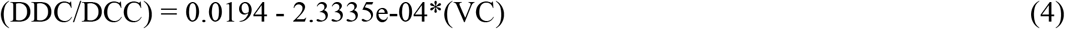

They are shown in Figs. 7 and 8 by solid and dashed lines, respectively. Relationships (3) and (4) are the results of correlation analysis performed in [9] for countries and regions in the whole world and Europe, respectively with the use of JHU datasets corresponding September 1, 2021.

**Fig. 7.**
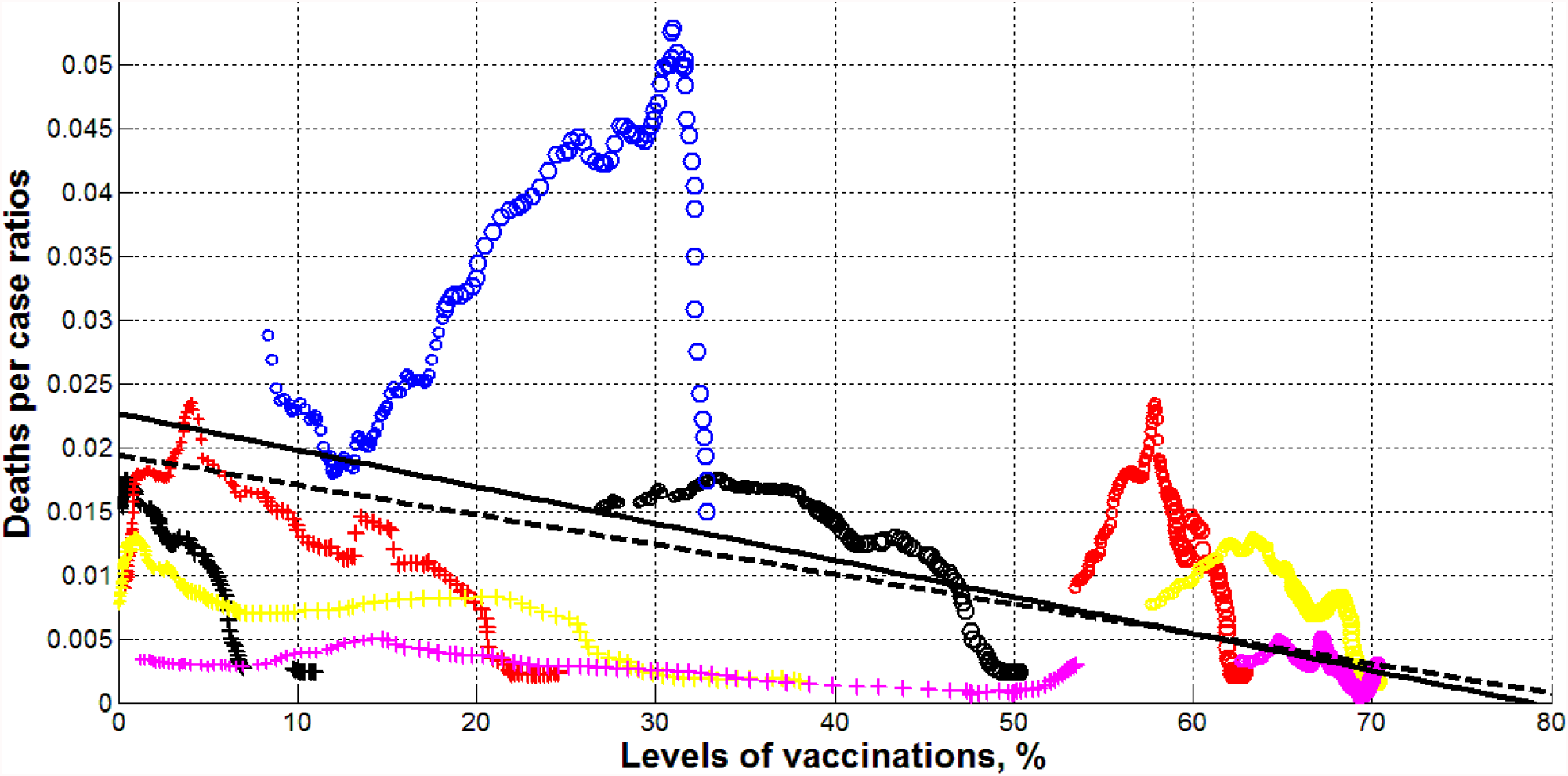
Averaged deaths per case ratios versus vaccination levels for northern region (Ukraine – blue, EU-yellow, USA – red, UK – magenta, the whole world –black). “Circles” and “crosses” correspond to the numbers of fully vaccinated people and boosters per hundred, respectively (Tables 5 and 6). Small markers correspond to the period before November 12, 2021; large – for later period. Black solid and dashed lines represent correlation equations (3) (for the whole world) and (4) (for Europe), respectively.

**Fig. 8.**
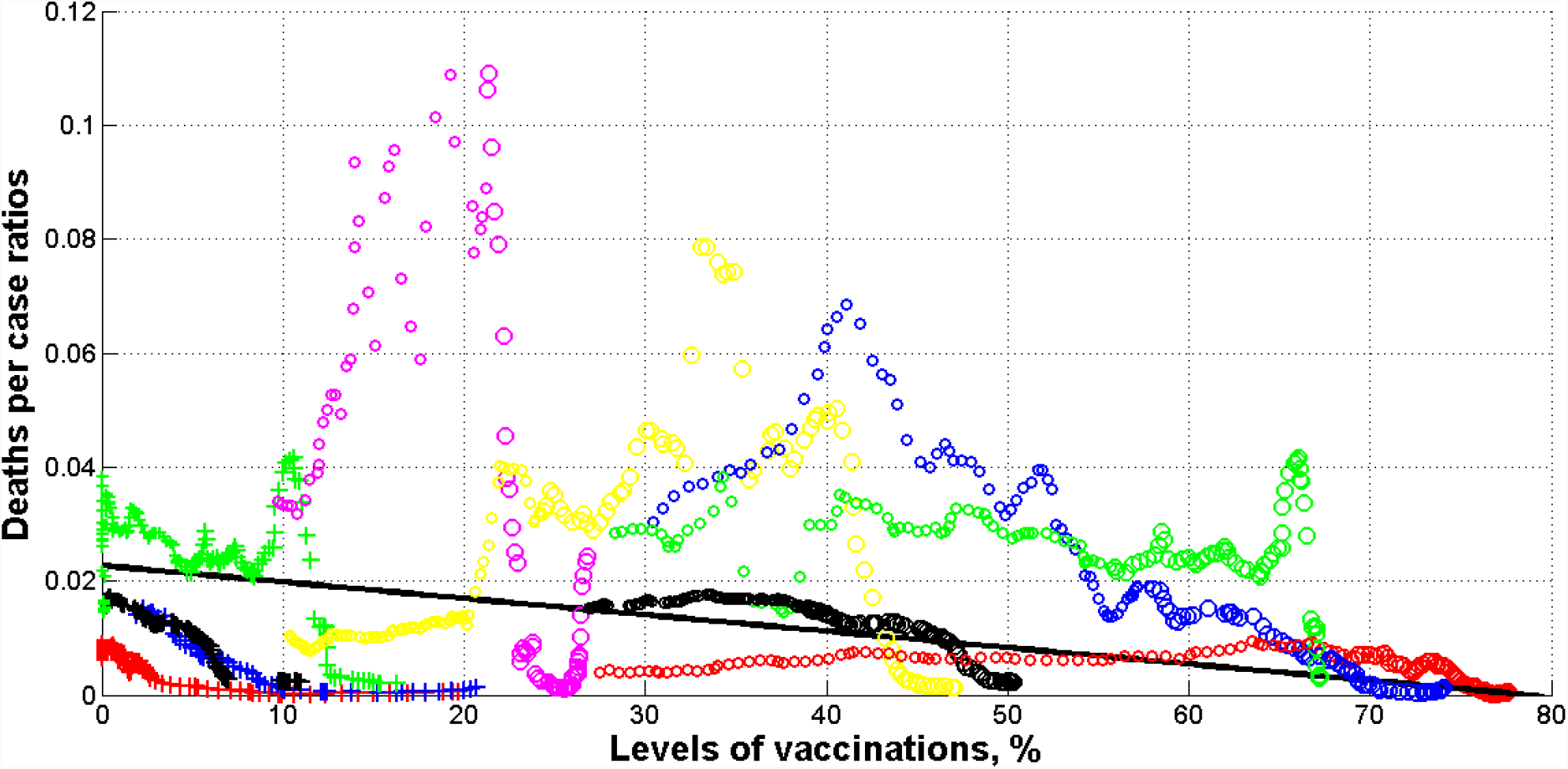
Averaged deaths per case ratios versus vaccination levels for southern region in September-January, 2020, 2021, and 2022 (Argentina – blue, India - yellow, Brazil – green, South Africa – magenta, Australia-red, and the whole world –black). “Circles” and “crosses” correspond to the numbers of fully vaccinated people and boosters per hundred, respectively (Tables 5 and 6). Small markers correspond to the period before November 12, 2021; large – for later period. The black solid line represent correlation equations (3) for the whole world.

It can be seen that averaged DDC/DCC values can be rather scattered, especially for countries or periods with low vaccination levels : Ukraine (blue “circles” in Fig. 7), South Africa (magenta “circles” in Fig. 8), India (yellow “circles” in Fig. 8), Argentina (blue “circles” in Fig. 8). Nevertheless, the mortality rate diminishes with the increase of VC values and tends to the relationships (3) and (4). The same trend is visible for the percentage of boosters (see “crosses” in Figs. 7 and 8). The only one exception is the UK (see magenta “crosses” in Fig. 8). This country has the highest level of boosters, nevertheless the DDC/DCC values have started to increase in January 2022. In any case, the influence of boosters needs special investigations.

## Conclusions

The third year of the pandemic allowed us to compare the pandemic dynamics in the period from September 2020 to January 2021 with the same period one year later for Ukraine, EU, the UK, USA, India, Brazil, South Africa, Argentina, Australia, and in the whole world. The presented results indicate the feasibility of vaccination, although it does not prevent infection. But those who are vaccinated are much less likely to die (this may also indicate a milder course of the disease).

## Data Availability

All data produced in the present work are contained in the manuscript

## References

1. Nesteruk I. Coronasummer in Ukraine and Austria. [Preprint.] ResearchGate. 2020 June. DOI: 10.13140/RG.2.2.32738.56002

2. Nesteruk I. COVID19 pandemic dynamics. Springer Nature, 2021. https://link.springer.com/book/10.1007/978-981-33-6416-5

3. Nesteruk I. Detections and SIR simulations of the COVID-19 pandemic waves in Ukraine. Comput. Math. Biophys. 2021;9:46–65. https://doi.org/10.1515/cmb-2020-0117

4. World Health Organization. “ Coronavirus disease (COVID-2019) situation reports”. https://www.who.int/emergencies/diseases/novel-coronavirus-2019/situation-reports/. Retrieved Oct. 3. 2020.

5. COVID-19 Data Repository by the Center for Systems Science and Engineering (CSSE) at Johns Hopkins University (JHU). https://github.com/owid/covid-19-data/tree/master/public/data

6. Nesteruk I, Rodionov O, Nikitin A.V, and Walczak S. Influences of seasonal and demographic factors on the COVID-19 pandemic dynamics. EAI Endorsed Transactions on Bioengineering and Bioinformatics. Published online Dec 8. 2021. DOI: 10.4108/eai.8-12-2021.172364

7. Nesteruk I. Comparison of the COVID-19 pandemics dynamics in Ukraine and Israel in the summer of 2021. ResearchGate (Preprint), September, 2021 DOI: 10.13140/RG.2.2.29989.22249

8. Nesteruk I. Influence of Possible Natural and Artificial Collective Immunity on New COVID-19 Pandemic Waves in Ukraine and Israel. Explor Res Hypothesis Med 2021; Published online: Nov 11, 2021. doi: 10.14218/ERHM.2021.00044. https://www.xiahepublishing.com/2472-0712/ERHM-2021-00044

9. Nesteruk I, Rodionov O. Impact of Vaccination and Testing Levels on the Dynamics of the COVID-19 Pandemic and its Cessation. J Biomed Res Environ Sci. 2021 Nov 23; 2(11): 1141–1147. doi: 10.37871/jbres1361. Article ID: JBRES1361. Available at: https://www.jelsciences.com/articles/jbres1361.pdf

10. Nesteruk I. Final sizes and durations of new COVID-19 pandemic waves in Ukraine and around the world predicted by generalized SIR model. MedRxiv. Posted November 22, 2021.doi: https://doi.org/10.1101/2021.11.22.21266683

11. Nesteruk I. Final sizes and durations of new COVID-19 pandemic waves in Poland and Germany predicted by generalized SIR model. MedRxiv. Posted December 14, 2021.doi: https://doi.org/10.1101/2021.12.14.21267771

